# From Big Data to the clinic: methodological and statistical enhancements to implement the UK Biobank imaging framework in a memory clinic

**DOI:** 10.1101/2024.08.02.24311402

**Authors:** Grace Gillis, Gaurav Bhalerao, Jasmine Blane, Robert Mitchell, Pieter M Pretorius, Celeste McCracken, Thomas E Nichols, Stephen M Smith, Karla L Miller, Fidel Alfaro-Almagro, Vanessa Raymont, Lola Martos, Clare E Mackay, Ludovica Griffanti

## Abstract

**Introduction:** The analysis tools and statistical methods used in large neuroimaging research studies differ from those applied in clinical contexts, making it unclear whether these techniques can be translated to a memory clinic setting. The Oxford Brain Health Clinic (OBHC) was established in 2020 to bridge this gap between research studies and memory clinics.

**Methods:** We optimised the UK Biobank imaging framework for the memory clinic setting by integrating enhanced quality control (QC) processes (MRIQC, QUAD, and DSE decomposition) and supplementary dementia-informed analyses (lobar volumes, NBM volumes, WMH classification, PSMD, cortical diffusion MRI metrics, and tract volumes) into the analysis pipeline. We explored associations between resultant imaging-derived phenotypes (IDPs) and clinical phenotypes in the OBHC patient population (N=213), applying hierarchical FDR correction to account for multiple testing.

**Results:** 14-24% of scans were flagged by automated QC tools, but upon visual inspection, only 0-2.4% of outputs were excluded. The pipeline successfully generated 5683 IDPs aligned with UK Biobank and 110 IDPs targeted towards dementia-related changes. We replicated established associations and found novel associations between brain metrics and age, cognition, and dementia-related diagnoses.

**Conclusion:** The imaging protocol is feasible, acceptable, and yields high-quality data that is usable for both clinical and research purposes. We validated the use of this methodology in a real-world memory clinic population, which demonstrates the potential of this enhanced pipeline to bridge the gap between big data studies and clinical settings.

**Key Points:**

1. The imaging methods, analysis techniques, and population characteristics in research studies often differ to those in traditional clinical settings.
2. To bridge this gap, we optimised the UK Biobank imaging framework for memory clinic use by integrating enhanced quality control (QC) and supplementary analyses targeted towards dementia-related changes.
3. We generated 5683 imaging-derived phenotypes (IDPs) aligned with UK Biobank and 110 supplementary dementia-informed IDPs that captured both established and novel associations between brain metrics and dementia-related clinical phenotypes, highlighting the value of integrating UK Biobank-aligned imaging and analyses in a real-world memory clinic population.

## 1 Introduction

State-of-the-art neuroimaging is included in large population studies like UK Biobank to assess baseline ‘brain health’ and to predict the risk of diseases and disorders such as Alzheimer’s Disease (AD) (Harms et al., 2018; Miller et al., 2016). Major advances in image analysis methodology and statistical methods have been developed within the UK Biobank imaging substudy (Alfaro-Almagro et al., 2018, 2021), meaning that well-powered conclusions can be drawn about the relationship between brain structure and function and a range of health and lifestyle factors. However, these advances widen the (already substantial) gap between neuroimaging methodology available in research settings and those available in clinical practice.

Structural brain imaging is included in the diagnostic guidelines for dementia, typically as a computerised tomography (CT) scan used primarily for ruling out alternative causes of cognitive impairment (Jack et al., 2016; National Institute for Health and Care Excellence (NICE), 2018). Compared to CT, structural magnetic resonance imaging (MRI) offers higher resolution and contrast, with greater sensitivity to brain changes like atrophy and white matter hyperintensities, both of which are hallmarks of AD and other forms of dementia (Harper et al., 2015). There are automated analysis tools to quantify these structural changes, including commercial software (Pemberton et al., 2021), but standard radiology practice continues to rely on qualitative or semi-quantitative methods using visual rating scales (Fazekas et al., 1987; Harper et al., 2015; Scheltens et al., 1995). Advanced MR imaging and analysis have also revealed differences in cerebral blood flow and structural and functional connectivity in patients with cognitive impairment (Liu et al., 2023; Penalba-Sánchez et al., 2023; Teipel & Grothe, 2023; Xiao et al., 2023), but these methods are currently not recommended for memory clinic use since their diagnostic and prognostic value is less established (Egle et al., 2022; Haidar et al., 2023; Vemuri et al., 2012). Thus, our understanding of the links between brain and phenotypic changes is constrained by the data currently available (i.e., cohort studies rather than real-world clinical populations).

Participants in cohort studies, however, tend to be younger and healthier than patients with memory problems, making it difficult to (i) draw conclusions about dementia-related changes and (ii) test the performance of analysis tools in brains representing more advanced disease stages. Dementia cohort studies have been established to address these limitations (e.g., ADNI - Petersen et al., 2010). However, these cohorts are often overused, which limits generalisability (Borchert et al., 2023), and use strict inclusion/exclusion criteria, making the data poorly representative of the overall memory clinic patient population (Langbaum et al., 2023; Lim et al., 2023). Methodological differences further widen the gap between large cohorts and smaller clinical studies. For example, while smaller clinical studies can employ visual inspection to control the quality and accuracy of raw and processed MRI data, big data studies must rely on automated quality control (QC) tools. Moreover, differences in study design (e.g., data-vs hypothesis-driven) and statistical power also require distinct, context-dependent statistical approaches.

To bridge the technological, methodological, and population differences between research studies and memory clinics, the Oxford Brain Health Clinic (OBHC) was established in August 2020 as a joint clinical-research service (O’Donoghue et al., 2023). In South Oxfordshire, National Health Service (NHS) patients with memory concerns are referred by their GPs to an Oxford Health NHS Foundation Trust memory clinic and may subsequently be referred to the OBHC for high-quality multimodal assessments, including an MRI protocol aligned with UK Biobank (Griffanti et al., 2022). The results of these assessments are sent back to the referring memory clinic as a detailed report, which the psychiatrist takes into consideration during the follow-up diagnostic appointment with the patient.

Here, we tested the suitability of employing UK Biobank image acquisition and analysis pipelines in patients referred to the OBHC. We evaluated the feasibility and acceptability of the protocol in this ‘real-world’ population by assessing completion rates and data quality, including the use of automated quality control tools. Pipeline adaptations, previously described (Griffanti et al., 2022), were incorporated to overcome challenges associated with brain changes in older populations, and additional dementia-informed measures were added to the standard set of IDPs.

Finally, the utility of the standard and dementia-informed IDPs was tested by performing a set of well-established associations with age, cognition and diagnosis, including careful consideration of the most appropriate statistical adaptations (i.e., hierarchical false discovery rate procedure) for a clinical population.

## 2 Methods

### 2.1 OBHC Model and Patients

In the UK, NHS patients over 65 with memory concerns are typically referred by their GP to psychiatry-based memory services. In Oxfordshire, these patients may be triaged by the memory clinic and referred to the Oxford Brain Health Clinic (OBHC) if they are able to travel and eligible for an MRI scan. The OBHC appointment involves several high-quality assessments as standard, including a detailed cognitive assessment and an MRI scan, and offers multiple avenues for optional research participation. Patients can consent to join the OBHC Research Database and can choose to complete a range of additional research assessments including: more time in the MRI scanner, a saliva sample for genotyping, and detailed questionnaires for both themselves and their relative. By reducing the barriers to research participation, this model makes the OBHC population highly representative of the ‘real-world’ local memory clinic population. The OBHC Research Database was reviewed and approved by the South Central – Oxford C research ethics committee (SC/19/0404). Please refer to O’Donoghue et al. (2023) for more details about the protocols and assessments.

Assessment results are compiled into a report which is used by clinicians to support their diagnostic decision during the patient’s subsequent memory clinic visit. As of May 2023, 213 patients were MRI scanned as part of their NHS assessment at the OBHC and consented to the use of their data for research purposes.

### 2.2 Joint Clinical-Research Neuroimaging Protocol

The MRI scanning protocol used at the OBHC is available in Supplementary Table 1. This protocol is aligned with the UK Biobank brain imaging protocol (Miller et al., 2016). It is restructured to prioritise the modalities with known clinical relevance (‘core clinical’ sequences: T1-weighted, T2 fluid attenuated inversion recovery - T2-FLAIR, and susceptibility-weighted - swMRI) whilst enabling consenting patients to stay in the scanner for additional research sequences (diffusion MRI - dMRI, arterial spin labelling - ASL, and resting-state functional MRI - rfMRI) (Griffanti et al., 2022).

The core clinical MRI scans for all patients are transferred to the NHS clinical records system and reported by a neuroradiologist (PP), using a structured radiology report template (Griffanti et al., 2022). The resulting report is sent to the referring memory clinic. If the patient gives consent for their data to be used for research, then the image files from all completed scans are pseudonymised and forwarded to a secure research server. Full details of the MRI protocol and operating procedures are available online (O’Donoghue et al., 2022).

### 2.3 Clinical Outcome Data

Cognitive performance is measured during the OBHC appointment with Addenbrooke’s Cognitive Examination III (ACE-III), which assesses cognition across five domains of memory, attention, language, fluency, and visuospatial skills. Patient diagnoses are based on the OBHC assessments, clinician judgement, and any additional assessments completed during the subsequent memory clinic visit, on average 2.5 months after their OBHC appointment. These diagnoses are extracted from secondary care electronic healthcare records (EHRs). For this study, primary diagnoses were categorised as dementia (ICD10 codes F00, F01, F02, F03), mild cognitive impairment (MCI - F06.7), and no dementia-related diagnoses (F10, F31, F32, F41, and patients who attended their memory clinic appointment but did not receive a formal diagnosis).

### 2.4 Pipeline Adaptations

Scans were processed using a version of the UK Biobank image analysis pipeline (Alfaro-Almagro et al., 2018; Stephen Smith et al., 2022). Figure 1 shows a schematic overview of the adapted pipeline. The pipeline was adapted to include lesion-masking of the grey matter segmentations (SIENAX) and CSF-masking of the hippocampal segmentations (FIRST), as previously described (Griffanti et al., 2022), to obtain accurate segmentations in the presence of atrophy and high vascular pathology. Downstream adaptations were also applied to the parts of the pipeline that rely on these corrected segmentations (ASL and VBM analyses). Additional IDPs were added to the pipeline to capture other putative dementia-related changes, as described below.

**Figure 1:**
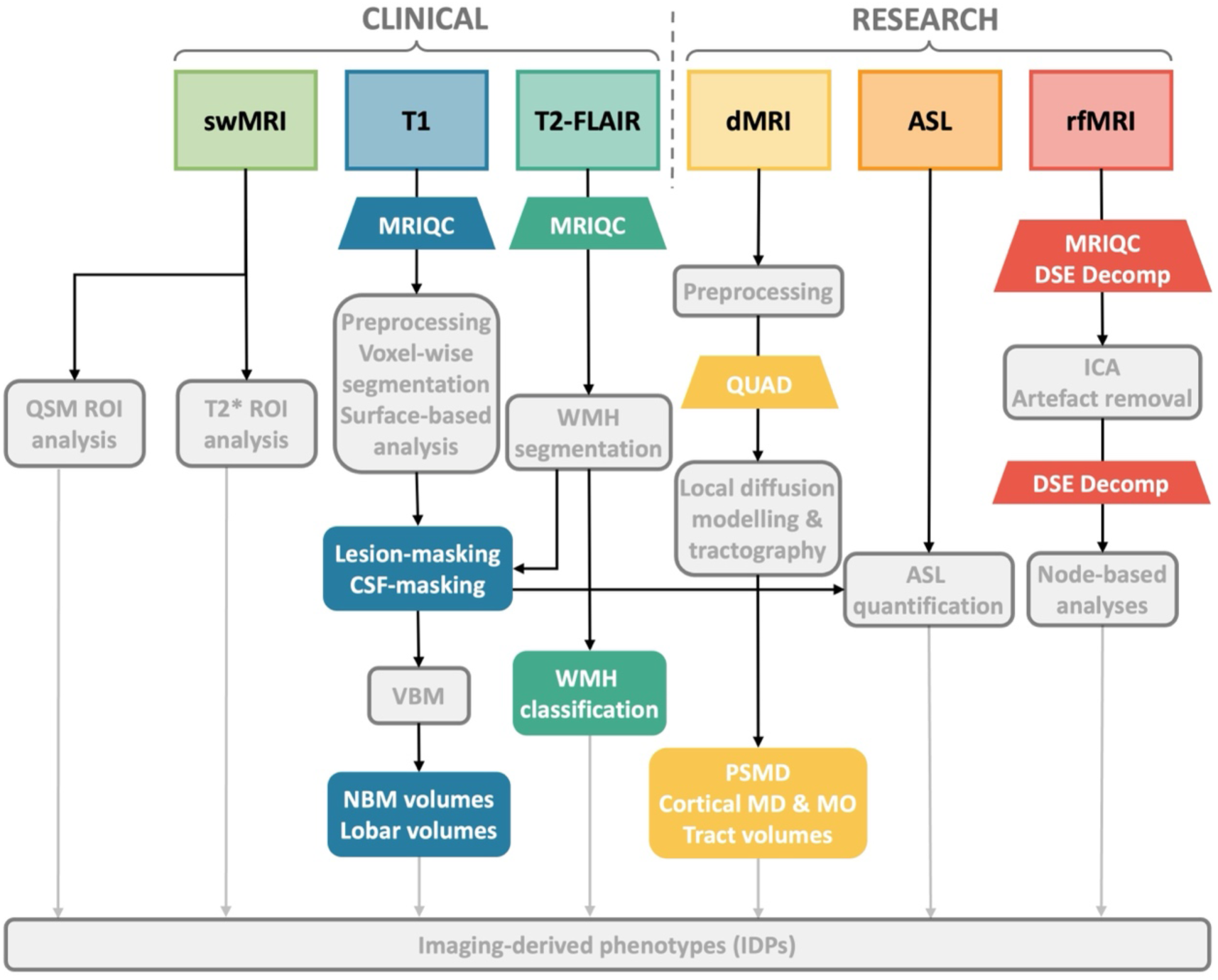
Simplified overview of the UK Biobank image analysis pipeline (in grey) with adaptations and enhancements for OBHC use (in colour). See Alfaro-Almagro et al. (2018) for details of the UKB pipeline. swMRI, susceptibility-weighted MRI; T1, T1-weighted imaging; T2-FLAIR, T2-weighted fluid attenuated inversion recovery imaging; dMRI, diffusion-weighted MRI; ASL, arterial spin labelling; rfMRI, resting-state functional MRI; QSM, quantitative susceptibility mapping; ROI, region of interest; MRIQC, MRI Quality Control tool (see (Esteban et al., 2017)); VBM, voxel-based morphometry; NBM, nucleus basalis of Meynert; WMH, white matter hyperintensity; QUAD, QUality Assessment of DMRI (see (Bastiani et al., 2019)); PSMD, peak width of skeletonised mean diffusivity; MD, mean diffusivity; MO, mode of anisotropy; DSE, D-var, S-var, E-var (see (Afyouni & Nichols, 2018)); ICA, independent component analysis.

#### 2.4.1 Lobar volumes

Predominance of atrophy in one or more lobes can guide differential diagnosis, for example distinguishing between Alzheimer’s disease, semantic dementia, and frontotemporal lobar degeneration (FTLD) (Rabinovici et al., 2007). Masks of the frontal, parietal, temporal, and occipital lobes (MNI structural atlas – see (Collins et al., 1995; Mazziotta et al., 2001)) were non-linearly registered to the T1-weighted scan and applied to the corrected SIENAX grey matter (GM) segmentation to yield lobar GM volumes.

#### 2.4.2 WMH volumes

The default UK Biobank pipeline calculates the periventricular, deep, and total white matter hyperintensity (WMH) volumes derived using BIANCA (Griffanti et al., 2016). Given the relevance of the spatial distributions of WMHs to underlying dementia-related pathologies (Biesbroek et al., 2016, 2024; Veldsman et al., 2020), we additionally generated tract-wise WMH volumes by calculating the overlap of the WMH mask with 48 white matter (WM) tract masks registered to T1 space (Mori et al., 2008).

#### 2.4.3 Cholinergic analyses

Given the relevance of the cholinergic systems in dementia (Bohnen et al., 2018), additional IDPs were added to measure the volumes of the left and right nucleus basalis of Meynert (NBM) in the basal forebrain. A widely used histologically-defined NBM mask (Zaborszky et al., 2008), non-linearly registered to T1 space, was masked with the GM partial volume estimate.

#### 2.4.4 Additional dMRI metrics

The peak width of the skeletonised mean diffusivity (PSMD) is a robust marker of overall cerebrovascular pathology and holds biomarker potential in dementia (Low et al., 2020; Satizabal et al., 2020). It shows good correlations with cognition, explaining variability in cognition beyond traditional measures of cerebrovascular-related WM changes such as WMH volume (Satizabal et al., 2020). PSMD was calculated as the width between the 5^th^ and 95^th^ percentiles of the skeletonised mean diffusivity maps, aligned with the protocol outlined by Baykara and colleagues (2016).

Tractography-defined WM tract volumes have also been proposed as estimates of tract-specific atrophy with moderate subject-differentiating power (Besseling et al., 2012; Groot et al., 2015). Following Warrington and colleagues (2020), we binarized each tractography-defined tract, normalised for the number of valid streamlines and registered to dMRI space, at 0.005 and calculated its volume.

Grey matter mean diffusivity (MD) metrics are also gaining popularity as potential biomarkers in dementia and cognitive decline (Douaud et al., 2011, 2022; Illán-Gala et al., 2019; Montal et al., 2021; Weston et al., 2020). The mode of anisotropy (MO) has demonstrated sensitivity to WM changes in MCI and early dementia (Douaud et al., 2011), although it seems potentially less informative in GM structures (Beer et al., 2020; Douaud et al., 2013). Using the GM partial volume estimate (PVE) after lesion-masking, weighted MD and MO were calculated within the corrected FIRST segmentations of the hippocampi and amygdala as well as 5 ROIs defined with the UKB GM atlas: parahippocampal gyrus, precuneus, superior frontal cortex, superior parietal cortex and supramarginal cortex.

### 2.5 Integrated Quality Control

Automated QC of the scans was integrated into the analysis pipeline where possible. Details of the image-quality metrics (IQMs) included in the different tools are summarised in Table 1. MRIQC (Esteban et al., 2017) was run for T1 and T2-FLAIR, EDDY QUAD (Bastiani et al., 2019) for dMRI, and both MRIQC and DSE decomposition (Afyouni & Nichols, 2018) for rfMRI. Given the importance of T1 as an anatomical reference for other modalities and all analyses, we do perform manual QC using static visual summaries (9 orthogonal slices, generated with the FSL command slicesdir) of T1 scans, brain extracted and registered to MNI space. FIRST outputs are overlaid on the CSF map (output of FAST), and structures with full overlap are identified as mislocalised subcortical segmentations and excluded from subsequent analyses.

**Table 1:** Automated quality control (QC) tools with categories of image-quality metrics (IQMs). For details about the individual measures please refer to Esteban et al. (2017) for MRIQC, Afyouni and Nichols (2018) for DSE and Bastiani et al. (2019) for EDDY QUAD.

| Tool | Image-Quality Metric (IQM) Categories |
| --- | --- |
| **MRIQC:**<br>**T1 & T2-FLAIR** | Noise measures (*CJV for T1, CNR, SNR within CSF, GM, and for T1 WM, QI2*); information theory measures (*EFC, FBER*); bias field measures (*INU_range for T1, INU_med*); carotid vessels/fat hyperintensity (*WM2MAX*) |
| **MRIQC:**<br>**rfMRI** | Noise measures (*SNR, tSNR, GSR*); information theory measures (*EFC, FBER*); global correlation (*GCOR*); outliers (*AOR*); quality (*AQI*) |
| **DSE**<br>**Decomposition:**<br>**rfMRI** | Global and whole % S-var and % D-var (pre- and post-FIX) |
| **EDDY QUAD:**<br>**dMRI** | Head motion; susceptibility; outliers; noise measures |

Scans with outliers (>1.5 IQR away from Q1/Q3) in at least one of the IQMs or visual inspection of the T1 summary were flagged for further inspection of the core pipeline outputs: corrected SIENAX and FIRST segmentations for T1, BIANCA segmentations for T2-FLAIR, tractography from dMRI, and preprocessed DSE decomposition for rfMRI. Low-quality raw or derived images were excluded from subsequent analysis.

### 2.6 Statistics

Numeric variables, including all IDPs, ACE-III scores, and age, were normalised to unit variance using quantile normalisation (Peterson & Cavanaugh, 2020), implemented with the R package bestNormalise (Ryan A. Peterson, 2021). Due to sample size limiting the degrees of freedom compared to UKB, we compromised on the number of covariates and included only age, sex, and head size. We investigated the extent to which IDPs predict age and cognition by performing linear regression with each IDP and covariates (Equations 1-2). We included total cognitive score as a covariate in the linear regression with age (Equation 1) to highlight age-related changes that are not purely related to cognitive impairment in this memory clinic population where cognition may be expected to account for a large share of the observed variance. Associations with diagnoses were assessed with ordinal regression (Equation 3). We additionally investigated associations of clinical phenotypes with T2*, QSM, and tractography IDPs when controlling for ROI/tract volumes.

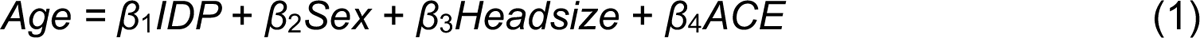

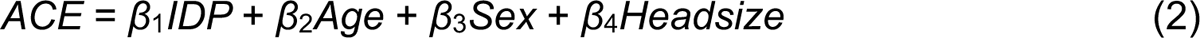

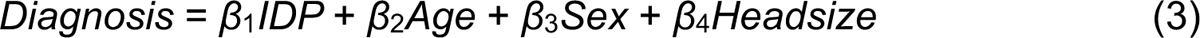

In order to control for multiple testing across modalities while enabling interpretation within modality, we used a hierarchical false discovery rate procedure (Yekutieli, 2008). This guarantees that the false discovery rate (FDR, the expected proportion of false positives among detections) is controlled within each modality. The method proceeds by computing omnibus p-values for each modality with a Simes test, and then the modalities that are significant at the 5% FDR level have their IDPs tested with FDR that uses a more stringent threshold according to the number of significant omnibus p-values.

## 3 Results

### 3.1 Demographics

See Table 2 for OBHC patient demographics. Figure 2 illustrates the prevalence of diagnoses that were obtained subsequently, grouped into three diagnostic categories (dementia, MCI, and no dementia-related diagnosis) along with the distributions of ACE-III cognitive scores for these diagnostic groups.

**Figure 2:**
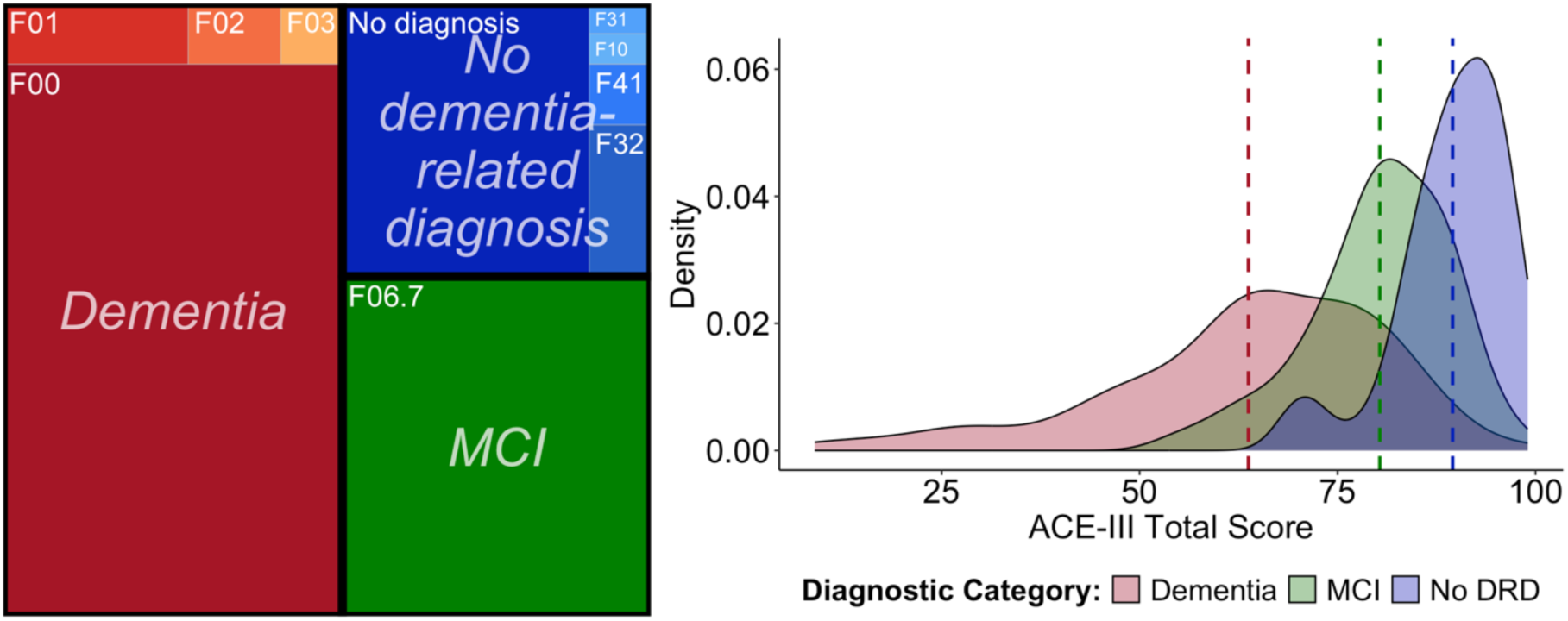
Subsequent diagnoses of OBHC patients and distribution of cognitive scores across the diagnostic groups. Dashed lines on density plot are the group means. F00, Alzheimer’s dementia; F01, vascular dementia; F02, Dementia in other diseases classified elsewhere; F03, unspecified dementia; F06.7, mild cognitive impairment; F31, bipolar disorder; F10, alcohol-related disorders; F41, other anxiety disorders; F32, depressive episode. DRD, dementia-related diagnosis. ACE-III, Addenbrooke’s Cognitive Examination III.

**Table 2:** Demographics of MRI-scanned OBHC patient population: overall and stratified by the subsequent diagnostic category. 1 patient missing a diagnosis was excluded from stratified results. DRD, dementia-related diagnosis; ACE-III, Addenbrooke’s Cognitive Examination III; ApoE, Apolipoprotein E.

| Characteristic | Overall | Stratified by diagnostic category |  |  |
| --- | --- | --- | --- | --- |
| Sample Size | N=213 | Dementia (N=111) | MCI (N=56) | No DRD (N=45) |
| Age (years) – mean ± SD (range) | 78.0 ± 6.2 (65-101) | 79.8 ± 6.5 (65-101) | 76.8 ± 5.5 (66-88) | 74.9 ± 4.7 (65-85) |
| Sex - % F (M/F) | 47.4% (112/101) | 45.9% (60/51) | 53.6% (26/30) | 44.4% (25/20) |
| ACE-III Total Score – mean ± SD (range) | 73.9 ± 17.4 (9-99) | 63.5 ± 17.3 (9-98) | 80.3 ± 8.8 (55-97) | 89.5 ± 6.9 (70-99) |
| Years FT education – mean ± SD (range) [N] | 13.1 ± 3.5 (3-21) [N=192] | 12.9 ± 3.6 (3-21) [N=99] | 12.7 ± 3.5 (4-21) [N=52] | 14.1 ± 3.3 (9-21) [N=40] |
| Clinical Frailty Score – mean ± SD (range) | 2.9 ± 1.4 (1-7) | 3.2 ± 1.5 (1-7) | 2.7 ± 1.2 (1-6) | 2.4 ± 1.1 (1-6) |
| ApoE - % with at least 1 E4 allele [N/total genotyped] | 41.4% [55/133] | 49.2% [31/63] | 37.5% [12/32] | 31.6% [12/38] |

### 3.2 Feasibility and acceptability

As of May 2023, 244 patients attended the OBHC, 93.4% (N=228) of whom consented to join the OBHC Research Database. 93.4% of these patients (N=213) were able to complete at least one of the UKB-aligned MRI sequences (excluded patients had MRI contraindications (N=4), incompatible body habitus/kyphosis (N=4), claustrophobia (N=3), or discomfort/anxiety (N=4)). 211 patients completed all core clinical sequences (swMRI, T1, T2-FLAIR; 16 minutes and 29 seconds), with 2 scans terminated prematurely due to patient discomfort. Of the 165 patients (67.6%) that initially consented to the additional MRI research sequences, 127 were still able and willing to stay in the scanner when asked after the clinical sequences. 120 patients (56.3% of those who started the scanning protocol) completed all sequences (37 minutes and 46 seconds total).

Figure 3 summarises the available MRI data. The high rates of willing consent and completion indicate that the protocol is feasible and acceptable. Integrating this research-quality scanning protocol into clinical workflows via the OBHC also yielded high rates of MRI scans suitable for detailed radiology reporting for clinical purposes. From the 213 patients that started the MRI session, the neuroradiologist completed 212 full structured radiology reports and 1 partial report due to scan termination. One additional scan was noted as poor quality, but this did not impede reporting.

**Figure 3:**
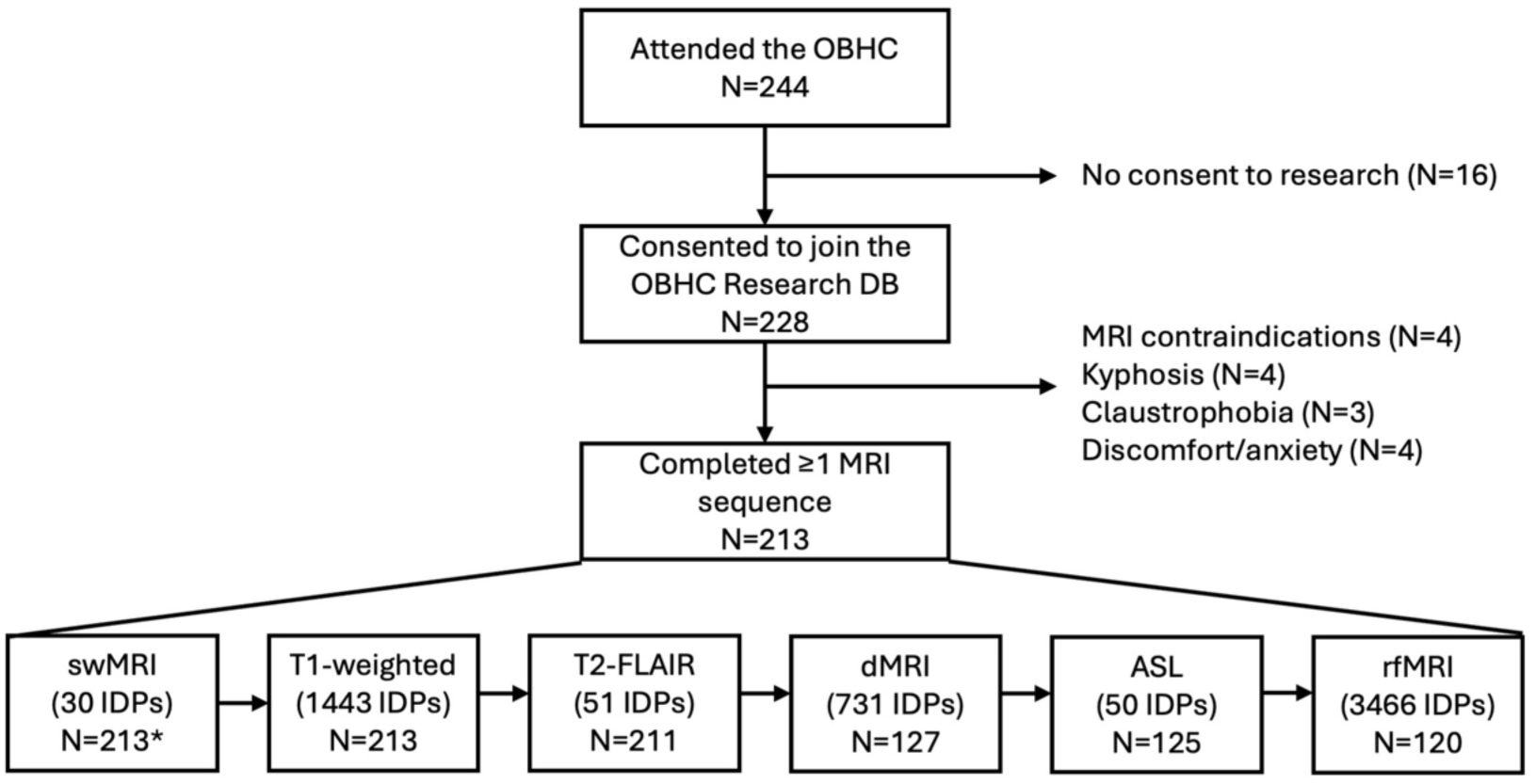
Available MRI data with the number of IDPs generated from each modality. One scan was unsuccessfully archived from the scanner and excluded from subsequent analyses. * For 19 swMRI scans with incomplete data (uncombined channel images unavailable), the scanner-reconstructed swMRI images were used as inputs for the core swMRI pipeline, but the QSM pipeline was skipped since it relies on phase images prior to high-pass filtering. See Supplementary Figure 1 for the equivalence of the T2* IDPs generated from the uncombined and scanner-combined images.

### 3.3 Quality Control

Enhanced QC of the T1-weighted, T2-FLAIR, dMRI, and rfMRI scans flagged 14-24% of scans for further inspection (Table 3). See Supplementary Table 2 for counts of IQMs flagged for each modality. Visual inspection of core outputs from T1, T2-FLAIR, and dMRI revealed that 0-2.4% of segmentations were low quality and subsequently excluded. Visual inspection of DSE plots revealed that although residual noise was high (S-var > 75% at timepoints) in 97.1% (33/34) of processed rfMRI data, no scans were deemed unusable based on this criterion. See Supplementary Table 3 for full visual QC results.

**Table 3:** Quality control results.

| Modality | % flagged (N/total) | % excluded (N/total) |
| --- | --- | --- |
| T1 | 14.6% (31/212) | 1.4% (3/212) all IDPs<br>2.4% (5/212) one or more IDPs |
| T2-FLAIR | 16.7% (35/210) | 1.0% (2/210) |
| dMRI | 19.0% (24/126) | 0% (0/126) |
| rfMRI | Pre-FIX: 24.2% (29/120)<br>Post-FIX: 10.0% (12/120) | 0% (0/120) |

### 3.4 Associations with age

Using covariates of sex, head size, and total cognitive score (ACE-III), IDPs from 5 out of the 6 MRI modalities were significantly associated with age (Figure 4-Figure 5). The strongest negative associations were observed with volumetric measures derived from the T1-weighted scans (p=240), with the strongest associations being with the total grey matter volume (SIENAX) and some cerebellar volumes (VBM). Periventricular, deep, and total white matter hyperintensity (WMH) volumes all positively associated with age. Mean T2* in the right amygdala was negatively associated with age, while no other swMRI-derived metrics survived correction for multiple testing. The fractional anisotropy (FA) of 14 tracts negatively associated with age, while mean diffusivity (MD) from 24 tracts positively associated with age. The NODDI-derived metrics ICVF and ISOVF negatively and positively associated with age, respectively, in many tracts. 32 ASL-derived measures associated with age (negative associations with cerebral blood flow and positive associations with arrival time). No rfMRI-derived IDPs were significantly associated with age. A similar pattern was found across all IDPs when not covarying for total cognitive score (Supplementary Figures 2-3).

**Figure 4:**
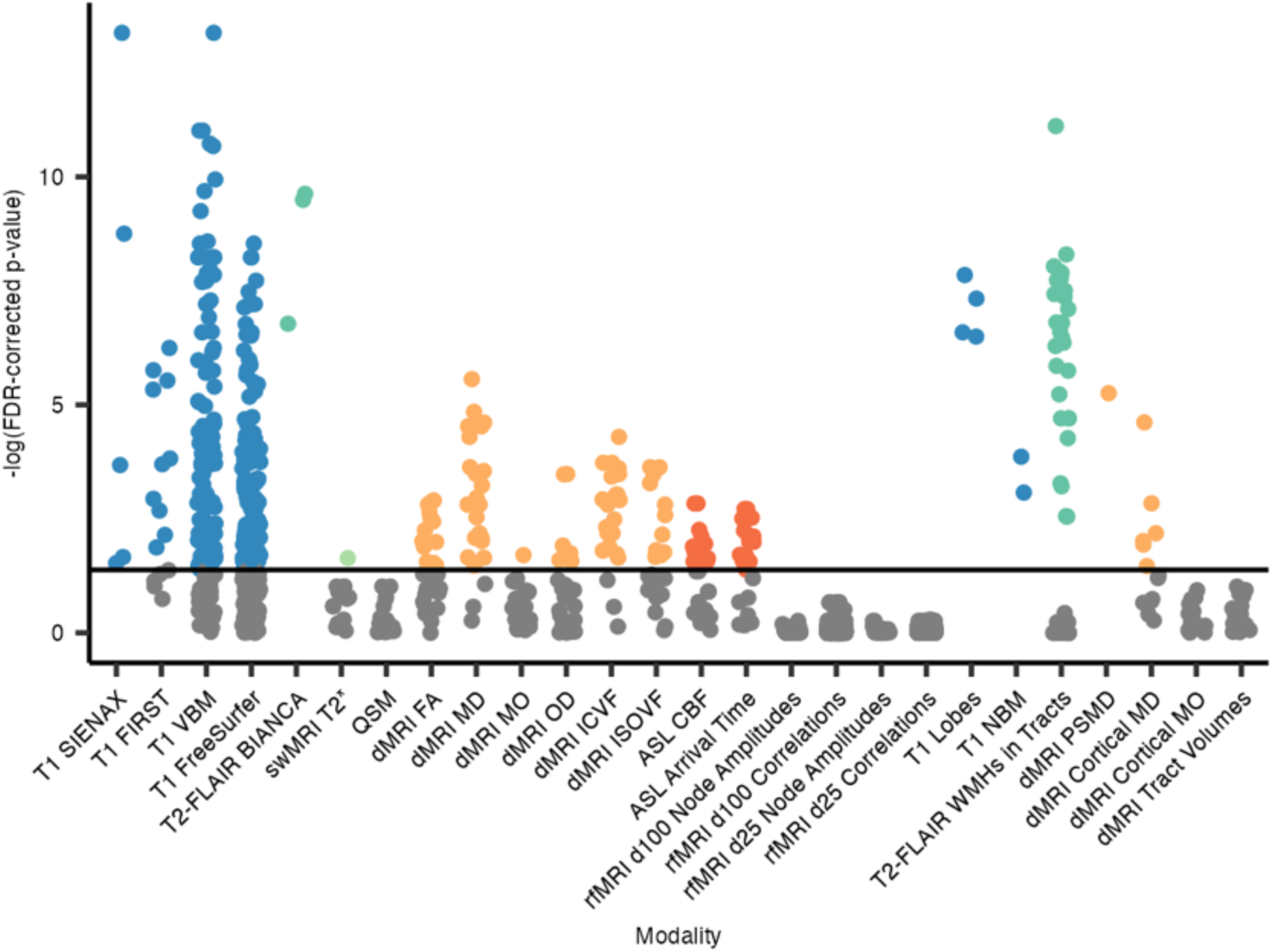
FDR-corrected p-values, hierarchical by modality, for associations between IDPs and age. Each dot represents one IDP, grouped by analysis tool/method and colour-coded by scan modality if significant. SIENAX, Structural Image Evaluation using Normalization of Atrophy (cross-sectionally); FIRST, FMRIB’s Integrated Registration and Segmentation Tool; VBM, voxel-based morphometry; BIANCA, Brain Intensity AbNormality Classification Algorithm; FA, fractional anisotropy; MD, mean diffusivity; MO, mode of anisotropy; OD, orientation dispersion index; ICVF, intra-cellular volume fraction; ISOVF, isotropic volume fraction; CBF, cerebral blood flow; NBM, nucleus basalis of Meynert; WMH, white matter hyperintensity; PSMD, peak width of skeletonised mean diffusivity.

**Figure 5:**
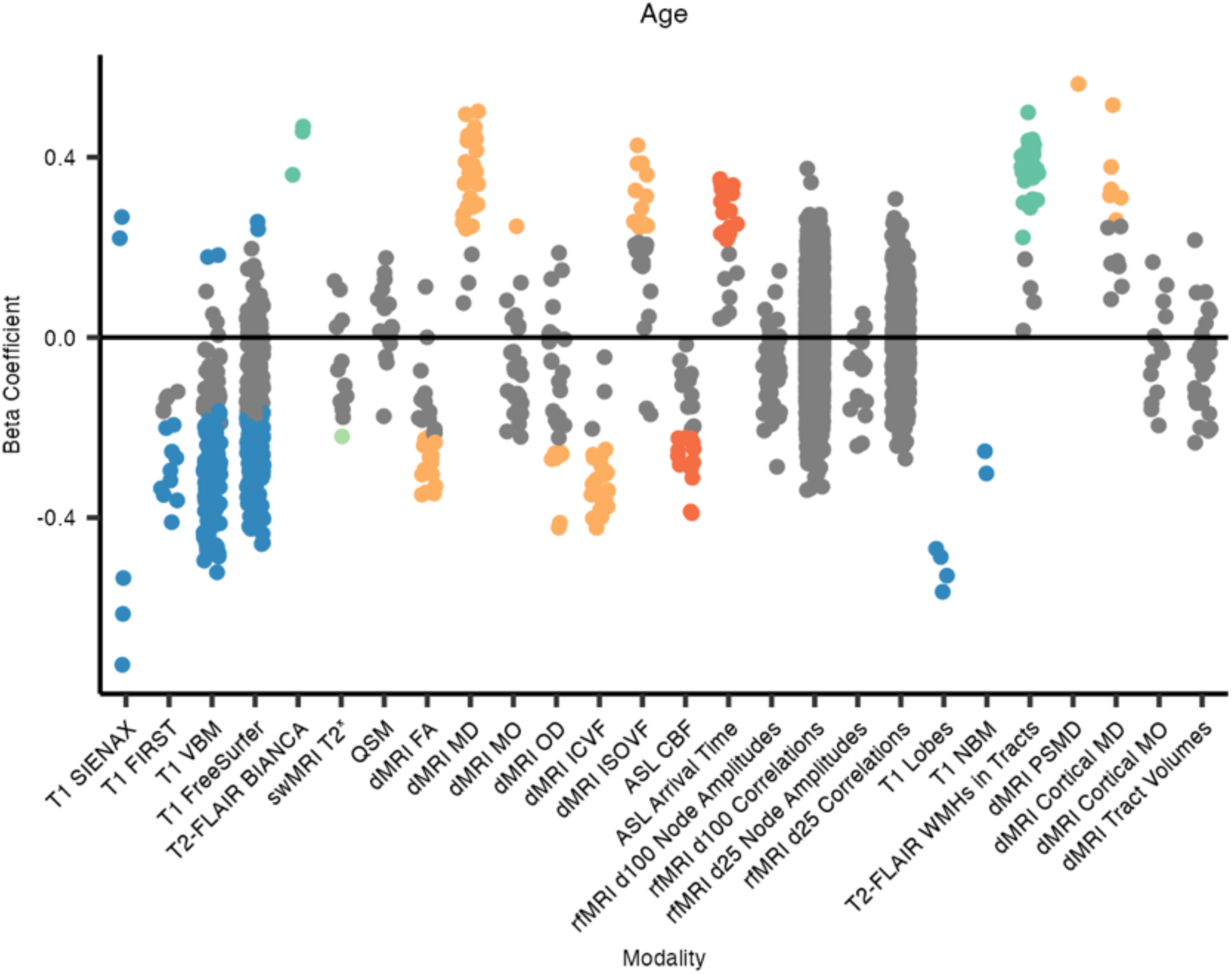
Beta coefficients for associations between IDPs and age. All numeric variables are unit standardised, meaning that 1 standard deviation (SD) increase in an IDP value is associated with a β SD difference in age. Coloured dots indicate associations significant at the 5% FDR level, hierarchical by modality. SIENAX, Structural Image Evaluation using Normalization of Atrophy (cross-sectionally); FIRST, FMRIB’s Integrated Registration and Segmentation Tool; VBM, voxel-based morphometry; BIANCA, Brain Intensity AbNormality Classification Algorithm; FA, fractional anisotropy; MD, mean diffusivity; MO, mode of anisotropy; OD, orientation dispersion index; ICVF, intra-cellular volume fraction; ISOVF, isotropic volume fraction; CBF, cerebral blood flow; NBM, nucleus basalis of Meynert; WMH, white matter hyperintensity; PSMD, peak width of skeletonised mean diffusivity.

Regarding the additional dementia-informed IDPs, all 4 lobar grey matter (GM) volumes negatively associated with age, with the strongest being with the temporal GM volume. Volumes of the bilateral nuclei basalis of Meynert also negatively associated with age. WMH volumes within 30 tracts were positively associated with age, with the most significant being the WMH volume in the right superior corona radiata. The peak width of the skeletonised MD (PSMD) positively associated with age, as did 6 cortical MD IDPs. See Supplementary Table 4 for the full list of associations that were significant following multiple testing correction.

When controlling for ROI/tract volumes, no swMRI-derived IDPs and fewer tractography-based IDPs survived hierarchical FDR correction (Supplementary Figures 4-5).

### 3.5 Associations with cognition

IDPs from 4 out of the 6 MRI modalities were significantly associated with ACE-III total scores, after controlling for age, sex, and head size (Figure 6-Figure 7). Regarding the core UK Biobank-aligned IDPs, most of the significant associations were with volumetric measures derived from the T1-weighted scans (p=160), with the strongest positive associations being with volumes of the posterior left middle temporal gyrus (VBM) and the peripheral GM (SIENAX). Periventricular and total white matter hyperintensity (WMH) volumes negatively associated with ACE-III total scores. Of the core dMRI-derived IDPs, most significant ones relate to the parahippocampal cinguli. Mean diffusivity (MD) metrics in the parahippocampal cinguli were most strongly negatively associated with cognition, while most other significant associations, such as with FA, MO, and ICVF, are positive. 11 node amplitude IDPs from rfMRI were positively associated with cognition, primarily relating to the salience, frontoparietal, and language networks.

**Figure 6:**
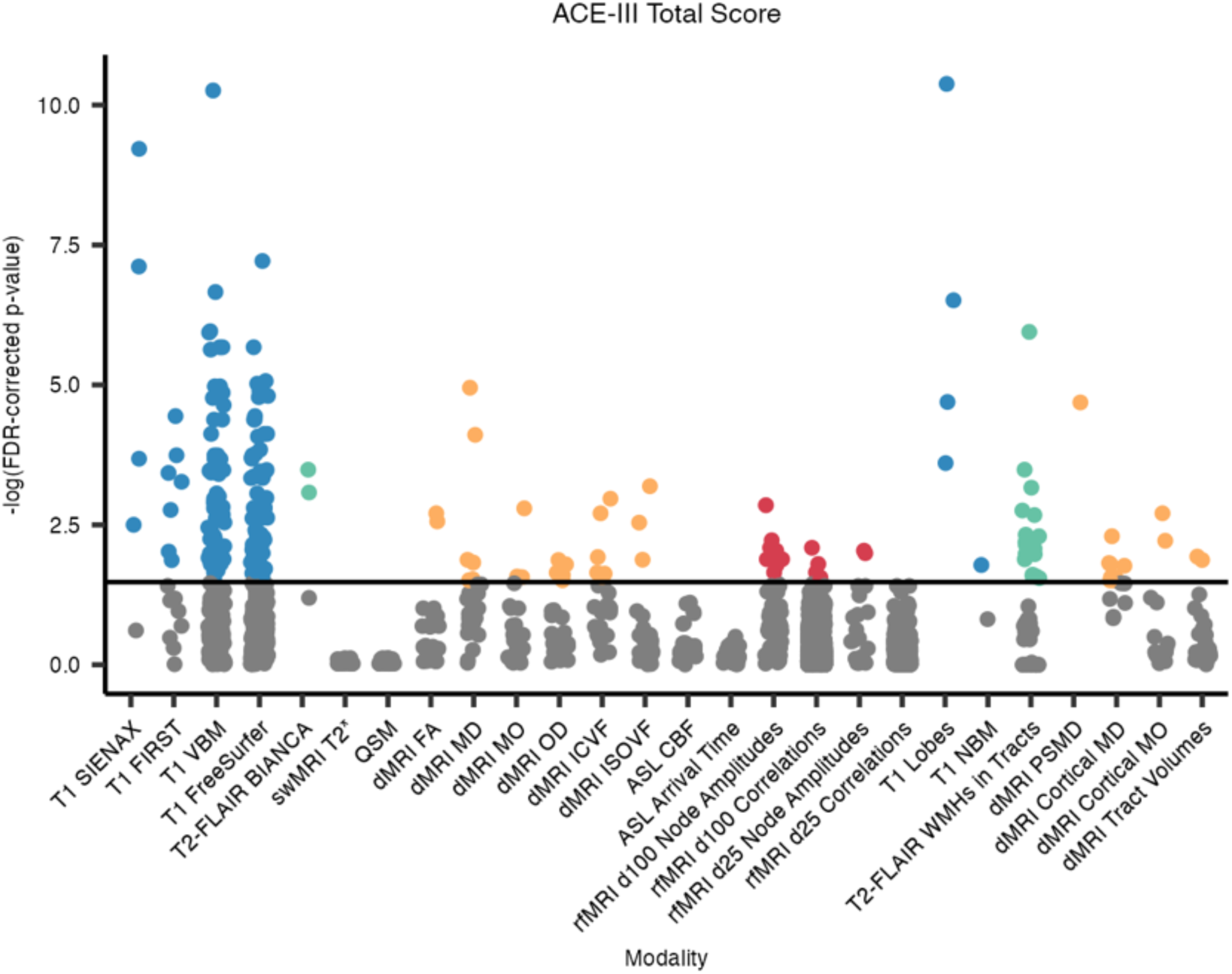
FDR-corrected p-values, hierarchical by modality, for associations between IDPs and ACE-III total cognitive scores. Each dot represents one IDP, grouped by analysis tool/method and colour-coded by scan modality if significant. SIENAX, Structural Image Evaluation using Normalization of Atrophy (cross-sectionally); FIRST, FMRIB’s Integrated Registration and Segmentation Tool; VBM, voxel-based morphometry; BIANCA, Brain Intensity AbNormality Classification Algorithm; FA, fractional anisotropy; MD, mean diffusivity; MO, mode of anisotropy; OD, orientation dispersion index; ICVF, intra-cellular volume fraction; ISOVF, isotropic volume fraction; CBF, cerebral blood flow; NBM, nucleus basalis of Meynert; WMH, white matter hyperintensity; PSMD, peak width of skeletonised mean diffusivity.

**Figure 7:**
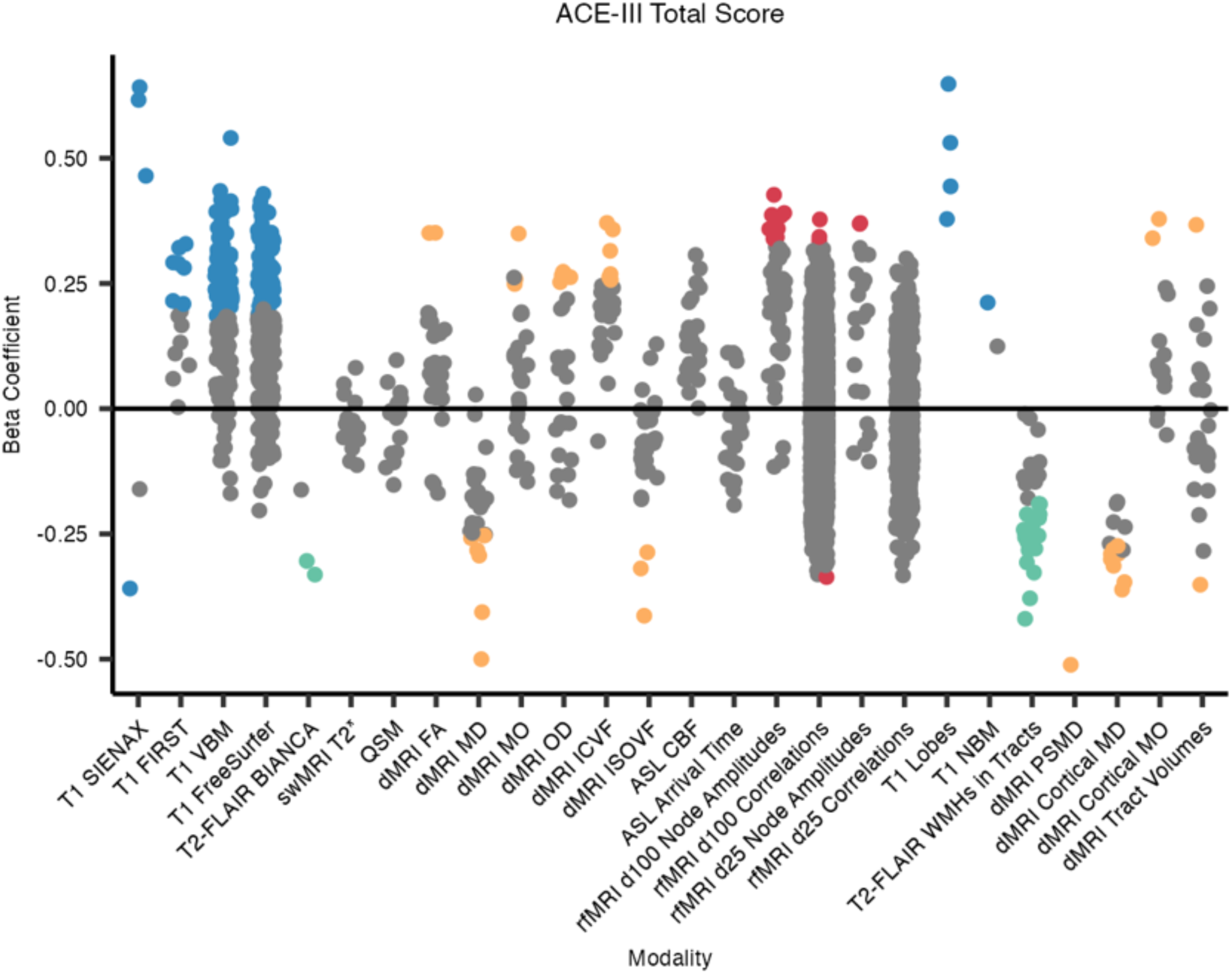
Beta coefficients for associations between IDPs and ACE-III total cognitive scores. All numeric variables are unit standardised, meaning that 1 standard deviation (SD) increase in an IDP value is associated with a β SD difference in ACE-III total cognitive score. Coloured dots indicate associations significant at the 5% FDR level, hierarchical by modality. SIENAX, Structural Image Evaluation using Normalization of Atrophy (cross-sectionally); FIRST, FMRIB’s Integrated Registration and Segmentation Tool; VBM, voxel-based morphometry; BIANCA, Brain Intensity AbNormality Classification Algorithm; FA, fractional anisotropy; MD, mean diffusivity; MO, mode of anisotropy; OD, orientation dispersion index; ICVF, intra-cellular volume fraction; ISOVF, isotropic volume fraction; CBF, cerebral blood flow; NBM, nucleus basalis of Meynert; WMH, white matter hyperintensity; PSMD, peak width of skeletonised mean diffusivity.

Regarding the additional dementia-informed IDPs, all 4 lobar grey matter (GM) volumes positively associated with ACE-III, with the strongest being with the temporal GM volume. WMH volumes within 20 tracts were negatively associated with ACE-III, with the most significant being the WMH volume in the splenium of the corpus collosum. PSMD negatively associated with cognition and was the second-most significant dMRI-derived metric overall. Cortical MD metrics, including in the bilateral supramarginal gyri, hippocampi, amygdalae, and parahippocampal cinguli, were negatively associated with cognitive scores, while MO in the bilateral supramarginal gyri positively associated with cognition. See Supplementary Table 5 for the full list of significant associations.

When controlling for ROI/tract volumes, no swMRI-derived IDPs and fewer tractography-based IDPs survived hierarchical FDR correction (Supplementary Figures 6-7).

### 3.6 Associations with diagnoses

A similar pattern of associations is observed between IDPs and diagnostic groups (dementia, MCI, no dementia-related diagnosis; Figure 8-Figure 9). This is likely because the results of cognitive assessment play an important role in dementia diagnosis. See Supplementary Table 6 for the full list of significant associations.

**Figure 8:**
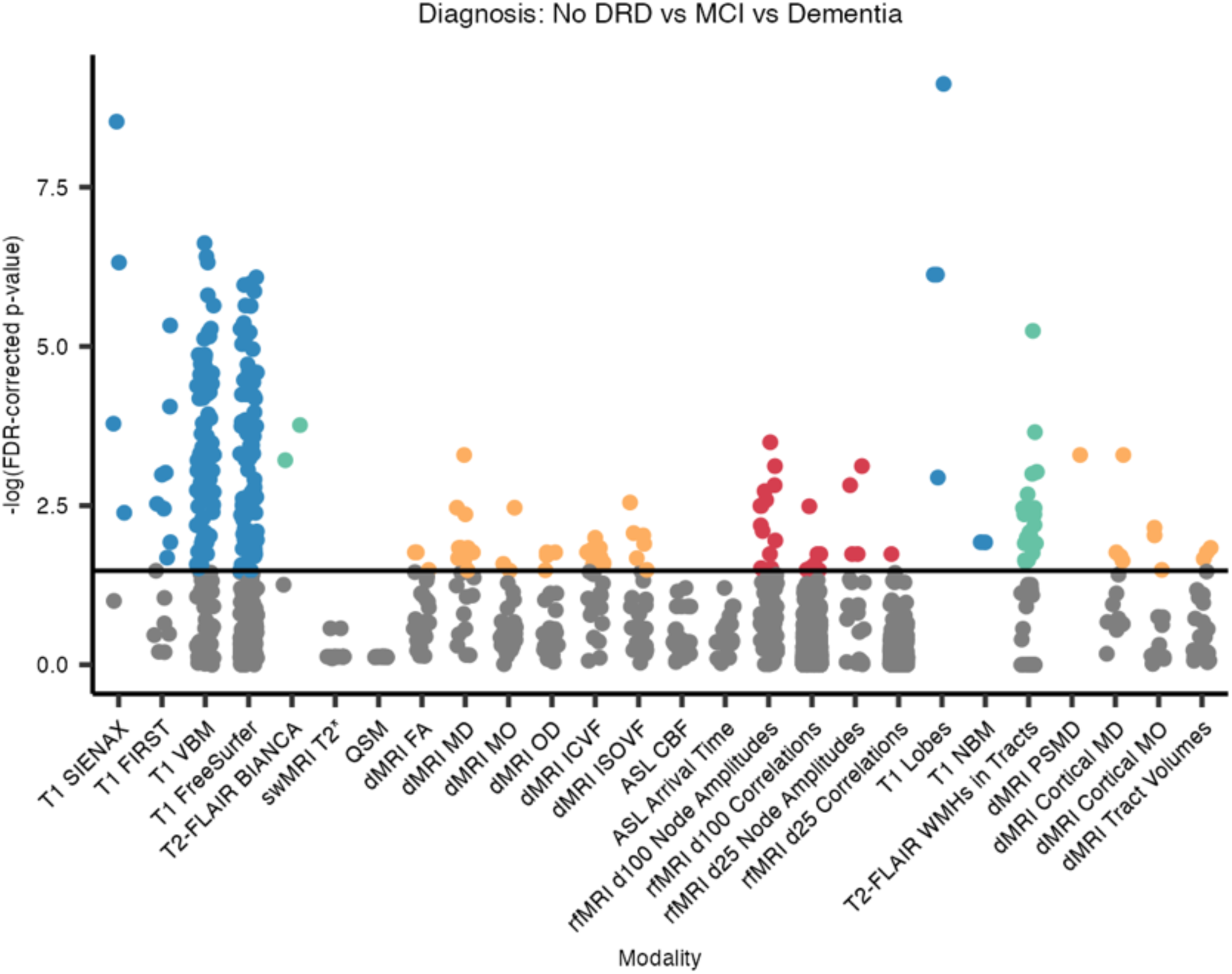
FDR-corrected p-values, hierarchical by modality, for associations between IDPs and diagnostic groups. Each dot represents one IDP, grouped by analysis tool/method and colour-coded by scan modality if significant. DRD, dementia-related diagnosis; SIENAX, Structural Image Evaluation using Normalization of Atrophy (cross-sectionally); FIRST, FMRIB’s Integrated Registration and Segmentation Tool; VBM, voxel-based morphometry; BIANCA, Brain Intensity AbNormality Classification Algorithm; FA, fractional anisotropy; MD, mean diffusivity; MO, mode of anisotropy; OD, orientation dispersion index; ICVF, intra-cellular volume fraction; ISOVF, isotropic volume fraction; CBF, cerebral blood flow; NBM, nucleus basalis of Meynert; WMH, white matter hyperintensity; PSMD, peak width of skeletonised mean diffusivity.

**Figure 9:**
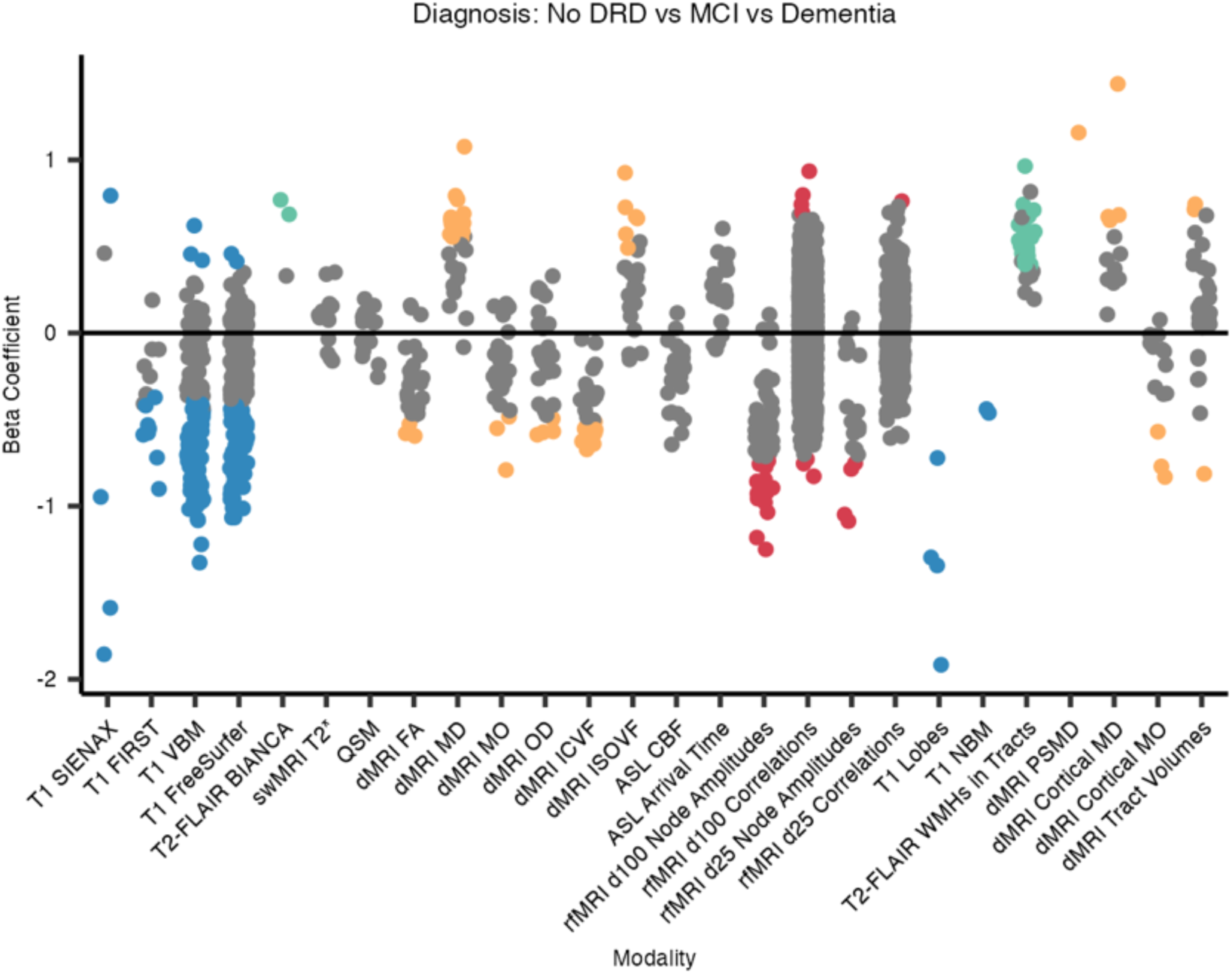
Beta coefficients for associations between IDPs and diagnostic groups. All numeric variables are unit standardised, meaning that for a 1 standard deviation (SD) increase in an IDP value, the log-odds of receiving a dementia-related diagnosis increases (or decreases if negative) by the β coefficient. Coloured dots indicate associations significant at the 5% FDR level, hierarchical by modality. DRD, dementia-related diagnosis; SIENAX, Structural Image Evaluation using Normalization of Atrophy (cross-sectionally); FIRST, FMRIB’s Integrated Registration and Segmentation Tool; VBM, voxel-based morphometry; BIANCA, Brain Intensity AbNormality Classification Algorithm; FA, fractional anisotropy; MD, mean diffusivity; MO, mode of anisotropy; OD, orientation dispersion index; ICVF, intra-cellular volume fraction; ISOVF, isotropic volume fraction; CBF, cerebral blood flow; NBM, nucleus basalis of Meynert; WMH, white matter hyperintensity; PSMD, peak width of skeletonised mean diffusivity.

## 4 Discussion

In this study, we have optimised a well-established imaging framework for a typical NHS memory clinic population by integrating enhanced quality control processes and dementia-specific analyses. Our detailed QC revealed that the imaging protocol is feasible, acceptable, and yields high-quality data that is usable for both clinical and research purposes. We found that this enhanced pipeline can generate reliable UKB-aligned IDPs and dementia-informed supplementary IDPs. We replicated established associations from earlier work and found novel associations between brain metrics and age, cognition, and diagnosed dementia. Together, these findings validate the use of this enhanced methodology in this real-world population and demonstrate its potential to bridge the gap between big data studies and clinical settings.

The Oxford Brain Health Clinic (OBHC) model of integrated, tiered research opportunities (O’Donoghue et al., 2023) continues to yield high rates of consent for research participation (93.4%) and additional scanner time (67.6%). Completion rates likewise remain high, with 99.1% and 72.7% of consenting patients completing the clinical and research sequences, respectively. All but one of the 213 scans were usable for radiologist reporting and therefore provided usable, structured data to the memory clinic.

One strength of this study is the seamless integration of rigorous QC measures into the UKB pipeline, creating a comprehensive framework for analysing real-world datasets. Automated QC tools (MRIQC, QUAD, and DSE decomposition) were used to flag T1-weighted, T2-FLAIR, dMRI, and rfMRI scans for visual inspection. We performed detailed QC at the level of individual IQMs, inspecting any scans that were outliers for at least one of these metrics. This process revealed that most of the raw data and pipeline outputs were useable. Although we accept that our outlier flagging approach might be overly sensitive, the percentage of flagged and excluded scans or IDPs in our study (Table 3) is consistent with other studies including automated QC and visual inspection. For example, in a sample of 4282 scans by Alfaro-Almagro and colleagues (2018), 17.5% of T1-weighted scans were flagged for inspection and 1.7% were excluded. Hence, this work supports the clinical application of automated QC tools as part of a staged QC approach. In clinical settings, we cannot afford to blindly exclude data based solely on fixed criteria or metrics, and we have demonstrated that this approach is useful to reduce the burden of visual inspection.

The analysis pipeline generates IDPs that are aligned with UK Biobank, while being suitable and relevant to a memory clinic population. In addition to incorporating previously described modifications to account for the high degree of atrophy and vascular burden (Griffanti et al., 2022), we added additional IDPs to capture other previously reported dementia-related changes. Many of these additional analyses are computationally light, ensuring that they won’t significantly increase the computational load beyond the existing UKB pipeline. The code is openly available on Gitlab (https://git.fmrib.ox.ac.uk/open-science/analysis/brain-health-clinic-mri).

We were then able to validate our obtained IDPs by replicating associations with well-known brain health changes in this real-world memory clinic population, carefully controlling multiple testing within each modality with a hierarchical FDR procedure. As anticipated, associations with age, cognition, and diagnoses were mainly with volumetric IDPs, particularly corresponding to temporal lobe structures. Associations with periventricular and total WMH volumes were stronger than with deep WMH volumes, in keeping with research studies in older adults (Bolandzadeh et al., 2012; Griffanti et al., 2018). Some of the strongest dMRI-derived associations were with the bilateral parahippocampal cinguli, which have previously been widely implicated in dementia and cognitive decline (Bozzali et al., 2011; Hirschfeld et al., 2023). In addition to the tensor-based metrics (e.g., MD), NODDI-derived metrics (i.e., ICVF and ISOVF) also show significant associations with clinical phenotypes, consistent with the global findings of McCracken and colleagues (2022). From rfMRI, associations with node amplitudes were most consistent. Using the same node labels as Lee and colleagues (2023), left fronto-parietal and language node amplitudes were positively associated with cognition meanwhile higher node amplitudes in the default mode, salience, and attention networks were associated with lower odds of receiving a dementia-related diagnosis.

The dementia-informed IDPs may capture additional variability in this memory clinic population. Lobar grey matter volumes have well-established relevance to cognitive status and prognosis, with the smaller temporal lobe volumes particularly associated with cognitive decline and dementia (Harper et al., 2017; Rabinovici et al., 2007; Visser et al., 2002; Woodworth et al., 2022). We, too, found the strongest associations with the temporal lobe (age: β=-0.56, corrected p=1.42 x 10^-8^; ACE-III: β=0.65, corrected p=4.21 x 10^-11^; diagnosis: OR=0.147, corrected p=7.56 x 10^-10^), although all 4 lobes were significantly associated. In our analyses, many tract-specific WMH volumes associated with clinical phenotypes to a similar or greater extent than total WMH volume or even the periventricular-deep subclassification, supporting the literature on the relevance of WMH regional distributions (Biesbroek et al., 2016, 2024; Veldsman et al., 2020). The peak width of the skeletonised mean diffusivity (PSMD) was strongly associated with age, cognition and diagnoses, with effect sizes similar to those reported by Satizabal and colleagues (Satizabal et al., 2020). Our findings extend the evidence supporting the use of this simple summary measure of small vessel disease to a memory clinic setting (Baykara et al., 2016; Deary et al., 2019).

In keeping with the literature on cortical mean diffusivity (MD) (Douaud et al., 2011, 2022; Illán-Gala et al., 2019; Montal et al., 2021; Weston et al., 2020), we also observed associations with age (p=6), cognition (p=8), and diagnosis (p=4), supporting the potential of MD to detect microstructural cortical changes in a memory clinic population. Although cortical mode of anisotropy (MO) is less common, we observed significant associations with MO in the bilateral supramarginal gyri (cognition and diagnosis) and right amygdala (diagnosis), supporting the ability of this measure to also detect dementia-related changes. Some tract volumes were significantly associated with cognition (left corticospinal tract and major fornix) and diagnoses (left corticospinal tract, major fornix, and left medial lemniscus), but the directionality of these associations was inconsistent. Like many dMRI-derived measures, changes in tract volumes are less specific, being influenced by both atrophy and microstructural changes, but nevertheless they can highlight tract-specific atrophy patterns or flag poorer-quality reconstructions (Groot et al., 2015).

Consistent with the literature on the cholinergic systems in dementia (Bohnen et al., 2018; Lagarde et al., 2024; Schumacher et al., 2022), we found that volumes of the Nucleus basalis of Meynert (NBM) bilaterally correlated with dementia diagnosis and age. The left NBM volume also associated with cognition. These associations were not as strong as with many other volumetric IDPs, but they may nevertheless hold value for differential diagnosis and prognosis.

In this work, we present a pragmatic statistical approach to enable data-driven exploratory research in the clinical setting. Large sample sizes may be needed to reliably detect associations in the style of big data analyses (Marek et al., 2022), but without proper adjustments these sample sizes also inflate the risks of spurious associations. Indeed, moderate sample sizes may also be sufficient to detect associations with larger true effect sizes (Spisak et al., 2023). As opposed to big data studies which may include hundreds of covariates (Alfaro-Almagro et al., 2021), here we employ a core set of covariates more in line with clinical settings. We utilise hierarchical FDR-correction by first correcting for comparisons across MRI modalities and then within modality using the Benjamini-Hochberg procedure applied to those that survive the Simes test (Benjamini & Hochberg, 1995; Yekutieli, 2008). With hierarchical FDR control, we mitigate the risks associated with non-independent analyses in small sample sizes (Nichols & Poline, 2009; Vul et al., 2009; Yarkoni, 2009). This novel application in a memory clinic setting enables data-driven exploration of high-dimensional neuroimaging data without relying on the select few large cohort studies.

A number of methodological limitations need to be considered when interpreting this study. Although this enhanced pipeline includes integrated QC for some modalities (T1, T2-FLAIR, dMRI, and rfMRI), more work is needed to develop appropriate QC metrics and tools for swMRI, QSM, and ASL. For the modalities with integrated QC, it is important to note that the IQMs are no-reference measures (i.e., without a ground truth). Flagging outliers was used as common criterion across IQMs across modalities, but this may not be necessarily the best method to detect quality deviations. Automated classifiers using IQMs as features do exist, but they are mostly for T1-weighted scans, and classification accuracy varies substantially on clinical datasets (Bhalerao et al., 2024). When using traditional cut-offs for DSE plots, our rfMRI QC reveals that some further optimisation may be required for this memory clinic population, but it remains unclear whether different thresholds may be more appropriate for this population, as no dataset was deemed unusable after further visual inspection. This work concentrated on volumetric analyses, but additional automated QC (e.g., Qoala-T - Klapwijk et al., 2019) and optimisation of Freesurfer surface-based outputs may also be warranted, especially in this clinical population with substantial WMHs (Oi et al., 2023).

In addition, although our selection of dementia-informed IDPs is non-exhaustive, they demonstrate that the UKB IDPs may underrepresent the full picture when there is a clear clinical question. Compared to big data studies, the sample size here affects our ability to accurately detect associations. However, in this study we demonstrate the utility of using hierarchical FDR for detecting both known and novel associations in the presence of limited data. Future work could expand on the set of essential covariates used here and include analyses with greater sensitivity to non-linear associations. Nevertheless, the findings here serve to demonstrate the possibilities enabled by integrating UK Biobank-aligned imaging and analyses, enriched for sensitivity to dementia-related changes, directly into clinical settings. Further work will aim to address the extent to which these IDPs capture unique variability and inform differential diagnosis and precision phenotyping in a memory clinic setting.

This enriched and integrated quality control-analysis pipeline for memory clinics offers possibilities in both clinical and research spheres. In clinical practice, the combination of established and novel quantitative measures has the potential to significantly improve the accuracy of differential diagnoses and predicted responses to treatment. In contrast to UK Biobank where participants are mostly healthy, this study better describes the brain alterations associated with dementia in a sample of memory clinic attendees. Moreover, because the metrics here are aligned with UK Biobank, any important advancements can be easily re-integrated with the larger field of brain imaging research, thereby facilitating further work in personalised medicine, normative modelling. Wider use of this analysis-QC pipeline in memory clinic settings also enables unique research opportunities in real-world clinical populations. We currently present a rich dataset of deeply-phenotyped, unselected memory clinic patients, but this dementia-enhanced analysis pipeline is also well-suited to wider use and offers a framework to plug-in and pilot additional analyses suited to similar clinical applications.

## 5 Data and code availability

The complete OBHC MRI protocol and scanning procedure is available through the WIN MR Protocols Database at: https://open.win.ox.ac.uk/protocols/stable/6974395a-3745-4861-b8cc-1887e787d1c4 (O’Donoghue et al., 2022b).

The UKB-dementia pipeline presented here and used for this analysis is openly available (https://git.fmrib.ox.ac.uk/open-science/analysis/brain-health-clinic-mri) along with the original UK Biobank brain MRI analysis pipeline (https://git.fmrib.ox.ac.uk/falmagro/uk_biobank_pipeline_v_1.5/-/tree/master).

Additional scripts used for OBHC analyses are also available (https://git.fmrib.ox.ac.uk/gillisc/01_bhc_imaging.git). Interactive versions of the figures are available (https://users.ox.ac.uk/~scat8503/).

The MRI data presented in this paper will be available via the Dementias Platform UK (https://portal.dementiasplatform.uk/CohortDirectory/Item?fingerPrintID=BHC), and access will be granted through an application process, reviewed by the OBHC Data Access Group. The OBHC Data Access Group will start accepting applications to access OBHC data upon publication of the present work. Data will continue to be released in batches as the OBHC progresses in order to minimise the risk of participant identification.

## 6 Author contributions

**Grace Gillis:** Conceptualization, Data curation, Formal analysis, Investigation, Methodology, Software, Validation, Visualisation, Writing – original draft. **Gaurav Bhalerao:** Methodology, Software, Validation, Writing – review & editing. **Jasmine Blane:** Investigation, Project administration, Writing – review & editing. **Robert Mitchell:** Investigation, Writing – review & editing. **Pieter M Pretorius:** Investigation, Writing – review & editing. **Celeste McCracken:** Methodology, Writing – review & editing. **Thomas E Nichols:** Methodology, Writing – review & editing. **Steve M Smith:** Methodology, Software, Funding acquisition, Writing – review & editing. **Karla L. Miller:** Methodology, Software, Funding acquisition, Writing – review & editing. **Fidel Alfaro-Almagro:** Methodology, Software, Writing – review & editing. **Vanessa Raymont:** Conceptualization, Resources, Funding acquisition, Supervision, Writing – review & editing. **Lola Martos:** Conceptualization, Resources, Funding acquisition, Writing – review & editing. **Clare E Mackay:** Conceptualization, Methodology, Resources, Funding acquisition, Supervision, Writing – review & editing. **Ludovica Griffanti:** Conceptualization, Methodology, Resources, Funding acquisition, Supervision, Writing – review & editing.

## Acknowledgements

We are grateful to the operations team of the OBHC (see https://www.psych.ox.ac.uk/research/translational-neuroimaging-group/team/oxford-brain-health-clinic), Christine Kindler, Gwenaëlle Douaud, and Benjamin Tendler.

## 9 Supplementary Material

The following are the supplementary material to this article: [See Gillis_UKB-Dementia_Supp.pdf].

