## Supplemental Materials for "From Big Data to the clinic: methodological and statistical enhancements to implement the UK Biobank imaging framework in a memory clinic" (from another version of this preprint: version 1, 2024-08-04)

### Supplementary Materials

#### Supplementary Table 1: Protocol

| **Modality** | **Acqui-sition time** | **Resolution** | **Matrix** | **Key Parameters** | **UKB Match** |
|---|---|---|---|---|---|
| **BHC core clinical protocol** | 16:29 | | | | |
| Localiser | 0:14 | | | | No |
| Diffusion MRI (dMRI 3-scan trace) | 0:43 | 0.8x0.8x4 mm | 260x260x27 | TR = 3800 ms, 3 dirs, b = 0, 1000 s/mm$^2$ | N/A |
| Susceptibility-weighted MRI (swMRI) | 4:46 | 0.8x0.8x1.5 mm | 256x288x80 | TE1/TE2/TR = 10/20/30 ms, R = 2 | Adapted |
| T1 (MPRAGE) | 4:54 | 1.0x1.0x1.0 mm | 256x256x208 | TI/TR = 800/2000 ms, R = 2 | Exact |
| T2-FLAIR (SPACE) | 5:52 | 1.0x1.0x1.05 mm | 256x256x192 | TI/TR = 1800/5000 ms, R = 2 | Exact |
| **BHC research protocol** | 21:17 | | | | |
| ASL localiser (time of flight) | 0:42 | | | | N/A |
| Diffusion MRI (dMRI) | 7:08 | 2.0x2.0x2.0 mm | 104x104x72 | TR = 3600 ms, 50 dirs/shell, b = 0,1000,2000 s/mm2, MB = 3, blip-reversed b = 0 | Exact |
| Arterial spin labelling (PCASL) | 7:17 | 3.4x3.4x4.5 mm | 64x64x24 | TE/TR = 14/4400 ms, tag = 1400 ms, seven PLDs = 250,500…1750 ms, 1 M0 calibration image | Not in UKB initial protocol |
| Resting state fMRI (rfMRI) | 6:10 | 2.4x2.4x2.4 mm | 88x88x64 | TE/TR = 39/735 ms, α = 52°, MB = 8 | Exact |

MPRAGE = Magnetization Prepared RApid Gradient Echo; FLAIR = Fluid-attenuated inversion recovery; SPACE = Sampling Perfection with Application optimized Contrasts using different flip angle Evolution; ASL = Arterial Spin Labelling; PCASL = pseudo-continuous ASL; TR = Repetition time; TE = Echo time; TI = Inversion time; R = In-plane acceleration factor; MB = Multi-band acceleration factor; α = flip angle; PLD = postlabelling delay; M0 = equilibrium magnetization, required for ASL quantification. Full MRI protocols are available online (BHC: https://open.win.ox.ac.uk/protocols/stable/6974395a-3745–4861-b8cc-1887e787d1c4 (O'Donoghue *et al.*, 2022));
UKB: https://open.win.ox.ac.uk/protocols/stable/d2b297c3-4a4f-4fde-9b7c-7a8ae6e5fa83).

#### Supplementary Table 2: IQMs

Image quality metrics (IQMs) flagged by automated quality control. N is the number of scans flagged as outliers (>1.5 IQR away from Q1/Q3) for each IQM. DSE decomposition, which was run separately on the raw and processed rfMRI data, returns the whole (or total) D-var, S-var, and E-var sum-of-squares decomposition as well the global components within each of these. For details about the individual IQMs, please refer to Esteban et al. (2017) for MRIQC, Afyouni and Nichols (2018) for DSE and Bastiani et al. (2019) for EDDY QUAD.

| Modality | Flagged Image Quality Metrics (IQMs) |
|---|---|
| **T1 (MRIQC)** | CJV (N=1); CNR (N=1); SNR_CSF (N=0); SNR_GM (N= 5); SNR_WM (N=5); QI2 (N=7); EFC (N=2); FBER (N=5); INU_med (N=1); INU_range (N=1); WM2MAX (N=4) |
| **T2-FLAIR (MRIQC)** | CNR (N=6); SNR_CSF (N=1); SNR_GM (N=3); QI2 (N=3); EFC (N=3); FBER (N=3); INU_med (N=3); WM2MAX (N=18) |
| **dMRI (EDDY QUAD)** | SNR (N=3); CNR_b1000 (N=0); CNR_b1997 (N=1); absolute motion (N=8); relative motion (N=6); susceptibility (N=3); total % outliers (N=11) |
| **rfMRI (MRIQC, DSE decomposition)** | EFC (N=0); FBER (N=1); SNR (N=2); tSNR (N=2); GSR_x (N=7); GSR_y (N=0); GCOR (N=10); AOR (N=6); AQI (N=4); raw data % d-Var (whole – N=0; global – N=8); raw data % s-Var (whole – N=0; global – N=7); processed data % d-Var (whole – N=1; global – N=8); processed data % s-Var (whole – N=1; global – N=10) |

#### Supplementary Table 3: Visual QC

Detailed results of visual inspection of flagged scans. One patient dataset, flagged for T1-weighted and T2-FLAIR, could not be processed or visually inspected due to a scan archive failure (excluded from subsequent analyses).

| Modality (N inspected) | Output visually inspected | Quality | | |
|---|---|---|---|---|
| | | High (N) | Medium (N) | Low (N) |
| **T1-weighted (31)** | *Corrected SIENAX segmentation* | 90.0% (27) | 3.3% (1) | 6.7% (2) |
| | *Corrected FIRST segmentation* | 73.3% (22) | 3.3% (1) | 23.3% (7) |
| **T2-FLAIR (35)** | *BIANCA segmentation* | 70.6% (24) | 26.5% (9) | 2.9% (1) |
| **dMRI (24)** | *Tractography* | 70.8% (17) | 29.2% (7) | 0% (0) |
| | | **S-var exceeds 75%** | | |
| | | **Never (N)** | **Rarely (N)** | **Often (N)** |
| **rfMRI (34)** | *DSE plots* | 2.9% (1) | 29.4% (10) | 67.6% (23) |

#### Supplementary Table 4: Associations with age

Associations between IDPs and age after adjusting for ACE-III total cognitive score. Corr-pval are after hierarchical FDR correction.

| Variable | coef | pval | corr-pval |
|---|---|---|---|
| T1_SIENAX_GM_unnorm_vol_LM | -0.727 | 3.51E-16 | 6.93E-14 |
| T1_GM_parcellation_L_VI_Cerebellum_vol_LM | -0.521 | 2.73E-16 | 6.93E-14 |
| T2_FLAIR_WMHvol_Superior_corona_radiata_R | 0.499 | 1.51E-13 | 7.69E-12 |
| T1_GM_parcellation_R_VI_Cerebellum_vol_LM | -0.481 | 7.34E-14 | 9.67E-12 |
| T1_GM_parcellation_R_Crus_I_Cerebellum_vol_LM | -0.479 | 9.80E-14 | 9.68E-12 |
| T1_GM_parcellation_R_Crus_II_Cerebellum_vol_LM | -0.472 | 2.38E-13 | 1.88E-11 |
| T1_GM_parcellation_L_Crus_I_Cerebellum_vol_LM | -0.470 | 3.20E-13 | 2.11E-11 |
| T1_GM_parcellation_R_Amygdala_vol_LM | -0.495 | 2.02E-12 | 1.14E-10 |
| T1_GM_parcellation_R_Hippocampus_vol_LM | -0.480 | 4.16E-12 | 2.05E-10 |
| WMH_pvent | 0.468 | 9.23E-12 | 2.35E-10 |
| WMH_total | 0.457 | 1.88E-11 | 3.19E-10 |
| T1_GM_parcellation_L_Crus_II_Cerebellum_vol_LM | -0.441 | 1.28E-11 | 5.64E-10 |
| T1_SIENAX_peripheral_GM_unnorm_vol_LM | -0.614 | 4.45E-11 | 1.76E-09 |
| T1_GM_parcellation_R_V_Cerebellum_vol_LM | -0.437 | 7.26E-11 | 2.61E-09 |
| FS_HippSubfield_rh_volume_Whole.FS_Hippocampal.body. | -0.459 | 9.57E-11 | 2.91E-09 |
| T1_GM_parcellation_R_Parahippocampal_Gyr_ant_vol_LM | -0.459 | 8.91E-11 | 2.91E-09 |
| T2_FLAIR_WMHvol_Body_of_corpus_callosum | 0.430 | 3.94E-10 | 5.03E-09 |
| FS_HippSubfield_rh_volume_fimbria. | -0.420 | 2.57E-10 | 5.82E-09 |
| T1_GM_parcellation_R_Caudate_vol_LM | -0.486 | 2.20E-10 | 5.82E-09 |
| T1_GM_parcellation_L_Amygdala_vol_LM | -0.465 | 2.30E-10 | 5.82E-09 |
| T1_GM_parcellation_R_VIIb_Cerebellum_vol_LM | -0.413 | 2.65E-10 | 5.82E-09 |
| T1_GM_parcellation_R_VIIIa_Cerebellum_vol_LM | -0.415 | 2.38E-10 | 5.82E-09 |
| FS_HippSubfield_rh_volume_GC.ML.DG.body. | -0.456 | 2.82E-10 | 5.86E-09 |
| T1_GM_parcellation_L_VIIIa_Cerebellum_vol_LM | -0.411 | 3.02E-10 | 5.97E-09 |
| T2_FLAIR_WMHvol_Anterior_corona_radiata_R | 0.420 | 8.97E-10 | 9.14E-09 |
| T1_GM_parcellation_R_I.IV_Cerebellum_vol_LM | -0.407 | 5.77E-10 | 1.08E-08 |
| T2_FLAIR_WMHvol_Splenium_of_corpus_callosum | 0.439 | 1.36E-09 | 1.16E-08 |
| T2_FLAIR_WMHvol_Anterior_limb_of_internal_capsule_R | 0.437 | 1.74E-09 | 1.27E-08 |
| T1_GM_parcellation_R_Vent_Striatum_vol_LM | -0.418 | 7.28E-10 | 1.31E-08 |
| T1_GM_parcellation_R_Temp_Fusiform_ant_vol_LM | -0.434 | 8.22E-10 | 1.41E-08 |
| T1_GMvol_temp | -0.565 | 8.63E-10 | 1.42E-08 |
| T2_FLAIR_WMHvol_Posterior_thalamic_radiation_R | 0.407 | 2.69E-09 | 1.71E-08 |
| T2_FLAIR_WMHvol_Superior_corona_radiata_L | 0.407 | 3.27E-09 | 1.85E-08 |
| FS_HippSubfield_rh_volume_subiculum.body. | -0.436 | 1.20E-09 | 1.89E-08 |

| | | | |
|---|---|---|---|
| T1_GM_parcellation_L_VIIb_Cerebellum_vol_LM | -0.399 | 1.26E-09 | 1.91E-08 |
| T1_GM_parcellation_Vermis_VIIIa_Cerebellum_vol_LM | -0.404 | 1.36E-09 | 2.00E-08 |
| T1_GM_parcellation_R_Planum_Polare_vol_LM | -0.460 | 1.43E-09 | 2.01E-08 |
| T2_FLAIR_WMHvol_Superior_fronto.occipital_fasciculus_R | 0.428 | 6.07E-09 | 3.09E-08 |
| T2_FLAIR_WMHvol_Posterior_corona_radiata_L | 0.391 | 6.76E-09 | 3.13E-08 |
| FS_HippSubfield_rh_volume_CA4.body. | -0.425 | 2.42E-09 | 3.29E-08 |
| T2_FLAIR_WMHvol_Anterior_limb_of_internal_capsule_L | 0.410 | 9.09E-09 | 3.72E-08 |
| T2_FLAIR_WMHvol_Posterior_thalamic_radiation_L | 0.404 | 9.49E-09 | 3.72E-08 |
| T2_FLAIR_WMHvol_Genu_of_corpus_callosum | 0.384 | 1.18E-08 | 4.31E-08 |
| T1_GMvol_par | -0.529 | 3.55E-09 | 4.68E-08 |
| T1_GM_parcellation_L_Parahippocampal_Gyr_ant_vol_LM | -0.444 | 4.03E-09 | 5.13E-08 |
| FS_HippSubfield_lh_volume_fimbria. | -0.396 | 5.31E-09 | 6.17E-08 |
| T1_GM_parcellation_R_Precentral_Gyr_vol_LM | -0.417 | 5.12E-09 | 6.17E-08 |
| T1_GM_parcellation_L_Hippocampus_vol_LM | -0.435 | 5.27E-09 | 6.17E-08 |
| FS_HippSubfield_rh_volume_Whole.FS_Hippocampus. | -0.420 | 5.73E-09 | 6.47E-08 |
| FS_aparc.DKTatlas_lh_thickness_precentral. | -0.388 | 6.65E-09 | 7.29E-08 |
| T2_FLAIR_WMHvol_Anterior_corona_radiata_L | 0.375 | 2.32E-08 | 7.89E-08 |
| T1_GM_parcellation_L_V_Cerebellum_vol_LM | -0.381 | 1.13E-08 | 1.21E-07 |
| T2_FLAIR_WMHvol_Superior_longitudinal_fasciculus_R | 0.374 | 4.88E-08 | 1.56E-07 |
| T2_FLAIR_WMHvol_Posterior_corona_radiata_R | 0.364 | 5.27E-08 | 1.58E-07 |
| WMH_deep | 0.361 | 5.86E-08 | 1.66E-07 |
| FS_HippSubfield_rh_volume_presubiculum.head. | -0.397 | 1.62E-08 | 1.68E-07 |
| T1_GM_parcellation_Vermis_VIIb_Cerebellum_vol_LM | -0.363 | 2.46E-08 | 2.49E-07 |
| T2_FLAIR_WMHvol_External_capsule_L | 0.379 | 9.37E-08 | 2.51E-07 |
| FS_aparc.DKTatlas_rh_volume_entorhinal. | -0.379 | 2.68E-08 | 2.57E-07 |
| T1_GM_parcellation_L_I.IV_Cerebellum_vol_LM | -0.362 | 2.63E-08 | 2.57E-07 |
| T1_GMvol_front | -0.487 | 2.74E-08 | 2.57E-07 |
| FS_HippSubfield_rh_volume_Whole.FS_Hippocampal.head. | -0.402 | 3.14E-08 | 2.89E-07 |
| FS_HippSubfield_lh_volume_GC.ML.DG.body. | -0.418 | 3.37E-08 | 3.03E-07 |
| T1_GMvol_occ | -0.469 | 3.62E-08 | 3.17E-07 |
| T2_FLAIR_WMHvol_Posterior_limb_of_internal_capsule_L | 0.421 | 1.32E-07 | 3.37E-07 |
| T2_FLAIR_WMHvol_Superior_fronto.occipital_fasciculus_L | 0.367 | 1.79E-07 | 4.34E-07 |
| T2_FLAIR_WMHvol_External_capsule_R | 0.380 | 2.21E-07 | 5.13E-07 |
| T1_GM_parcellation_L_Caudate_vol_LM | -0.431 | 6.72E-08 | 5.65E-07 |
| R_hipp_masked | -0.362 | 6.70E-08 | 5.65E-07 |
| T1_GM_parcellation_L_X_Cerebellum_vol_LM | -0.391 | 7.75E-08 | 6.38E-07 |
| FS_HippSubfield_lh_volume_CA4.body. | -0.403 | 7.98E-08 | 6.44E-07 |
| T1_GM_parcellation_R_VIIIb_Cerebellum_vol_LM | -0.353 | 9.05E-08 | 7.15E-07 |
| FS_aparc.DKTatlas_rh_thickness_precentral. | -0.354 | 1.29E-07 | 9.96E-07 |

| | | | |
|---|---|---|---|
| T1_GM_parcellation_R_Postcentral_Gyr_vol_LM | -0.371 | 1.39E-07 | 1.06E-06 |
| FS_HippSubfield_lh_volume_subiculum.body. | -0.399 | 1.46E-07 | 1.09E-06 |
| T1_GM_parcellation_R_Lingual_Gyr_vol_LM | -0.436 | 1.62E-07 | 1.19E-06 |
| FS_HippSubfield_lh_volume_Whole.FS_Hippocampal.body. | -0.395 | 1.88E-07 | 1.35E-06 |
| T2_FLAIR_WMHvol_Superior_longitudinal_fasciculus_L | 0.354 | 6.29E-07 | 1.40E-06 |
| IDP_T1_FIRST_right_thalamus_volume | -0.410 | 2.51E-07 | 1.74E-06 |
| FS_HippSubfield_rh_volume_subiculum.head. | -0.369 | 2.49E-07 | 1.74E-06 |
| T1_GM_parcellation_L_Precentral_Gyr_vol_LM | -0.372 | 2.60E-07 | 1.77E-06 |
| T2_FLAIR_WMHvol_Posterior_limb_of_internal_capsule_R | 0.403 | 8.39E-07 | 1.78E-06 |
| T1_GM_parcellation_L_Vent_Striatum_vol_LM | -0.358 | 2.96E-07 | 1.98E-06 |
| FS_aparc.DKTatlas_rh_thickness_entorhinal. | -0.348 | 3.39E-07 | 2.23E-06 |
| IDP_dMRI_ProbtrackX_MD_atr_l | 0.502 | 1.25E-08 | 2.72E-06 |
| IDP_T1_FIRST_left_accumbens_volume | -0.350 | 4.52E-07 | 2.93E-06 |
| FS_aparc.DKTatlas_lh_thickness_entorhinal. | -0.349 | 4.92E-07 | 3.14E-06 |
| FS_HippSubfield_rh_volume_CA3.head. | -0.350 | 5.63E-07 | 3.53E-06 |
| T1_GM_parcellation_R_Front_Orbital_vol_LM | -0.385 | 6.44E-07 | 3.97E-06 |
| IDP_T1_FIRST_right_accumbens_volume | -0.335 | 7.62E-07 | 4.63E-06 |
| FS_HippSubfield_rh_volume_CA1.head. | -0.354 | 8.47E-07 | 5.07E-06 |
| dMRI_PSMD | 0.563 | 5.09E-08 | 5.55E-06 |
| T2_FLAIR_WMHvol_Cingulum_cingulate_gyrus_R | 0.407 | 2.88E-06 | 5.87E-06 |
| FS_HippSubfield_lh_volume_Whole.FS_Hippocampus. | -0.373 | 1.12E-06 | 6.63E-06 |
| T1_GM_parcellation_R_Parahippocampal_Gyr_post_vol_LM | -0.332 | 1.43E-06 | 8.30E-06 |
| T1_GM_parcellation_R_Precuneous_vol_LM | -0.369 | 1.54E-06 | 8.83E-06 |
| T1_GM_parcellation_R_Temp_Pole_vol_LM | -0.343 | 1.87E-06 | 1.05E-05 |
| IDP_dMRI_ProbtrackX_MD_atr_r | 0.467 | 1.95E-07 | 1.42E-05 |
| FS_HippSubfield_rh_volume_GC.ML.DG.head. | -0.336 | 3.34E-06 | 1.86E-05 |
| T2_FLAIR_WMHvol_Sagittal_stratum_R | 0.347 | 1.00E-05 | 1.97E-05 |
| T2_FLAIR_WMHvol_Retrolenticular_part_of_internal_capsule_L | 0.361 | 1.08E-05 | 1.98E-05 |
| T2_FLAIR_WMHvol_Tapetum_L | 0.305 | 1.06E-05 | 1.98E-05 |
| FS_aparc.DKTatlas_lh_thickness_superiortemporal. | -0.334 | 3.77E-06 | 2.07E-05 |
| T1_GM_parcellation_R_Heschl_Gyr_vol_LM | -0.329 | 3.90E-06 | 2.11E-05 |
| IDP_dMRI_ProbtrackX_MD_ifo_r | 0.448 | 5.56E-07 | 2.42E-05 |
| dMRI_MD_precuneousR | 0.516 | 4.57E-07 | 2.42E-05 |
| IDP_dMRI_ProbtrackX_MD_str_r | 0.445 | 6.75E-07 | 2.45E-05 |
| T1_GM_parcellation_R_Intracalcarine_vol_LM | -0.306 | 4.91E-06 | 2.62E-05 |
| FS_aparc.DKTatlas_rh_thickness_superiortemporal. | -0.320 | 4.99E-06 | 2.63E-05 |
| T1_GM_parcellation_Vermis_VI_Cerebellum_vol_LM | -0.305 | 5.56E-06 | 2.89E-05 |
| T1_GM_parcellation_L_Front_Orbital_vol_LM | -0.367 | 5.64E-06 | 2.90E-05 |
| IDP_dMRI_ProbtrackX_MD_cgh_r | 0.495 | 1.22E-06 | 2.95E-05 |

| | | | |
|---|---|---|---|
| IDP_dMRI_ProbtrackX_MD_ifo_l | 0.441 | 1.22E-06 | 2.95E-05 |
| IDP_dMRI_ProbtrackX_MD_unc_r | 0.437 | 1.02E-06 | 2.95E-05 |
| T1_GM_parcellation_L_Temp_Pole_vol_LM | -0.348 | 7.12E-06 | 3.61E-05 |
| T1_GM_parcellation_R_Occ_Fusiform_Gyr_vol_LM | -0.315 | 7.51E-06 | 3.75E-05 |
| T1_GM_parcellation_L_Postcentral_Gyr_vol_LM | -0.316 | 7.90E-06 | 3.89E-05 |
| T1_GM_parcellation_L_VIIIb_Cerebellum_vol_LM | -0.295 | 7.97E-06 | 3.89E-05 |
| T1_GM_parcellation_R_X_Cerebellum_vol_LM | -0.322 | 8.07E-06 | 3.89E-05 |
| FS_HippSubfield_rh_volume_CA3.body. | -0.306 | 8.75E-06 | 4.17E-05 |
| FS_HippSubfield_lh_volume_presubiculum.head. | -0.332 | 9.27E-06 | 4.36E-05 |
| T1_GM_parcellation_L_Planum_Polare_vol_LM | -0.344 | 9.73E-06 | 4.52E-05 |
| FS_aparc.DKTatlas_lh_volume_precentral. | -0.324 | 1.00E-05 | 4.60E-05 |
| T1_GM_parcellation_L_Temp_Fusiform_ant_vol_LM | -0.315 | 1.02E-05 | 4.65E-05 |
| FS_aparc.DKTatlas_lh_thickness_superiorfrontal. | -0.313 | 1.13E-05 | 5.01E-05 |
| T1_GM_parcellation_L_Central_Opercular_vol_LM | -0.354 | 1.12E-05 | 5.01E-05 |
| IDP_dMRI_ProbtrackX_MD_str_l | 0.415 | 2.39E-06 | 5.04E-05 |
| IDP_dMRI_ProbtrackX_ICVF_atr_l | -0.423 | 2.54E-06 | 5.04E-05 |
| T2_FLAIR_WMHvol_Tapetum_R | 0.288 | 3.04E-05 | 5.35E-05 |
| FS_aparc.DKTatlas_rh_volume_paraFS_Hippocampal. | -0.302 | 1.24E-05 | 5.44E-05 |
| FS_aparc.DKTatlas_lh_thickness_paracentral. | -0.297 | 1.31E-05 | 5.69E-05 |
| FS_HippSubfield_lh_volume_Whole.FS_Hippocampal.head. | -0.331 | 1.41E-05 | 6.00E-05 |
| T1_GM_parcellation_L_Intracalcarine_vol_LM | -0.299 | 1.41E-05 | 6.00E-05 |
| FS_HippSubfield_rh_volume_molecular.layer.HP.body. | -0.306 | 1.47E-05 | 6.20E-05 |
| T1_GM_parcellation_R_Temp_Fusiform_post_vol_LM | -0.326 | 1.66E-05 | 6.92E-05 |
| FS_HippSubfield_rh_volume_CA4.head. | -0.312 | 1.92E-05 | 7.89E-05 |
| T1_GM_parcellation_L_Juxtapositional_Lobule_vol_LM | -0.302 | 2.06E-05 | 8.40E-05 |
| FS_aparc.DKTatlas_rh_thickness_superiorfrontal. | -0.298 | 2.27E-05 | 9.15E-05 |
| FS_HippSubfield_rh_volume_molecular.layer.HP.head. | -0.302 | 2.58E-05 | 1.03E-04 |
| FS_aparc.DKTatlas_rh_thickness_transversetemporal. | -0.285 | 2.71E-05 | 1.07E-04 |
| T1_GM_parcellation_Vermis_Crus_II_Cerebellum_vol_LM | -0.278 | 3.02E-05 | 1.18E-04 |
| FS_HippSubfield_lh_volume_GC.ML.DG.head. | -0.313 | 3.29E-05 | 1.27E-04 |
| T1_GM_parcellation_L_Lingual_Gyr_vol_LM | -0.336 | 3.34E-05 | 1.28E-04 |
| T1_NBMvol_L | -0.302 | 3.62E-05 | 1.37E-04 |
| FS_aparc.DKTatlas_lh_volume_paracentral. | -0.300 | 3.72E-05 | 1.39E-04 |
| FS_aparc.DKTatlas_rh_volume_fusiform. | -0.311 | 3.74E-05 | 1.39E-04 |
| T1_GM_parcellation_L_Temp_Occ_Fusiform_vol_LM | -0.304 | 4.11E-05 | 1.50E-04 |
| L_hipp_masked | -0.295 | 4.11E-05 | 1.50E-04 |
| FS_aparc.DKTatlas_lh_volume_entorhinal. | -0.298 | 4.40E-05 | 1.60E-04 |
| FS_HippSubfield_lh_volume_CA4.head. | -0.313 | 4.54E-05 | 1.63E-04 |
| FS_aparc.DKTatlas_rh_thickness_supramarginal. | -0.287 | 4.62E-05 | 1.64E-04 |

| | | | |
|---|---|---|---|
| T1_GM_parcellation_R_Lateral_Occ_Inf_vol_LM | -0.310 | 4.93E-05 | 1.74E-04 |
| FS_HippSubfield_lh_volume_CA3.head. | -0.297 | 5.13E-05 | 1.78E-04 |
| FS_aparc.DKTatlas_lh_thickness_postcentral. | -0.274 | 5.14E-05 | 1.78E-04 |
| IDP_dMRI_ProbtrackX_ICVF_atr_r | -0.402 | 1.11E-05 | 1.86E-04 |
| IDP_dMRI_ProbtrackX_ICVF_ifo_r | -0.401 | 1.03E-05 | 1.86E-04 |
| FS_aparc.DKTatlas_lh_volume_postcentral. | -0.307 | 5.46E-05 | 1.87E-04 |
| FS_HippSubfield_rh_volume_presubiculum.body. | -0.283 | 5.76E-05 | 1.93E-04 |
| T1_GM_parcellation_R_Supramarginal_Gyr_ant_vol_LM | -0.287 | 5.72E-05 | 1.93E-04 |
| T1_GM_parcellation_R_Juxtapositional_Lobule_vol_LM | -0.276 | 5.73E-05 | 1.93E-04 |
| IDP_T1_FIRST_left_thalamus_volume | -0.318 | 6.04E-05 | 2.00E-04 |
| T1_GM_parcellation_R_Supramarginal_Gyr_post_vol_LM | -0.297 | 6.14E-05 | 2.02E-04 |
| T1_SIENAX_brain.unnorm_vol_LM | -0.534 | 6.40E-05 | 2.09E-04 |
| T1_GM_parcellation_L_Paracingulate_Gyr_vol_LM | -0.285 | 6.65E-05 | 2.15E-04 |
| IDP_dMRI_ProbtrackX_MD_unc_l | 0.390 | 1.57E-05 | 2.33E-04 |
| IDP_dMRI_ProbtrackX_ISOVF_atr_l | 0.386 | 1.64E-05 | 2.33E-04 |
| IDP_dMRI_ProbtrackX_ISOVF_atr_r | 0.386 | 1.71E-05 | 2.33E-04 |
| IDP_dMRI_ProbtrackX_ICVF_unc_r | -0.383 | 1.91E-05 | 2.45E-04 |
| FS_aparc.DKTatlas_rh_thickness_inferiortemporal. | -0.283 | 7.69E-05 | 2.47E-04 |
| IDP_dMRI_ProbtrackX_MD_fmi | 0.382 | 2.34E-05 | 2.83E-04 |
| T1_GM_parcellation_R_Paracingulate_Gyr_vol_LM | -0.280 | 1.01E-04 | 3.22E-04 |
| IDP_dMRI_ProbtrackX_MD_cst_l | 0.365 | 3.56E-05 | 3.30E-04 |
| IDP_dMRI_ProbtrackX_ICVF_cgc_l | -0.401 | 3.47E-05 | 3.30E-04 |
| IDP_dMRI_ProbtrackX_ICVF_cgh_r | -0.399 | 3.32E-05 | 3.30E-04 |
| IDP_dMRI_ProbtrackX_ICVF_unc_l | -0.375 | 3.39E-05 | 3.30E-04 |
| IDP_dMRI_ProbtrackX_OD_atr_l | -0.412 | 3.63E-05 | 3.30E-04 |
| IDP_dMRI_ProbtrackX_ISOVF_cgh_r | 0.426 | 2.93E-05 | 3.30E-04 |
| IDP_dMRI_ProbtrackX_OD_atr_r | -0.422 | 3.84E-05 | 3.35E-04 |
| IDP_dMRI_ProbtrackX_ICVF_ifo_l | -0.376 | 4.47E-05 | 3.74E-04 |
| T1_GM_parcellation_Vermis_VIIIb_Cerebellum_vol_LM | -0.256 | 1.24E-04 | 3.93E-04 |
| FS_aparc.DKTatlas_lh_thickness_supramarginal. | -0.271 | 1.31E-04 | 4.11E-04 |
| FS_HippSubfield_lh_volume_CA1.head. | -0.283 | 1.33E-04 | 4.15E-04 |
| FS_aparc.DKTatlas_lh_thickness_transversetemporal. | -0.260 | 1.35E-04 | 4.16E-04 |
| FS_aparc.DKTatlas_rh_thickness_paracentral. | -0.257 | 1.46E-04 | 4.47E-04 |
| FS_aparc.DKTatlas_lh_volume_lateralorbitofrontal. | -0.309 | 1.48E-04 | 4.50E-04 |
| FS_aparc.DKTatlas_rh_volume_precentral. | -0.285 | 1.53E-04 | 4.62E-04 |
| FS_HippSubfield_lh_volume_subiculum.head. | -0.279 | 1.67E-04 | 4.99E-04 |
| FS_HippSubfield_lh_volume_molecular.layer.HP.head. | -0.278 | 1.74E-04 | 5.17E-04 |
| IDP_dMRI_ProbtrackX_ISOVF_unc_r | 0.361 | 6.45E-05 | 5.21E-04 |
| T2_FLAIR_WMHvol_Cingulum_cingulate_gyrus_L | 0.307 | 3.08E-04 | 5.24E-04 |

| | | | |
|---|---|---|---|
| FS_HippSubfield_lh_volume_CA3.body. | -0.269 | 1.82E-04 | 5.32E-04 |
| FS_aparc.DKTatlas_lh_volume_fusiform. | -0.295 | 1.81E-04 | 5.32E-04 |
| T1_GM_parcellation_R_Temp_Occ_Fusiform_vol_LM | -0.284 | 1.93E-04 | 5.60E-04 |
| IDP_dMRI_ProbtrackX_MD_ilf_r | 0.367 | 7.53E-05 | 5.87E-04 |
| FS_aparc.DKTatlas_lh_thickness_rostralmiddlefrontal. | -0.258 | 2.04E-04 | 5.87E-04 |
| T2_FLAIR_WMHvol_Sagittal_stratum_L | 0.299 | 3.71E-04 | 6.11E-04 |
| FS_aparc.DKTatlas_lh_thickness_caudalmiddlefrontal. | -0.258 | 2.19E-04 | 6.26E-04 |
| FS_aparc.DKTatlas_rh_thickness_fusiform. | -0.256 | 2.57E-04 | 7.30E-04 |
| T1_NBMvol_R | -0.253 | 2.97E-04 | 8.39E-04 |
| T1_GM_parcellation_R_IX_Cerebellum_vol_LM | -0.239 | 3.23E-04 | 9.03E-04 |
| T1_GM_parcellation_R_Sup_Temp_Gyr_ant_vol_LM | -0.273 | 3.25E-04 | 9.04E-04 |
| IDP_dMRI_ProbtrackX_ICVF_ptr_r | -0.355 | 1.24E-04 | 9.29E-04 |
| IDP_dMRI_ProbtrackX_ICVF_str_r | -0.349 | 1.28E-04 | 9.29E-04 |
| FS_aparc.DKTatlas_rh_thickness_parsopercularis. | -0.245 | 3.51E-04 | 9.68E-04 |
| FS_aparc.DKTatlas_lh_thickness_parstriangularis. | -0.246 | 3.86E-04 | 1.06E-03 |
| IDP_dMRI_ProbtrackX_MD_ptr_r | 0.347 | 1.55E-04 | 1.09E-03 |
| IDP_T1_FIRST_brain_stem.4th_ventricle_volume | -0.267 | 4.22E-04 | 1.15E-03 |
| FS_aparc.DKTatlas_lh_thickness_lateralorbitofrontal. | -0.242 | 4.29E-04 | 1.16E-03 |
| FS_aparc.DKTatlas_rh_volume_inferiortemporal. | -0.279 | 4.36E-04 | 1.17E-03 |
| IDP_dMRI_ProbtrackX_ICVF_str_l | -0.340 | 1.74E-04 | 1.18E-03 |
| IDP_dMRI_ProbtrackX_ICVF_cgc_r | -0.355 | 1.89E-04 | 1.22E-03 |
| IDP_dMRI_ProbtrackX_ICVF_cgh_l | -0.359 | 1.96E-04 | 1.22E-03 |
| IDP_dMRI_ProbtrackX_ICVF_fmi | -0.338 | 1.93E-04 | 1.22E-03 |
| IDP_dMRI_ProbtrackX_FA_atr_l | -0.347 | 2.07E-04 | 1.25E-03 |
| IDP_dMRI_ProbtrackX_FA_ifo_r | -0.331 | 2.23E-04 | 1.32E-03 |
| T1_GM_parcellation_R_Inf_Temp_Gyr_ant_vol_LM | -0.260 | 5.00E-04 | 1.33E-03 |
| T1_GM_parcellation_L_Front_Operculum_vol_LM | -0.260 | 5.03E-04 | 1.33E-03 |
| FS_aparc.DKTatlas_rh_volume_superiortemporal. | -0.276 | 5.17E-04 | 1.36E-03 |
| FS_HippSubfield_rh_volume_CA1.body. | -0.236 | 5.34E-04 | 1.40E-03 |
| T1_GM_parcellation_R_Insula_vol_LM | -0.272 | 5.48E-04 | 1.42E-03 |
| T1_GM_parcellation_L_Heschl_Gyr_vol_LM | -0.256 | 5.49E-04 | 1.42E-03 |
| dMRI_MD_R_hipp | 0.378 | 2.52E-04 | 1.44E-03 |
| ASL_Right_Thalamus | -0.390 | 5.82E-05 | 1.45E-03 |
| ASL_Left_Thalamus | -0.386 | 3.78E-05 | 1.45E-03 |
| IDP_dMRI_ProbtrackX_FA_fmi | -0.344 | 2.72E-04 | 1.52E-03 |
| IDP_dMRI_ProbtrackX_MD_slf_l | 0.342 | 2.92E-04 | 1.53E-03 |
| IDP_dMRI_ProbtrackX_MD_slf_r | 0.340 | 3.03E-04 | 1.53E-03 |
| IDP_dMRI_ProbtrackX_ICVF_mcp | -0.325 | 3.03E-04 | 1.53E-03 |
| IDP_dMRI_ProbtrackX_ISOVF_str_r | 0.326 | 2.91E-04 | 1.53E-03 |

| | | | |
|---|---|---|---|
| T1_GM_parcellation_L_Precuneous_vol_LM | -0.280 | 6.62E-04 | 1.69E-03 |
| T1_GM_parcellation_R_Front_Operculum_vol_LM | -0.253 | 6.61E-04 | 1.69E-03 |
| FS_aparc.DKTatlas_lh_area_transversetemporal. | 0.257 | 6.67E-04 | 1.69E-03 |
| T1_GM_parcellation_L_Front_Pole_vol_LM | -0.286 | 6.98E-04 | 1.75E-03 |
| T1_GM_parcellation_L_Sup_Parietal_Lobule_vol_LM | -0.250 | 6.95E-04 | 1.75E-03 |
| FS_aparc.DKTatlas_lh_volume_superiorfrontal. | -0.262 | 7.18E-04 | 1.78E-03 |
| FS_aparc.DKTatlas_rh_area_paraFS_Hippocampal. | -0.269 | 7.36E-04 | 1.82E-03 |
| ASL_arrival_gm_Left_Temporal_Lobe | 0.351 | 1.54E-04 | 1.93E-03 |
| ASL_arrival_Left_Putamen | 0.344 | 1.37E-04 | 1.93E-03 |
| FS_aparc.DKTatlas_rh_volume_isthmuscingulate. | -0.245 | 7.87E-04 | 1.93E-03 |
| IDP_T1_FIRST_left_putamen_volume | -0.253 | 8.52E-04 | 2.08E-03 |
| IDP_dMRI_ProbtrackX_FA_cgh_r | -0.349 | 4.48E-04 | 2.22E-03 |
| IDP_dMRI_ProbtrackX_ISOVF_str_l | 0.313 | 5.40E-04 | 2.62E-03 |
| T2_FLAIR_WMHvol_Retrolenticular_part_of_internal_capsule_R | 0.222 | 1.73E-03 | 2.76E-03 |
| T2_FLAIR_WMHvol_Fornix_cres.Stria_terminalis_R | 0.360 | 1.82E-03 | 2.82E-03 |
| IDP_dMRI_ProbtrackX_MD_cst_r | 0.310 | 6.13E-04 | 2.90E-03 |
| ASL_arrival_Right_Caudate | 0.338 | 3.19E-04 | 2.98E-03 |
| ASL_arrival_Left_Caudate | 0.329 | 3.58E-04 | 2.98E-03 |
| IDP_dMRI_ProbtrackX_FA_ptr_r | -0.305 | 6.43E-04 | 2.98E-03 |
| ASL_arrival_Right_Putamen | 0.319 | 4.34E-04 | 3.10E-03 |
| IDP_dMRI_ProbtrackX_ICVF_ilf_r | -0.315 | 7.17E-04 | 3.26E-03 |
| T1_GM_parcellation_L_IX_Cerebellum_vol_LM | -0.212 | 1.36E-03 | 3.29E-03 |
| FS_aparc.DKTatlas_rh_thickness_caudalmiddlefrontal. | -0.223 | 1.38E-03 | 3.32E-03 |
| FS_aparc.DKTatlas_rh_thickness_parsorbitalis. | -0.221 | 1.40E-03 | 3.33E-03 |
| T1_GM_parcellation_L_Parahippocampal_Gyr_post_vol_LM | -0.240 | 1.40E-03 | 3.33E-03 |
| FS_HippSubfield_lh_volume_FS_Hippocampal.tail. | -0.243 | 1.42E-03 | 3.36E-03 |
| T1_GM_parcellation_R_Front_Medial_vol_LM | -0.226 | 1.44E-03 | 3.38E-03 |
| IDP_dMRI_ProbtrackX_FA_ifo_l | -0.304 | 8.10E-04 | 3.60E-03 |
| T1_GM_parcellation_R_Central_Opercular_vol_LM | -0.255 | 1.70E-03 | 3.98E-03 |
| FS_aparc.DKTatlas_lh_volume_lingual. | -0.223 | 1.80E-03 | 4.18E-03 |
| FS_aparc.DKTatlas_lh_thickness_insula. | -0.212 | 1.83E-03 | 4.22E-03 |
| T1_GM_parcellation_R_Lateral_Occ_Sup_vol_LM | -0.236 | 2.05E-03 | 4.72E-03 |
| FS_aparc.DKTatlas_lh_thickness_parsopercularis. | -0.214 | 2.09E-03 | 4.75E-03 |
| FS_aparc.DKTatlas_rh_volume_lateralorbitofrontal. | -0.252 | 2.09E-03 | 4.75E-03 |
| IDP_dMRI_ProbtrackX_ICVF_ptr_l | -0.305 | 1.11E-03 | 4.84E-03 |
| ASL_gm_Right_Occipital_Lobe | -0.311 | 8.83E-04 | 5.52E-03 |
| IDP_dMRI_ProbtrackX_FA_unc_l | -0.293 | 1.31E-03 | 5.59E-03 |
| ASL_arrival_gm_LICA | 0.320 | 1.03E-03 | 5.70E-03 |

| | | | |
|---|---|---|---|
| FS_aparc.DKTatlas_lh_area_lingual. | -0.226 | 2.84E-03 | 6.39E-03 |
| FS_aparc.DKTatlas_rh_area_rostralanteriorcingulate. | -0.231 | 2.85E-03 | 6.39E-03 |
| FS_aparc.DKTatlas_rh_volume_parsorbitalis. | -0.216 | 2.91E-03 | 6.48E-03 |
| IDP_dMRI_ProbtrackX_MD_ar_l | 0.286 | 1.66E-03 | 6.54E-03 |
| IDP_dMRI_ProbtrackX_ICVF_cst_l | -0.286 | 1.68E-03 | 6.54E-03 |
| IDP_dMRI_ProbtrackX_ICVF_slf_l | -0.300 | 1.61E-03 | 6.54E-03 |
| IDP_dMRI_ProbtrackX_ICVF_slf_r | -0.300 | 1.57E-03 | 6.54E-03 |
| dMRI_MD_precuneousL | 0.328 | 1.66E-03 | 6.54E-03 |
| T1_GM_parcellation_R_Planum_Tempe_vol_LM | -0.234 | 2.96E-03 | 6.57E-03 |
| T1_GM_parcellation_Vermis_IX_Cerebellum_vol_LM | -0.198 | 3.02E-03 | 6.65E-03 |
| IDP_dMRI_ProbtrackX_ISOVF_cst_l | 0.286 | 1.83E-03 | 6.98E-03 |
| IDP_T1_FIRST_right_pallidum_volume | -0.201 | 3.24E-03 | 7.10E-03 |
| FS_HippSubfield_rh_volume_HATA. | -0.215 | 3.38E-03 | 7.37E-03 |
| ASL_gm_Left_Occipital_Lobe | -0.291 | 1.71E-03 | 7.63E-03 |
| ASL_arrival_gm_Left_Frontal_Lobe | 0.301 | 1.73E-03 | 7.63E-03 |
| ASL_arrival_gm_Left_Parietal_Lobe | 0.307 | 1.83E-03 | 7.63E-03 |
| FS_aparc.DKTatlas_lh_volume_pericalcarine. | -0.198 | 3.60E-03 | 7.77E-03 |
| T1_GM_parcellation_L_Mid_Front_Gyr_vol_LM | -0.220 | 3.62E-03 | 7.77E-03 |
| T1_GM_parcellation_R_Mid_Temp_Gyr_temporoOcc_part_vol_LM | -0.217 | 3.61E-03 | 7.77E-03 |
| FS_aparc.DKTatlas_rh_thickness_insula. | -0.203 | 3.80E-03 | 8.11E-03 |
| FS_aparc.DKTatlas_lh_volume_medialorbitofrontal. | -0.230 | 3.83E-03 | 8.13E-03 |
| FS_aparc.DKTatlas_rh_volume_paracentral. | -0.209 | 3.89E-03 | 8.22E-03 |
| IDP_dMRI_ProbtrackX_MD_ilf_l | 0.290 | 2.23E-03 | 8.25E-03 |
| IDP_dMRI_ProbtrackX_MD_ptr_l | 0.295 | 2.23E-03 | 8.25E-03 |
| IDP_dMRI_ProbtrackX_MD_mcp | 0.272 | 2.54E-03 | 9.24E-03 |
| T1_GM_parcellation_R_Subcallosal_vol_LM | -0.251 | 4.44E-03 | 9.32E-03 |
| IDP_dMRI_ProbtrackX_FA_cst_l | -0.266 | 2.80E-03 | 9.69E-03 |
| IDP_dMRI_ProbtrackX_MD_cgc_l | 0.290 | 2.78E-03 | 9.69E-03 |
| dMRI_MD_R_amyg | 0.315 | 2.80E-03 | 9.69E-03 |
| FS_aparc.DKTatlas_lh_thickness_precuneus. | -0.198 | 4.71E-03 | 9.80E-03 |
| T1_GM_parcellation_L_Occ_Pole_vol_LM | -0.203 | 4.70E-03 | 9.80E-03 |
| ASL_arrival_gm_70..GM | 0.277 | 2.83E-03 | 1.01E-02 |
| ASL_arrival_gm_gmmask_pure_cort | 0.281 | 2.65E-03 | 1.01E-02 |
| IDP_dMRI_ProbtrackX_FA_atr_r | -0.278 | 2.99E-03 | 1.02E-02 |
| T1_GM_parcellation_R_Sup_Temp_Gyr_post_vol_LM | -0.213 | 5.20E-03 | 1.07E-02 |
| ASL_gm_VBA | -0.277 | 3.34E-03 | 1.11E-02 |
| FS_aparc.DKTatlas_lh_area_pericalcarine. | -0.190 | 5.63E-03 | 1.16E-02 |
| dMRI_MD_parahippL | 0.310 | 3.46E-03 | 1.16E-02 |

| | | | |
|---|---|---|---|
| FS_HippSubfield_lh_volume_molecular.layer.HP.body. | -0.201 | 5.70E-03 | 1.17E-02 |
| FS_aparc.DKTatlas_lh_thickness_medialorbitofrontal. | -0.198 | 5.75E-03 | 1.17E-02 |
| ASL_gm_gmmask_pure_cort | -0.277 | 3.81E-03 | 1.19E-02 |
| IDP_dMRI_ProbtrackX_OD_mcp | -0.269 | 3.69E-03 | 1.22E-02 |
| FS_aparc.DKTatlas_rh_volume_superiorfrontal. | -0.233 | 6.33E-03 | 1.28E-02 |
| IDP_dMRI_ProbtrackX_ICVF_fma | -0.268 | 3.95E-03 | 1.29E-02 |
| ASL_Left_Caudate | -0.283 | 4.37E-03 | 1.29E-02 |
| IDP_dMRI_ProbtrackX_FA_ptr_l | -0.263 | 4.04E-03 | 1.30E-02 |
| FS_aparc.DKTatlas_lh_thickness_parsorbitalis. | -0.190 | 6.53E-03 | 1.32E-02 |
| IDP_T1_FIRST_right_putamen_volume | -0.194 | 6.67E-03 | 1.34E-02 |
| T1_GM_parcellation_L_Front_Medial_vol_LM | -0.194 | 6.95E-03 | 1.39E-02 |
| FS_aparc.DKTatlas_rh_volume_parstriangularis. | -0.200 | 7.45E-03 | 1.48E-02 |
| FS_aparc.DKTatlas_lh_volume_rostralanteriorcingulate. | -0.227 | 7.65E-03 | 1.51E-02 |
| IDP_dMRI_ProbtrackX_ISOVF_ptr_r | 0.257 | 4.87E-03 | 1.54E-02 |
| FS_aparc.DKTatlas_lh_volume_rostralmiddlefrontal. | -0.209 | 7.88E-03 | 1.54E-02 |
| FS_aparc.DKTatlas_rh_area_lingual. | -0.199 | 7.90E-03 | 1.54E-02 |
| IDP_dMRI_ProbtrackX_ICVF_ilf_l | -0.266 | 5.06E-03 | 1.58E-02 |
| FS_aparc.DKTatlas_lh_volume_inferiortemporal. | -0.210 | 8.24E-03 | 1.60E-02 |
| FS_aparc.DKTatlas_rh_thickness_precuneus. | -0.185 | 8.26E-03 | 1.60E-02 |
| FS_aparc.DKTatlas_rh_area_pericalcarine. | -0.183 | 8.30E-03 | 1.60E-02 |
| ASL_gm_70..GM | -0.263 | 5.77E-03 | 1.60E-02 |
| T1_GM_parcellation_L_Sup_Temp_Gyr_post_vol_LM | -0.203 | 8.48E-03 | 1.62E-02 |
| T1_GM_parcellation_L_Supracalcarine_vol_LM | -0.193 | 8.45E-03 | 1.62E-02 |
| FS_aparc.DKTatlas_rh_area_lateralorbitofrontal. | -0.239 | 8.67E-03 | 1.65E-02 |
| IDP_dMRI_ProbtrackX_ISOVF_ifo_r | 0.255 | 5.57E-03 | 1.71E-02 |
| T1_GM_parcellation_R_Inf_Front_Gyr_pars_triangularis_vol_LM | -0.181 | 9.18E-03 | 1.74E-02 |
| IDP_dMRI_ProbtrackX_OD_ilf_r | -0.270 | 5.87E-03 | 1.78E-02 |
| IDP_dMRI_ProbtrackX_ICVF_cst_r | -0.260 | 6.22E-03 | 1.86E-02 |
| ASL_wm_Left_Cerebral_White_Matter_80.. | -0.247 | 7.75E-03 | 1.94E-02 |
| ASL_arrival_gm_Right_Occipital_Lobe | 0.252 | 7.38E-03 | 1.94E-02 |
| IDP_dMRI_ProbtrackX_ISOVF_ilf_r | 0.252 | 6.61E-03 | 1.95E-02 |
| IDP_dMRI_ProbtrackX_MO_mcp | 0.247 | 6.86E-03 | 1.97E-02 |
| IDP_dMRI_ProbtrackX_ISOVF_unc_l | 0.248 | 6.78E-03 | 1.97E-02 |
| FS_aparc.DKTatlas_lh_area_fusiform. | -0.230 | 1.05E-02 | 1.97E-02 |
| IDP_dMRI_ProbtrackX_OD_slf_l | -0.259 | 7.01E-03 | 1.99E-02 |
| FS_aparc.DKTatlas_lh_area_superiortemporal. | 0.240 | 1.06E-02 | 1.99E-02 |
| T1_GM_parcellation_L_Pallidum_vol_LM | 0.178 | 1.09E-02 | 2.03E-02 |
| T1_GM_parcellation_R_Pallidum_vol_LM | 0.183 | 1.10E-02 | 2.04E-02 |

| | | | |
|---|---|---|---|
| ASL_arrival_gm_Left_Occipital_Lobe | 0.244 | 8.67E-03 | 2.06E-02 |
| T1_GM_parcellation_R_Mid_Temp_Gyr_ant_vol_LM | -0.191 | 1.13E-02 | 2.08E-02 |
| IDP_dMRI_ProbtrackX_ISOVF_cst_r | 0.244 | 7.69E-03 | 2.15E-02 |
| T1_SIENAX_CSF_unnorm_vol_LM | 0.220 | 1.18E-02 | 2.17E-02 |
| IDP_dMRI_ProbtrackX_MD_cgc_r | 0.255 | 7.99E-03 | 2.20E-02 |
| IDP_dMRI_ProbtrackX_MD_ml_l | 0.241 | 8.38E-03 | 2.25E-02 |
| IDP_dMRI_ProbtrackX_ICVF_ar_l | -0.249 | 8.30E-03 | 2.25E-02 |
| FS_HippSubfield_rh_volume_FS_Hippocampal.tail. | -0.180 | 1.24E-02 | 2.27E-02 |
| T1_GM_parcellation_L_Temp_Fusiform_post_vol_LM | -0.197 | 1.25E-02 | 2.28E-02 |
| FS_aparc.DKTatlas_rh_thickness_inferiorparietal. | -0.179 | 1.26E-02 | 2.28E-02 |
| IDP_SWI_T2star_right_amygdala.x | -0.220 | 7.66E-04 | 2.30E-02 |
| ASL_Right_Caudate | -0.250 | 1.02E-02 | 2.31E-02 |
| FS_aparc.DKTatlas_rh_thickness_rostralmiddlefrontal. | -0.173 | 1.30E-02 | 2.34E-02 |
| T1_GM_parcellation_L_Mid_Temp_Gyr_ant_vol_LM | -0.205 | 1.31E-02 | 2.35E-02 |
| FS_aparc.DKTatlas_rh_volume_inferiorparietal. | -0.208 | 1.34E-02 | 2.40E-02 |
| FS_aparc.DKTatlas_rh_volume_precuneus. | -0.196 | 1.38E-02 | 2.46E-02 |
| ASL_wm_wmmask_pure_cereb | -0.233 | 1.18E-02 | 2.46E-02 |
| ASL_arrival_Right_Thalamus | 0.232 | 1.15E-02 | 2.46E-02 |
| T1_GM_parcellation_Vermis_X_Cerebellum_vol_LM | -0.164 | 1.41E-02 | 2.49E-02 |
| T1_GM_parcellation_R_Front_Pole_vol_LM | -0.213 | 1.42E-02 | 2.51E-02 |
| IDP_dMRI_ProbtrackX_OD_ptr_l | -0.257 | 9.49E-03 | 2.52E-02 |
| T1_GM_parcellation_L_Sup_Front_Gyr_vol_LM | -0.186 | 1.44E-02 | 2.54E-02 |
| FS_aparc.DKTatlas_rh_thickness_paraFS_Hippocampal. | -0.169 | 1.48E-02 | 2.59E-02 |
| FS_aparc.DKTatlas_rh_thickness_postcentral. | -0.167 | 1.51E-02 | 2.63E-02 |
| T1_GM_parcellation_R_Angular_Gyr_vol_LM | -0.179 | 1.53E-02 | 2.64E-02 |
| IDP_dMRI_ProbtrackX_OD_ifo_r | -0.256 | 1.02E-02 | 2.67E-02 |
| FS_aparc.DKTatlas_lh_volume_cuneus. | -0.170 | 1.56E-02 | 2.70E-02 |
| ASL_gm_Left_Parietal_Lobe | -0.224 | 1.52E-02 | 2.71E-02 |
| ASL_wm_Right_Cerebral_White_Matter_80.. | -0.224 | 1.47E-02 | 2.71E-02 |
| ASL_arrival_gm_Right_Frontal_Lobe | 0.233 | 1.42E-02 | 2.71E-02 |
| ASL_arrival_gm_Right_Temporal_Lobe | 0.231 | 1.41E-02 | 2.71E-02 |
| ASL_wm_90..WM | -0.223 | 1.69E-02 | 2.73E-02 |
| ASL_arrival_gm_RICA | 0.230 | 1.60E-02 | 2.73E-02 |
| ASL_arrival_Left_Thalamus | 0.218 | 1.69E-02 | 2.73E-02 |
| IDP_dMRI_ProbtrackX_FA_slf_r | -0.240 | 1.06E-02 | 2.76E-02 |
| FS_HippSubfield_lh_volume_CA1.body. | -0.167 | 1.67E-02 | 2.87E-02 |
| IDP_dMRI_ProbtrackX_FA_unc_r | -0.233 | 1.15E-02 | 2.94E-02 |
| T1_SIENAX_WM_unnorm_vol_LM | 0.267 | 1.73E-02 | 2.96E-02 |
| T1_GM_parcellation_L_Cuneal_vol_LM | -0.173 | 1.80E-02 | 3.06E-02 |

| | | | |
|---|---|---|---|
| FS_aparc.DKTatlas_rh_area_isthmuscingulate. | -0.189 | 1.84E-02 | 3.11E-02 |
| T1_GM_parcellation_R_Parietal_Operculum_vol_LM | -0.179 | 1.85E-02 | 3.12E-02 |
| IDP_dMRI_ProbtrackX_FA_ilf_r | -0.226 | 1.32E-02 | 3.32E-02 |
| IDP_dMRI_ProbtrackX_MD_cgh_l | 0.247 | 1.32E-02 | 3.32E-02 |
| T1_GM_parcellation_L_Lateral_Occ_Sup_vol_LM | -0.189 | 1.99E-02 | 3.35E-02 |
| dMRI_MD_L_amyg | 0.260 | 1.38E-02 | 3.43E-02 |
| T1_GM_parcellation_L_Inf_Front_Gyr_pars_opercularis_vol_LM | -0.168 | 2.08E-02 | 3.48E-02 |
| FS_aparc.DKTatlas_rh_area_caudalanteriorcingulate. | -0.173 | 2.09E-02 | 3.48E-02 |
| T1_GM_parcellation_L_Occ_Fusiform_Gyr_vol_LM | -0.179 | 2.19E-02 | 3.64E-02 |
| IDP_dMRI_ProbtrackX_FA_ilf_l | -0.221 | 1.51E-02 | 3.69E-02 |
| FS_aparc.DKTatlas_lh_thickness_fusiform. | -0.168 | 2.42E-02 | 3.99E-02 |
| T1_GM_parcellation_R_Sup_Front_Gyr_vol_LM | -0.168 | 2.45E-02 | 4.03E-02 |
| ASL_arrival_gm_Right_Parietal_Lobe | 0.219 | 2.64E-02 | 4.12E-02 |

#### Supplementary Table 5: Associations with ACE-III

Associations between IDPs and ACE-III total cognitive score. Corr-pval are after hierarchical FDR correction.

| Variable | coef | pval | corr-pval |
|---|---|---|---|
| T1_GMvol_temp | 0.648 | 1.06E-13 | 4.21E-11 |
| T1_GM_parcellation_L_Mid_Temp_Gyr_post_vol_LM | 0.540 | 2.79E-13 | 5.51E-11 |
| T1_SIENAX_peripheral_GM_unnorm_vol_LM | 0.642 | 4.62E-12 | 6.08E-10 |
| FS_aparc.DKTatlas_lh_thickness_middletemporal. | 0.429 | 6.20E-10 | 6.12E-08 |
| T1_SIENAX_GM_unnorm_vol_LM | 0.617 | 9.72E-10 | 7.68E-08 |
| T1_GM_parcellation_L_Lateral_Occ_Sup_vol_LM | 0.435 | 3.33E-09 | 2.19E-07 |
| T1_GMvol_par | 0.531 | 5.46E-09 | 3.08E-07 |
| T1_GM_parcellation_L_Sup_Temp_Gyr_post_vol_LM | 0.398 | 2.23E-08 | 1.10E-06 |
| T2_FLAIR_WMHvol_Splenium_of_corpus_callosum | -0.420 | 2.22E-08 | 1.13E-06 |
| T1_GM_parcellation_L_Inf_Temp_Gyr_post_vol_LM | 0.417 | 2.64E-08 | 1.16E-06 |
| FS_aparc.DKTatlas_lh_volume_superiortemporal. | 0.415 | 6.44E-08 | 2.12E-06 |
| T1_GM_parcellation_L_Temp_Pole_vol_LM | 0.411 | 6.14E-08 | 2.12E-06 |
| T1_GM_parcellation_R_Mid_Temp_Gyr_post_vol_LM | 0.404 | 5.85E-08 | 2.12E-06 |
| T1_GM_parcellation_L_Insula_vol_LM | 0.414 | 7.71E-08 | 2.34E-06 |
| FS_aparc.DKTatlas_lh_volume_middletemporal. | 0.402 | 3.03E-07 | 8.55E-06 |
| FS_HippSubfield_lh_volume_Whole.FS_Hippocampal.body. | 0.394 | 3.62E-07 | 9.53E-06 |
| T1_GM_parcellation_L_Mid_Temp_Gyr_ant_vol_LM | 0.392 | 4.36E-07 | 1.06E-05 |
| T1_GM_parcellation_L_Sup_Parietal_Lobule_vol_LM | 0.357 | 4.56E-07 | 1.06E-05 |
| IDP_dMRI_ProbtrackX_MD_cgh_r | -0.500 | 5.15E-08 | 1.12E-05 |
| FS_HippSubfield_lh_volume_GC.ML.DG.body. | 0.392 | 5.87E-07 | 1.29E-05 |
| T1_GM_parcellation_L_Sup_Temp_Gyr_ant_vol_LM | 0.361 | 6.64E-07 | 1.38E-05 |
| FS_HippSubfield_lh_volume_subiculum.body. | 0.385 | 7.96E-07 | 1.57E-05 |
| FS_aparc.DKTatlas_lh_thickness_fusiform. | 0.345 | 8.83E-07 | 1.66E-05 |
| T1_GM_parcellation_L_Front_Pole_vol_LM | 0.399 | 9.56E-07 | 1.72E-05 |
| T1_GMvol_front | 0.444 | 1.18E-06 | 2.02E-05 |
| dMRI_PSMD | -0.511 | 1.89E-07 | 2.06E-05 |
| T1_GM_parcellation_L_Temp_Fusiform_post_vol_LM | 0.362 | 1.40E-06 | 2.30E-05 |
| FS_HippSubfield_lh_volume_CA4.body. | 0.369 | 2.36E-06 | 3.61E-05 |
| L_hipp_masked | 0.329 | 2.38E-06 | 3.61E-05 |
| FS_HippSubfield_lh_volume_subiculum.head. | 0.340 | 3.12E-06 | 4.17E-05 |
| FS_aparc.DKTatlas_lh_thickness_inferiortemporal. | 0.327 | 3.08E-06 | 4.17E-05 |
| T1_GM_parcellation_L_Subcallosal_vol_LM | 0.393 | 2.91E-06 | 4.17E-05 |
| T1_GM_parcellation_L_Precuneous_vol_LM | 0.373 | 3.17E-06 | 4.17E-05 |
| T1_GM_parcellation_R_Front_Orbital_vol_LM | 0.361 | 5.88E-06 | 7.49E-05 |

| | | | |
|---|---|---|---|
| FS_HippSubfield_lh_volume_Whole.FS_Hippocampus. | 0.356 | 6.23E-06 | 7.50E-05 |
| FS_aparc.DKTatlas_lh_volume_inferiortemporal. | 0.346 | 6.26E-06 | 7.50E-05 |
| IDP_dMRI_ProbtrackX_MD_cgh_l | -0.406 | 1.07E-06 | 7.80E-05 |
| FS_HippSubfield_lh_volume_presubiculum.head. | 0.340 | 7.19E-06 | 8.35E-05 |
| FS_aparc.DKTatlas_lh_thickness_inferiorparietal. | 0.305 | 1.27E-05 | 1.43E-04 |
| FS_aparc.DKTatlas_rh_volume_middletemporal. | 0.351 | 1.56E-05 | 1.71E-04 |
| IDP_T1_FIRST_left_putamen_volume | 0.321 | 1.77E-05 | 1.81E-04 |
| FS_aparc.DKTatlas_lh_thickness_superiortemporal. | 0.318 | 1.79E-05 | 1.81E-04 |
| T1_GM_parcellation_R_Inf_Temp_Gyr_ant_vol_LM | 0.317 | 1.83E-05 | 1.81E-04 |
| T1_GM_parcellation_L_Parahippocampal_Gyr_ant_vol_LM | 0.350 | 1.70E-05 | 1.81E-04 |
| FS_aparc.DKTatlas_lh_volume_fusiform. | 0.335 | 2.05E-05 | 1.98E-04 |
| T1_SIENAX_CSF_unnorm_vol_LM | -0.359 | 2.25E-05 | 2.08E-04 |
| T1_GM_parcellation_L_Inf_Temp_Gyr_ant_vol_LM | 0.308 | 2.27E-05 | 2.08E-04 |
| FS_aparc.DKTatlas_lh_volume_superiorparietal. | 0.321 | 2.32E-05 | 2.09E-04 |
| T1_GM_parcellation_R_Inf_Temp_Gyr_post_vol_LM | 0.331 | 2.46E-05 | 2.11E-04 |
| T1_GM_parcellation_L_Amygdala_vol_LM | 0.341 | 2.46E-05 | 2.11E-04 |
| T1_GMvol_occ | 0.379 | 2.97E-05 | 2.49E-04 |
| T1_GM_parcellation_L_Parietal_Operculum_vol_LM | 0.305 | 3.95E-05 | 3.25E-04 |
| T1_GM_parcellation_L_Parahippocampal_Gyr_post_vol_LM | 0.304 | 4.04E-05 | 3.25E-04 |
| WMH_pvent | -0.331 | 1.92E-05 | 3.27E-04 |
| T2_FLAIR_WMHvol_Posterior_thalamic_radiation_L | -0.327 | 1.36E-05 | 3.27E-04 |
| FS_HippSubfield_lh_volume_Whole.FS_Hippocampal.head. | 0.318 | 4.55E-05 | 3.31E-04 |
| T1_GM_parcellation_R_Front_Pole_vol_LM | 0.344 | 4.49E-05 | 3.31E-04 |
| T1_GM_parcellation_R_Sup_Temp_Gyr_post_vol_LM | 0.305 | 4.29E-05 | 3.31E-04 |
| T1_GM_parcellation_L_Front_Orbital_vol_LM | 0.339 | 4.49E-05 | 3.31E-04 |
| T1_GM_parcellation_L_Caudate_vol_LM | 0.346 | 4.61E-05 | 3.31E-04 |
| T1_GM_parcellation_R_Caudate_vol_LM | 0.347 | 4.46E-05 | 3.31E-04 |
| T1_GM_parcellation_L_Front_Operculum_vol_LM | 0.302 | 4.87E-05 | 3.44E-04 |
| FS_HippSubfield_lh_volume_CA4.head. | 0.316 | 5.04E-05 | 3.50E-04 |
| T1_GM_parcellation_L_Hippocampus_vol_LM | 0.326 | 5.35E-05 | 3.65E-04 |
| FS_HippSubfield_rh_volume_GC.ML.DG.body. | 0.324 | 5.61E-05 | 3.69E-04 |
| T1_GM_parcellation_R_Temp_Fusiform_post_vol_LM | 0.312 | 5.58E-05 | 3.69E-04 |
| IDP_T1_FIRST_left_accumbens_volume | 0.292 | 5.77E-05 | 3.74E-04 |
| T1_GM_parcellation_R_Temp_Pole_vol_LM | 0.300 | 6.14E-05 | 3.91E-04 |
| FS_HippSubfield_lh_volume_CA3.body. | 0.288 | 6.99E-05 | 4.31E-04 |
| FS_aparc.DKTatlas_rh_volume_inferiorparietal. | 0.327 | 6.96E-05 | 4.31E-04 |
| FS_HippSubfield_rh_volume_subiculum.body. | 0.312 | 7.57E-05 | 4.60E-04 |
| FS_HippSubfield_lh_volume_GC.ML.DG.head. | 0.304 | 7.71E-05 | 4.62E-04 |
| IDP_T1_FIRST_right_accumbens_volume | 0.281 | 9.13E-05 | 5.38E-04 |

| | | | |
|---|---|---|---|
| IDP_dMRI_ProbtrackX_ISOVF_cgh_r | -0.413 | 1.19E-05 | 6.47E-04 |
| T2_FLAIR_WMHvol_Body_of_corpus_callosum | -0.307 | 5.37E-05 | 6.85E-04 |
| WMH_total | -0.304 | 8.18E-05 | 8.34E-04 |
| FS_aparc.DKTatlas_lh_volume_entorhinal. | 0.283 | 1.50E-04 | 8.68E-04 |
| T1_GM_parcellation_R_Insula_vol_LM | 0.300 | 1.52E-04 | 8.68E-04 |
| T1_GM_parcellation_L_Central_Opercular_vol_LM | 0.313 | 1.74E-04 | 9.80E-04 |
| FS_HippSubfield_lh_volume_FS_Hippocampal.tail. | 0.284 | 1.88E-04 | 1.05E-03 |
| IDP_dMRI_ProbtrackX_ICVF_cgh_l | 0.371 | 2.47E-05 | 1.08E-03 |
| T1_GM_parcellation_R_Mid_Temp_Gyr_ant_vol_LM | 0.277 | 1.97E-04 | 1.08E-03 |
| FS_HippSubfield_lh_volume_CA1.head. | 0.279 | 2.22E-04 | 1.20E-03 |
| T1_GM_parcellation_R_Lateral_Occ_Sup_vol_LM | 0.281 | 2.37E-04 | 1.27E-03 |
| rfMRI_d100_NodeAmplitudes_28 | 0.427 | 4.07E-07 | 1.41E-03 |
| T1_GM_parcellation_R_Sup_Temp_Gyr_ant_vol_LM | 0.279 | 2.90E-04 | 1.53E-03 |
| FS_aparc.DKTatlas_lh_thickness_entorhinal. | 0.267 | 2.99E-04 | 1.55E-03 |
| FS_HippSubfield_rh_volume_Whole.FS_Hippocampal.body. | 0.290 | 3.09E-04 | 1.58E-03 |
| FS_aparc.DKTatlas_lh_volume_supramarginal. | 0.279 | 3.11E-04 | 1.58E-03 |
| FS_aparc.DKTatlas_rh_volume_posteriorcingulate. | 0.258 | 3.22E-04 | 1.59E-03 |
| T1_GM_parcellation_L_Planum_Tempe_vol_LM | 0.281 | 3.18E-04 | 1.59E-03 |
| IDP_dMRI_ProbtrackX_MO_fmi | 0.350 | 4.42E-05 | 1.61E-03 |
| IDP_T1_FIRST_left_thalamus_volume | 0.290 | 3.54E-04 | 1.72E-03 |
| T1_GM_parcellation_R_Front_Operculum_vol_LM | 0.268 | 3.56E-04 | 1.72E-03 |
| T2_FLAIR_WMHvol_Posterior_thalamic_radiation_R | -0.279 | 2.06E-04 | 1.75E-03 |
| IDP_dMRI_ProbtrackX_FA_cgh_l | 0.351 | 6.84E-05 | 1.97E-03 |
| IDP_dMRI_ProbtrackX_ICVF_cgh_r | 0.358 | 8.10E-05 | 1.97E-03 |
| dMRI_MO_supramargR | 0.379 | 8.12E-05 | 1.97E-03 |
| FS_HippSubfield_lh_volume_presubiculum.body. | 0.254 | 4.31E-04 | 2.05E-03 |
| T1_GM_parcellation_L_Paracingulate_Gyr_vol_LM | 0.258 | 4.43E-04 | 2.09E-03 |
| T2_FLAIR_WMHvol_Anterior_corona_radiata_R | -0.276 | 2.90E-04 | 2.11E-03 |
| T1_GM_parcellation_L_Lateral_Occ_Inf_vol_LM | 0.265 | 4.91E-04 | 2.28E-03 |
| FS_aparc.DKTatlas_lh_volume_parsopercularis. | 0.259 | 5.21E-04 | 2.37E-03 |
| T1_GM_parcellation_L_Inf_Temp_Gyr_temporoOcc_part_vol_LM | 0.254 | 5.19E-04 | 2.37E-03 |
| FS_HippSubfield_rh_volume_CA4.body. | 0.274 | 5.38E-04 | 2.42E-03 |
| FS_aparc.DKTatlas_lh_area_superiortemporal. | 0.322 | 5.57E-04 | 2.47E-03 |
| FS_HippSubfield_lh_volume_molecular.layer.HP.body. | 0.250 | 5.84E-04 | 2.56E-03 |
| T1_GM_parcellation_R_Temp_Fusiform_ant_vol_LM | 0.271 | 5.96E-04 | 2.59E-03 |
| IDP_dMRI_ProbtrackX_FA_cgh_r | 0.351 | 1.26E-04 | 2.75E-03 |
| T1_GM_parcellation_R_Angular_Gyr_vol_LM | 0.249 | 6.69E-04 | 2.87E-03 |
| IDP_dMRI_ProbtrackX_ISOVF_cgh_l | -0.319 | 1.46E-04 | 2.89E-03 |

| | | | |
|---|---|---|---|
| T1_SIENAX_brain.unnorm_vol_LM | 0.465 | 7.44E-04 | 3.16E-03 |
| T1_GM_parcellation_R_Amygdala_vol_LM | 0.276 | 8.39E-04 | 3.52E-03 |
| T1_GM_parcellation_L_Supramarginal_Gyr_post_vol_LM | 0.261 | 8.94E-04 | 3.72E-03 |
| FS_aparc.DKTatlas_lh_thickness_paraFS_Hippocampal. | 0.226 | 9.56E-04 | 3.93E-03 |
| FS_aparc.DKTatlas_rh_area_middletemporal. | 0.297 | 1.09E-03 | 4.44E-03 |
| FS_HippSubfield_lh_volume_CA3.head. | 0.247 | 1.16E-03 | 4.63E-03 |
| FS_HippSubfield_rh_volume_subiculum.head. | 0.251 | 1.15E-03 | 4.63E-03 |
| T2_FLAIR_WMHvol_Superior_corona_radiata_L | -0.257 | 7.43E-04 | 4.74E-03 |
| T1_GM_parcellation_R_Central_Opercular_vol_LM | 0.266 | 1.22E-03 | 4.81E-03 |
| dMRI_MD_supramargL | -0.361 | 2.78E-04 | 5.06E-03 |
| T2_FLAIR_WMHvol_Posterior_corona_radiata_L | -0.247 | 8.95E-04 | 5.07E-03 |
| FS_aparc.DKTatlas_rh_thickness_inferiortemporal. | 0.238 | 1.34E-03 | 5.26E-03 |
| T2_FLAIR_WMHvol_Retrolenticular_part_of_internal_capsule_L | -0.282 | 1.04E-03 | 5.28E-03 |
| T1_GM_parcellation_R_Paracingulate_Gyr_vol_LM | 0.238 | 1.38E-03 | 5.35E-03 |
| T1_GM_parcellation_R_Sup_Parietal_Lobule_vol_LM | 0.223 | 1.43E-03 | 5.47E-03 |
| T1_GM_parcellation_R_Subcallosal_vol_LM | 0.281 | 1.50E-03 | 5.69E-03 |
| FS_aparc.DKTatlas_lh_volume_parstriangularis. | 0.235 | 1.56E-03 | 5.83E-03 |
| FS_aparc.DKTatlas_rh_volume_inferiortemporal. | 0.257 | 1.56E-03 | 5.83E-03 |
| FS_aparc.DKTatlas_lh_area_inferiortemporal. | 0.267 | 1.61E-03 | 5.93E-03 |
| rfMRI_d100_NodeAmplitudes_19 | 0.391 | 3.44E-06 | 5.97E-03 |
| FS_aparc.DKTatlas_rh_area_posteriorcingulate. | 0.257 | 1.66E-03 | 6.07E-03 |
| dMRI_MO_supramargL | 0.340 | 3.63E-04 | 6.09E-03 |
| T1_GM_parcellation_R_Planum_Polare_vol_LM | 0.268 | 1.71E-03 | 6.21E-03 |
| T2_FLAIR_WMHvol_Superior_longitudinal_fasciculus_L | -0.242 | 1.53E-03 | 6.50E-03 |
| T2_FLAIR_WMHvol_Tapetum_L | -0.230 | 1.52E-03 | 6.50E-03 |
| FS_aparc.DKTatlas_lh_volume_posteriorcingulate. | 0.236 | 1.81E-03 | 6.51E-03 |
| T2_FLAIR_WMHvol_Superior_corona_radiata_R | -0.254 | 1.82E-03 | 7.13E-03 |
| FS_HippSubfield_rh_volume_Whole.FS_Hippocampus. | 0.249 | 2.00E-03 | 7.13E-03 |
| FS_aparc.DKTatlas_rh_volume_medialorbitofrontal. | 0.236 | 2.04E-03 | 7.21E-03 |
| T1_GM_parcellation_R_Supramarginal_Gyr_post_vol_LM | 0.236 | 2.18E-03 | 7.62E-03 |
| rfMRI_d100_NodeAmplitudes_21 | 0.385 | 8.33E-06 | 8.11E-03 |
| rfMRI_d100_partialcorr_827 | 0.378 | 9.36E-06 | 8.11E-03 |
| T2_FLAIR_WMHvol_Cingulum_cingulate_gyrus_L | -0.266 | 2.31E-03 | 8.41E-03 |
| FS_aparc.DKTatlas_lh_volume_insula. | 0.238 | 2.54E-03 | 8.79E-03 |
| T2_FLAIR_WMHvol_Retrolenticular_part_of_internal_capsule_R | -0.216 | 2.64E-03 | 8.96E-03 |
| FS_aparc.DKTatlas_rh_thickness_entorhinal. | 0.223 | 2.64E-03 | 8.98E-03 |
| FS_aparc.DKTatlas_rh_thickness_middletemporal. | 0.216 | 2.64E-03 | 8.98E-03 |
| FS_aparc.DKTatlas_lh_volume_medialorbitofrontal. | 0.242 | 2.69E-03 | 9.08E-03 |

| | | | |
|---|---|---|---|
| rfMRI_d100_NodeAmplitudes_44 | 0.387 | 1.58E-05 | 9.14E-03 |
| rfMRI_d25_NodeAmplitudes_6 | 0.369 | 1.58E-05 | 9.14E-03 |
| FS_aparc.DKTatlas_rh_area_inferiorparietal. | 0.266 | 2.84E-03 | 9.50E-03 |
| IDP_T1_FIRST_right_putamen_volume | 0.215 | 2.86E-03 | 9.51E-03 |
| FS_aparc.DKTatlas_lh_area_superiorparietal. | 0.235 | 3.08E-03 | 1.01E-02 |
| rfMRI_d25_NodeAmplitudes_13 | 0.371 | 2.05E-05 | 1.02E-02 |
| T1_GM_parcellation_L_Sup_Front_Gyr_vol_LM | 0.225 | 3.14E-03 | 1.02E-02 |
| T1_GM_parcellation_R_Inf_Front_Gyr_pars_opercularis_vol_LM | 0.213 | 3.15E-03 | 1.02E-02 |
| T2_FLAIR_WMHvol_Posterior_corona_radiata_R | -0.216 | 3.29E-03 | 1.05E-02 |
| T2_FLAIR_WMHvol_Genu_of_corpus_callosum | -0.218 | 3.53E-03 | 1.06E-02 |
| T1_GM_parcellation_R_Inf_Temp_Gyr_temporoOcc_part_vol_LM | 0.233 | 3.44E-03 | 1.10E-02 |
| T1_GM_parcellation_R_Precuneous_vol_LM | 0.240 | 3.56E-03 | 1.13E-02 |
| dMRI_tractvol_fma | 0.367 | 7.49E-04 | 1.17E-02 |
| FS_aparc.DKTatlas_lh_thickness_supramarginal. | 0.213 | 3.74E-03 | 1.17E-02 |
| FS_aparc.DKTatlas_rh_volume_superiortemporal. | 0.238 | 3.72E-03 | 1.17E-02 |
| T1_GM_parcellation_L_Planum_Polare_vol_LM | 0.238 | 3.76E-03 | 1.17E-02 |
| T1_GM_parcellation_R_Parahippocampal_Gyr_ant_vol_LM | 0.236 | 3.85E-03 | 1.19E-02 |
| IDP_dMRI_ProbtrackX_ICVF_cgc_l | 0.315 | 8.19E-04 | 1.19E-02 |
| T1_GM_parcellation_R_Mid_Temp_Gyr_temporoOcc_part_vol_LM | 0.218 | 4.00E-03 | 1.23E-02 |
| T1_GM_parcellation_R_Lingual_Gyr_vol_LM | 0.260 | 4.26E-03 | 1.30E-02 |
| rfMRI_d100_NodeAmplitudes_15 | 0.359 | 3.45E-05 | 1.30E-02 |
| rfMRI_d100_NodeAmplitudes_24 | 0.355 | 3.41E-05 | 1.30E-02 |
| rfMRI_d100_NodeAmplitudes_45 | 0.360 | 3.75E-05 | 1.30E-02 |
| T2_FLAIR_WMHvol_Anterior_corona_radiata_L | -0.211 | 4.61E-03 | 1.31E-02 |
| IDP_dMRI_ProbtrackX_ISOVF_ptr_l | -0.287 | 9.73E-04 | 1.33E-02 |
| IDP_dMRI_ProbtrackX_MD_ptr_l | -0.293 | 1.04E-03 | 1.33E-02 |
| IDP_dMRI_ProbtrackX_OD_ar_l | 0.273 | 1.14E-03 | 1.34E-02 |
| dMRI_tractvol_cst_l | -0.351 | 1.17E-03 | 1.34E-02 |
| R_hipp_masked | 0.209 | 4.48E-03 | 1.35E-02 |
| FS_aparc.DKTatlas_rh_thickness_inferiorparietal. | 0.204 | 4.70E-03 | 1.40E-02 |
| FS_aparc.DKTatlas_rh_thickness_superiortemporal. | 0.211 | 4.75E-03 | 1.40E-02 |
| T1_GM_parcellation_R_Mid_Front_Gyr_vol_LM | 0.229 | 4.79E-03 | 1.40E-02 |
| T1_GM_parcellation_L_Cuneal_vol_LM | 0.207 | 4.77E-03 | 1.40E-02 |
| FS_aparc.DKTatlas_lh_volume_precuneus. | 0.217 | 5.08E-03 | 1.48E-02 |
| IDP_dMRI_ProbtrackX_MD_ilf_l | -0.283 | 1.37E-03 | 1.49E-02 |
| dMRI_MD_parahippR | -0.314 | 1.46E-03 | 1.52E-02 |

| | | | |
|---|---|---|---|
| T1_GM_parcellation_L_Angular_Gyr_vol_LM | 0.204 | 5.44E-03 | 1.57E-02 |
| rfMRI_d100_partialcorr_824 | 0.343 | 5.03E-05 | 1.58E-02 |
| T1_GM_parcellation_L_Front_Medial_vol_LM | 0.201 | 5.56E-03 | 1.59E-02 |
| IDP_dMRI_ProbtrackX_OD_str_r | 0.267 | 1.65E-03 | 1.61E-02 |
| dMRI_MD_precuneousR | -0.346 | 1.70E-03 | 1.61E-02 |
| T1_NBMvol_L | 0.212 | 5.81E-03 | 1.65E-02 |
| rfMRI_d100_NodeAmplitudes_29 | 0.342 | 5.76E-05 | 1.66E-02 |
| FS_aparc.DKTatlas_lh_volume_paraFS_Hippocampal. | 0.192 | 5.97E-03 | 1.68E-02 |
| T1_GM_parcellation_L_Occ_Fusiform_Gyr_vol_LM | 0.215 | 6.01E-03 | 1.68E-02 |
| dMRI_MD_L_hipp | -0.301 | 1.88E-03 | 1.70E-02 |
| T1_GM_parcellation_R_Hippocampus_vol_LM | 0.224 | 6.20E-03 | 1.73E-02 |
| FS_aparc.DKTatlas_lh_area_middletemporal. | 0.245 | 6.27E-03 | 1.73E-02 |
| FS_aparc.DKTatlas_rh_thickness_paraFS_Hippocampal. | 0.189 | 6.78E-03 | 1.86E-02 |
| FS_aparc.DKTatlas_lh_thickness_medialorbitofrontal. | 0.197 | 6.88E-03 | 1.87E-02 |
| FS_HippSubfield_rh_volume_CA1.head. | 0.209 | 7.26E-03 | 1.93E-02 |
| FS_HippSubfield_rh_volume_fimbria. | 0.205 | 7.33E-03 | 1.93E-02 |
| FS_aparc.DKTatlas_lh_area_supramarginal. | 0.243 | 7.22E-03 | 1.93E-02 |
| T1_GM_parcellation_L_Lingual_Gyr_vol_LM | 0.229 | 7.26E-03 | 1.93E-02 |
| T1_GM_parcellation_L_Temp_Fusiform_ant_vol_LM | 0.204 | 7.30E-03 | 1.93E-02 |
| FS_HippSubfield_lh_volume_fimbria. | 0.205 | 7.39E-03 | 1.93E-02 |
| FS_HippSubfield_rh_volume_Whole.FS_Hippocampal.head. | 0.215 | 7.75E-03 | 2.01E-02 |
| FS_aparc.DKTatlas_lh_volume_isthmuscingulate. | 0.206 | 7.84E-03 | 2.03E-02 |
| T1_GM_parcellation_R_Postcentral_Gyr_vol_LM | 0.205 | 7.99E-03 | 2.05E-02 |
| FS_aparc.DKTatlas_rh_thickness_fusiform. | 0.192 | 8.27E-03 | 2.11E-02 |
| T1_GM_parcellation_L_Supramarginal_Gyr_ant_vol_LM | 0.194 | 8.75E-03 | 2.22E-02 |
| rfMRI_d100_NodeAmplitudes_31 | 0.339 | 9.02E-05 | 2.23E-02 |
| rfMRI_d100_fullcorr_824 | 0.335 | 8.83E-05 | 2.23E-02 |
| IDP_dMRI_ProbtrackX_ICVF_slf_l | 0.269 | 2.70E-03 | 2.26E-02 |
| IDP_dMRI_ProbtrackX_OD_str_l | 0.266 | 2.66E-03 | 2.26E-02 |
| FS_HippSubfield_lh_volume_CA1.body. | 0.184 | 9.21E-03 | 2.32E-02 |
| IDP_dMRI_ProbtrackX_ICVF_ilf_l | 0.262 | 2.92E-03 | 2.36E-02 |
| T2_FLAIR_WMHvol_Tapetum_R | -0.190 | 9.16E-03 | 2.46E-02 |
| IDP_dMRI_ProbtrackX_OD_slf_l | 0.262 | 3.22E-03 | 2.50E-02 |
| FS_HippSubfield_rh_volume_CA3.body. | 0.188 | 1.04E-02 | 2.61E-02 |
| T2_FLAIR_WMHvol_Fornix_cres.Stria_terminalis_L | -0.379 | 1.07E-02 | 2.62E-02 |
| T2_FLAIR_WMHvol_Superior_fronto.occipital_fasciculus_R | -0.211 | 1.08E-02 | 2.62E-02 |
| IDP_dMRI_ProbtrackX_MO_cgc_r | 0.259 | 3.52E-03 | 2.64E-02 |
| IDP_dMRI_ProbtrackX_MO_cgc_l | 0.249 | 3.98E-03 | 2.72E-02 |
| IDP_dMRI_ProbtrackX_ICVF_unc_l | 0.257 | 4.02E-03 | 2.72E-02 |

| | | | |
|---|---|---|---|
| dMRI_MD_parahippL | -0.290 | 4.09E-03 | 2.72E-02 |
| dMRI_MD_R_amyg | -0.290 | 4.11E-03 | 2.72E-02 |
| rfMRI_d100_partialcorr_639 | -0.336 | 1.20E-04 | 2.77E-02 |
| T1_GM_parcellation_L_Inf_Front_Gyr_pars_opercularis_vol_LM | 0.186 | 1.13E-02 | 2.81E-02 |
| T2_FLAIR_WMHvol_Superior_longitudinal_fasciculus_R | -0.191 | 1.23E-02 | 2.84E-02 |
| IDP_dMRI_ProbtrackX_MD_slf_l | -0.258 | 4.61E-03 | 2.96E-02 |
| FS_aparc.DKTatlas_lh_area_parsopercularis. | 0.199 | 1.24E-02 | 3.05E-02 |
| IDP_dMRI_ProbtrackX_MD_unc_l | -0.253 | 5.10E-03 | 3.09E-02 |
| IDP_dMRI_ProbtrackX_OD_slf_r | 0.253 | 5.08E-03 | 3.09E-02 |
| dMRI_MD_supramargR | -0.274 | 5.25E-03 | 3.09E-02 |
| dMRI_MD_precuneousL | -0.281 | 5.42E-03 | 3.11E-02 |

#### Supplementary Table 6: Associations with diagnosis

Associations between IDPs and diagnostic groups. Corr-pval are after hierarchical FDR correction.

| Variable | coef | OR | pval | corr-pval |
|---|---|---|---|---|
| T1_GMvol_temp | -1.917 | 0.147 | 1.91E-12 | 7.56E-10 |
| T1_SIENAX_peripheral_GM_unnorm_vol_LM | -1.856 | 0.156 | 1.50E-11 | 2.96E-09 |
| T1_GM_parcellation_L_Front_Pole_vol_LM | -1.326 | 0.265 | 1.82E-09 | 2.40E-07 |
| T1_GM_parcellation_L_Parahippocampal_Gyr_ant_vol_LM | -1.221 | 0.295 | 3.94E-09 | 3.89E-07 |
| T1_SIENAX_GM_unnorm_vol_LM | -1.588 | 0.204 | 7.01E-09 | 4.81E-07 |
| T1_GM_parcellation_R_Mid_Temp_Gyr_post_vol_LM | -1.083 | 0.339 | 7.31E-09 | 4.81E-07 |
| T1_GMvol_front | -1.343 | 0.261 | 1.51E-08 | 7.48E-07 |
| T1_GMvol_par | -1.297 | 0.273 | 1.41E-08 | 7.48E-07 |
| FS_HippSubfield_lh_volume_GC.ML.DG.body. | -1.067 | 0.344 | 1.88E-08 | 8.24E-07 |
| FS_HippSubfield_lh_volume_Whole.FS_Hippocampal.body. | -1.067 | 0.344 | 2.59E-08 | 1.02E-06 |
| FS_HippSubfield_lh_volume_presubiculum.head. | -1.014 | 0.363 | 3.03E-08 | 1.09E-06 |
| FS_HippSubfield_lh_volume_CA4.body. | -1.017 | 0.362 | 4.13E-08 | 1.36E-06 |
| T1_GM_parcellation_L_Temp_Pole_vol_LM | -1.018 | 0.361 | 5.17E-08 | 1.57E-06 |
| FS_HippSubfield_lh_volume_Whole.FS_Hippocampus. | -1.011 | 0.364 | 8.32E-08 | 2.29E-06 |
| T1_GM_parcellation_R_Front_Orbital_vol_LM | -1.074 | 0.342 | 8.68E-08 | 2.29E-06 |
| FS_aparc.DKTatlas_lh_thickness_middletemporal. | -0.900 | 0.407 | 9.41E-08 | 2.32E-06 |
| FS_HippSubfield_lh_volume_Whole.FS_Hippocampal.head. | -0.960 | 0.383 | 1.85E-07 | 4.30E-06 |
| L_hipp_masked | -0.901 | 0.406 | 2.14E-07 | 4.70E-06 |
| T1_GM_parcellation_L_Insula_vol_LM | -0.989 | 0.372 | 2.53E-07 | 5.27E-06 |
| FS_HippSubfield_lh_volume_subiculum.body. | -0.960 | 0.383 | 2.68E-07 | 5.30E-06 |
| T2_FLAIR_WMHvol_Splenium_of_corpus_callosum | 0.964 | 2.621 | 1.12E-07 | 5.71E-06 |
| FS_HippSubfield_lh_volume_CA4.head. | -0.920 | 0.398 | 3.35E-07 | 6.01E-06 |
| T1_GM_parcellation_R_Temp_Pole_vol_LM | -0.961 | 0.383 | 3.20E-07 | 6.01E-06 |
| T1_GM_parcellation_R_Sup_Temp_Gyr_post_vol_LM | -0.880 | 0.415 | 4.06E-07 | 6.98E-06 |
| T1_GM_parcellation_L_Hippocampus_vol_LM | -0.972 | 0.379 | 4.63E-07 | 7.62E-06 |
| FS_HippSubfield_rh_volume_GC.ML.DG.body. | -0.932 | 0.394 | 4.85E-07 | 7.66E-06 |
| FS_HippSubfield_lh_volume_GC.ML.DG.head. | -0.891 | 0.410 | 6.07E-07 | 9.23E-06 |
| FS_HippSubfield_lh_volume_CA3.head. | -0.863 | 0.422 | 7.56E-07 | 1.11E-05 |
| T1_GM_parcellation_R_Paracingulate_Gyr_vol_LM | -0.864 | 0.421 | 9.97E-07 | 1.36E-05 |
| T1_GM_parcellation_R_Amygdala_vol_LM | -0.971 | 0.379 | 9.63E-07 | 1.36E-05 |

| | | | | |
|---|---|---|---|---|
| T1_GM_parcellation_L_Sup_Temp_Gyr_ant_vol_LM | -0.839 | 0.432 | 1.23E-06 | 1.57E-05 |
| T1_GM_parcellation_L_Mid_Temp_Gyr_post_vol_LM | -0.880 | 0.415 | 1.22E-06 | 1.57E-05 |
| T1_GM_parcellation_R_Front_Pole_vol_LM | -0.996 | 0.369 | 1.53E-06 | 1.89E-05 |
| FS_HippSubfield_rh_volume_Whole.FS_Hippocampal.body. | -0.884 | 0.413 | 1.60E-06 | 1.92E-05 |
| T1_GM_parcellation_R_Parahippocampal_Gyr_ant_vol_LM | -0.928 | 0.395 | 1.78E-06 | 2.07E-05 |
| FS_HippSubfield_rh_volume_subiculum.body. | -0.871 | 0.418 | 1.99E-06 | 2.24E-05 |
| FS_HippSubfield_lh_volume_CA3.body. | -0.784 | 0.457 | 2.26E-06 | 2.48E-05 |
| FS_HippSubfield_lh_volume_CA1.head. | -0.825 | 0.438 | 2.41E-06 | 2.57E-05 |
| T1_GM_parcellation_R_Insula_vol_LM | -0.923 | 0.397 | 2.52E-06 | 2.62E-05 |
| T1_GM_parcellation_L_Amygdala_vol_LM | -0.913 | 0.401 | 2.73E-06 | 2.77E-05 |
| FS_HippSubfield_lh_volume_subiculum.head. | -0.773 | 0.462 | 3.43E-06 | 3.39E-05 |
| FS_HippSubfield_rh_volume_CA4.body. | -0.813 | 0.443 | 3.79E-06 | 3.65E-05 |
| T1_GM_parcellation_L_Lateral_Occ_Sup_vol_LM | -0.773 | 0.462 | 4.09E-06 | 3.85E-05 |
| T1_GM_parcellation_R_Inf_Temp_Gyr_post_vol_LM | -0.819 | 0.441 | 4.55E-06 | 4.18E-05 |
| T1_GM_parcellation_L_Precuneous_vol_LM | -0.846 | 0.429 | 4.83E-06 | 4.34E-05 |
| T1_GM_parcellation_L_Mid_Temp_Gyr_ant_vol_LM | -0.783 | 0.457 | 5.11E-06 | 4.48E-05 |
| T1_GM_parcellation_L_Temp_Fusiform_ant_vol_LM | -0.790 | 0.454 | 5.64E-06 | 4.85E-05 |
| T1_GM_parcellation_L_Temp_Fusiform_post_vol_LM | -0.796 | 0.451 | 6.26E-06 | 5.26E-05 |
| FS_HippSubfield_rh_volume_Whole.FS_Hippocampus. | -0.821 | 0.440 | 7.23E-06 | 5.68E-05 |
| FS_aparc.DKTatlas_lh_thickness_inferiortemporal. | -0.722 | 0.486 | 7.11E-06 | 5.68E-05 |
| T1_GM_parcellation_L_Inf_Temp_Gyr_post_vol_LM | -0.822 | 0.440 | 7.34E-06 | 5.68E-05 |
| T1_GM_parcellation_L_Paracingulate_Gyr_vol_LM | -0.773 | 0.462 | 7.15E-06 | 5.68E-05 |
| T1_GM_parcellation_R_Precuneous_vol_LM | -0.842 | 0.431 | 7.49E-06 | 5.69E-05 |
| FS_HippSubfield_lh_volume_fimbria. | -0.778 | 0.459 | 8.03E-06 | 5.99E-05 |
| T1_GM_parcellation_L_Inf_Temp_Gyr_ant_vol_LM | -0.734 | 0.480 | 8.89E-06 | 6.50E-05 |
| FS_HippSubfield_rh_volume_CA1.head. | -0.781 | 0.458 | 9.33E-06 | 6.58E-05 |
| T1_GM_parcellation_R_Inf_Temp_Gyr_temporoOcc_part_vol_LM | -0.748 | 0.473 | 9.31E-06 | 6.58E-05 |
| R_hipp_masked | -0.720 | 0.487 | 1.27E-05 | 8.83E-05 |
| FS_aparc.DKTatlas_rh_thickness_entorhinal. | -0.700 | 0.496 | 1.59E-05 | 1.08E-04 |
| T1_GM_parcellation_L_Sup_Parietal_Lobule_vol_LM | -0.684 | 0.504 | 1.74E-05 | 1.16E-04 |
| T1_GM_parcellation_R_Inf_Temp_Gyr_ant_vol_LM | -0.707 | 0.493 | 2.04E-05 | 1.34E-04 |
| FS_aparc.DKTatlas_lh_volume_middletemporal. | -0.799 | 0.450 | 2.21E-05 | 1.43E-04 |
| FS_HippSubfield_rh_volume_Whole.FS_Hippocampal.head. | -0.754 | 0.470 | 2.41E-05 | 1.53E-04 |
| T1_GM_parcellation_R_Temp_Fusiform_ant_vol_LM | -0.778 | 0.459 | 2.60E-05 | 1.61E-04 |

| | | | | |
|---|---|---|---|---|
| T1_GM_parcellation_L_Front_Operculum_vol_LM | -0.698 | 0.497 | 2.56E-05 | 1.61E-04 |
| T1_SIENAX_CSF_unnorm_vol_LM | 0.793 | 2.211 | 2.69E-05 | 1.64E-04 |
| WMH_pvent | 0.769 | 2.157 | 6.76E-06 | 1.72E-04 |
| FS_aparc.DKTatlas_lh_volume_superiortemporal. | -0.750 | 0.472 | 2.88E-05 | 1.72E-04 |
| FS_HippSubfield_rh_volume_fimbria. | -0.714 | 0.490 | 3.13E-05 | 1.79E-04 |
| FS_aparc.DKTatlas_lh_thickness_entorhinal. | -0.679 | 0.507 | 3.09E-05 | 1.79E-04 |
| T1_GM_parcellation_R_Hippocampus_vol_LM | -0.765 | 0.466 | 3.13E-05 | 1.79E-04 |
| FS_HippSubfield_rh_volume_CA3.body. | -0.660 | 0.517 | 3.52E-05 | 1.98E-04 |
| FS_aparc.DKTatlas_lh_thickness_superiortemporal. | -0.683 | 0.505 | 3.80E-05 | 2.11E-04 |
| T2_FLAIR_WMHvol_Body_of_corpus_callosum | 0.742 | 2.100 | 1.30E-05 | 2.22E-04 |
| FS_aparc.DKTatlas_lh_volume_inferiortemporal. | -0.703 | 0.495 | 4.22E-05 | 2.32E-04 |
| T1_GM_parcellation_R_Mid_Temp_Gyr_ant_vol_LM | -0.685 | 0.504 | 4.37E-05 | 2.36E-04 |
| FS_HippSubfield_rh_volume_CA3.head. | -0.688 | 0.503 | 4.82E-05 | 2.57E-04 |
| T1_GM_parcellation_L_Front_Orbital_vol_LM | -0.752 | 0.471 | 4.89E-05 | 2.58E-04 |
| rfMRI_d100_NodeAmplitudes_28 | -1.250 | 0.286 | 9.24E-08 | 3.20E-04 |
| T1_GM_parcellation_R_Central_Opercular_vol_LM | -0.741 | 0.476 | 6.33E-05 | 3.29E-04 |
| FS_aparc.DKTatlas_lh_thickness_fusiform. | -0.632 | 0.532 | 7.04E-05 | 3.56E-04 |
| FS_aparc.DKTatlas_rh_volume_middletemporal. | -0.719 | 0.487 | 7.04E-05 | 3.56E-04 |
| T1_GM_parcellation_R_Caudate_vol_LM | -0.754 | 0.471 | 7.66E-05 | 3.83E-04 |
| FS_HippSubfield_rh_volume_GC.ML.DG.head. | -0.657 | 0.518 | 9.77E-05 | 4.83E-04 |
| FS_aparc.DKTatlas_rh_thickness_superiortemporal. | -0.648 | 0.523 | 1.01E-04 | 4.86E-04 |
| T1_GM_parcellation_L_Sup_Front_Gyr_vol_LM | -0.650 | 0.522 | 9.99E-05 | 4.86E-04 |
| T1_GM_parcellation_L_Sup_Temp_Gyr_post_vol_LM | -0.605 | 0.546 | 1.10E-04 | 5.05E-04 |
| T1_GM_parcellation_R_Subcallosal_vol_LM | -0.776 | 0.460 | 1.09E-04 | 5.05E-04 |
| T1_GM_parcellation_L_Central_Opercular_vol_LM | -0.738 | 0.478 | 1.09E-04 | 5.05E-04 |
| T1_GM_parcellation_R_Pallidum_vol_LM | 0.620 | 1.860 | 1.07E-04 | 5.05E-04 |
| IDP_dMRI_ProbtrackX_MD_cgh_r | 1.076 | 2.934 | 5.86E-06 | 5.08E-04 |
| dMRI_MD_precuneousR | 1.439 | 4.216 | 6.99E-06 | 5.08E-04 |
| dMRI_PSMD | 1.157 | 3.182 | 4.12E-06 | 5.08E-04 |
| FS_aparc.DKTatlas_rh_volume_medialorbitofrontal. | -0.677 | 0.508 | 1.14E-04 | 5.18E-04 |
| FS_aparc.DKTatlas_rh_volume_superiortemporal. | -0.709 | 0.492 | 1.28E-04 | 5.74E-04 |
| FS_HippSubfield_rh_volume_CA4.head. | -0.637 | 0.529 | 1.30E-04 | 5.78E-04 |
| FS_HippSubfield_lh_volume_presubiculum.body. | -0.603 | 0.547 | 1.36E-04 | 5.96E-04 |
| WMH_total | 0.685 | 1.984 | 4.80E-05 | 6.12E-04 |
| T1_GM_parcellation_L_Inf_Temp_Gyr_temporoOcc_part_vol_LM | -0.622 | 0.537 | 1.42E-04 | 6.15E-04 |
| T1_GM_parcellation_R_Temp_Fusiform_post_vol_LM | -0.662 | 0.516 | 1.43E-04 | 6.15E-04 |
| rfMRI_d100_NodeAmplitudes_44 | -1.182 | 0.307 | 6.56E-07 | 7.58E-04 |
| rfMRI_d25_NodeAmplitudes_6 | -1.088 | 0.337 | 6.23E-07 | 7.58E-04 |

| | | | | |
|---|---|---|---|---|
| FS_aparc.DKTatlas_lh_volume_entorhinal. | -0.588 | 0.556 | 2.08E-04 | 8.70E-04 |
| T1_GM_parcellation_L_Inf_Front_Gyr_pars_triangularis_vol_LM | -0.560 | 0.571 | 2.11E-04 | 8.70E-04 |
| T1_GM_parcellation_R_Sup_Temp_Gyr_ant_vol_LM | -0.636 | 0.530 | 2.10E-04 | 8.70E-04 |
| T1_GM_parcellation_R_Postcentral_Gyr_vol_LM | -0.639 | 0.528 | 2.07E-04 | 8.70E-04 |
| T1_GM_parcellation_L_Supramarginal_Gyr_post_vol_LM | -0.602 | 0.548 | 2.24E-04 | 8.93E-04 |
| T1_GM_parcellation_R_Lateral_Occ_Sup_vol_LM | -0.612 | 0.542 | 2.24E-04 | 8.93E-04 |
| T1_GM_parcellation_R_Planum_Polare_vol_LM | -0.722 | 0.486 | 2.22E-04 | 8.93E-04 |
| T2_FLAIR_WMHvol_Posterior_thalamic_radiation_L | 0.625 | 1.869 | 9.15E-05 | 9.33E-04 |
| IDP_T1_FIRST_left_accumbens_volume | -0.588 | 0.555 | 2.44E-04 | 9.63E-04 |
| T2_FLAIR_WMHvol_Superior_fronto.occipital_fasciculus_R | 0.710 | 2.035 | 1.18E-04 | 1.00E-03 |
| IDP_T1_FIRST_right_accumbens_volume | -0.577 | 0.561 | 2.65E-04 | 1.03E-03 |
| T1_GM_parcellation_L_Subcallosal_vol_LM | -0.713 | 0.490 | 2.67E-04 | 1.03E-03 |
| T1_GMvol_occ | -0.722 | 0.486 | 2.99E-04 | 1.15E-03 |
| FS_HippSubfield_lh_volume_CA1.body. | -0.556 | 0.573 | 3.24E-04 | 1.23E-03 |
| T1_GM_parcellation_R_Angular_Gyr_vol_LM | -0.552 | 0.576 | 3.28E-04 | 1.23E-03 |
| T1_GM_parcellation_R_Planum_Tempe_vol_LM | -0.646 | 0.524 | 3.31E-04 | 1.23E-03 |
| T1_GM_parcellation_L_Front_Medial_vol_LM | -0.573 | 0.564 | 3.73E-04 | 1.38E-03 |
| rfMRI_d100_NodeAmplitudes_21 | -1.036 | 0.355 | 2.19E-06 | 1.52E-03 |
| rfMRI_d25_NodeAmplitudes_13 | -1.049 | 0.350 | 1.92E-06 | 1.52E-03 |
| T1_GM_parcellation_L_Lateral_Occ_Inf_vol_LM | -0.568 | 0.567 | 4.18E-04 | 1.53E-03 |
| FS_aparc.DKTatlas_rh_volume_inferiorparietal. | -0.618 | 0.539 | 4.50E-04 | 1.63E-03 |
| T1_GM_parcellation_L_Caudate_vol_LM | -0.653 | 0.521 | 5.05E-04 | 1.81E-03 |
| rfMRI_d100_NodeAmplitudes_31 | -0.980 | 0.375 | 3.24E-06 | 1.87E-03 |
| FS_HippSubfield_lh_volume_FS_Hippocampal.tail. | -0.545 | 0.580 | 5.49E-04 | 1.94E-03 |
| T1_GM_parcellation_R_Front_Operculum_vol_LM | -0.562 | 0.570 | 5.47E-04 | 1.94E-03 |
| FS_HippSubfield_rh_volume_presubiculum.head. | -0.590 | 0.554 | 5.72E-04 | 2.00E-03 |
| T1_GM_parcellation_L_Parietal_Operculum_vol_LM | -0.537 | 0.584 | 5.98E-04 | 2.07E-03 |
| T2_FLAIR_WMHvol_Anterior_corona_radiata_R | 0.593 | 1.810 | 2.89E-04 | 2.10E-03 |
| FS_aparc.DKTatlas_lh_volume_insula. | -0.610 | 0.544 | 6.42E-04 | 2.21E-03 |
| FS_HippSubfield_lh_volume_molecular.layer.HP.head. | -0.556 | 0.573 | 6.83E-04 | 2.33E-03 |
| FS_HippSubfield_rh_volume_FS_Hippocampal.tail. | -0.526 | 0.591 | 6.89E-04 | 2.33E-03 |
| T1_GM_parcellation_L_Supramarginal_Gyr_ant_vol_LM | -0.535 | 0.586 | 7.07E-04 | 2.37E-03 |
| FS_aparc.DKTatlas_lh_volume_superiorparietal. | -0.584 | 0.558 | 7.38E-04 | 2.45E-03 |
| FS_HippSubfield_rh_volume_CA1.body. | -0.513 | 0.599 | 7.82E-04 | 2.57E-03 |
| rfMRI_d100_NodeAmplitudes_25 | -0.928 | 0.395 | 5.22E-06 | 2.59E-03 |

| | | | | |
|---|---|---|---|---|
| FS_aparc.DKTatlas_lh_thickness_medialorbitofrontal. | -0.510 | 0.600 | 7.97E-04 | 2.59E-03 |
| FS_aparc.DKTatlas_rh_thickness_middletemporal. | -0.519 | 0.595 | 7.99E-04 | 2.59E-03 |
| FS_aparc.DKTatlas_lh_thickness_paraFS_Hippocampal. | -0.504 | 0.604 | 8.56E-04 | 2.75E-03 |
| IDP_dMRI_ProbtrackX_ISOVF_cgh_r | 0.925 | 2.523 | 5.19E-05 | 2.83E-03 |
| IDP_T1_FIRST_left_putamen_volume | -0.552 | 0.576 | 9.30E-04 | 2.96E-03 |
| T1_GM_parcellation_R_Supramarginal_Gyr_post_vol_LM | -0.532 | 0.588 | 9.73E-04 | 3.07E-03 |
| FS_aparc.DKTatlas_lh_volume_paraFS_Hippocampal. | -0.488 | 0.614 | 9.99E-04 | 3.13E-03 |
| rfMRI_d100_NodeAmplitudes_7 | -0.922 | 0.398 | 8.30E-06 | 3.20E-03 |
| rfMRI_d100_NodeAmplitudes_45 | -0.957 | 0.384 | 7.91E-06 | 3.20E-03 |
| rfMRI_d100_partialcorr_442 | 0.934 | 2.546 | 9.37E-06 | 3.25E-03 |
| T1_GM_parcellation_L_Planum_Polare_vol_LM | -0.588 | 0.555 | 1.05E-03 | 3.26E-03 |
| IDP_dMRI_ProbtrackX_MD_cgh_l | 0.771 | 2.162 | 9.40E-05 | 3.42E-03 |
| IDP_dMRI_ProbtrackX_MO_fmi | -0.792 | 0.453 | 8.01E-05 | 3.42E-03 |
| T2_FLAIR_WMHvol_Genu_of_corpus_callosum | 0.552 | 1.737 | 5.77E-04 | 3.46E-03 |
| T2_FLAIR_WMHvol_Superior_corona_radiata_R | 0.585 | 1.795 | 6.79E-04 | 3.46E-03 |
| T2_FLAIR_WMHvol_Superior_corona_radiata_L | 0.554 | 1.740 | 6.34E-04 | 3.46E-03 |
| IDP_T1_FIRST_right_putamen_volume | -0.532 | 0.588 | 1.15E-03 | 3.54E-03 |
| FS_aparc.DKTatlas_lh_thickness_insula. | -0.486 | 0.615 | 1.19E-03 | 3.64E-03 |
| T1_GM_parcellation_L_Mid_Temp_Gyr_temporoOcc_part_vol_LM | -0.486 | 0.615 | 1.32E-03 | 4.01E-03 |
| T1_SIENAX_brain.unnorm_vol_LM | -0.948 | 0.388 | 1.36E-03 | 4.10E-03 |
| FS_aparc.DKTatlas_rh_thickness_insula. | -0.493 | 0.611 | 1.37E-03 | 4.11E-03 |
| IDP_dMRI_ProbtrackX_MD_ptr_l | 0.793 | 2.210 | 1.39E-04 | 4.34E-03 |
| T2_FLAIR_WMHvol_Posterior_thalamic_radiation_R | 0.535 | 1.708 | 9.69E-04 | 4.35E-03 |
| T2_FLAIR_WMHvol_Cingulum_cingulate_gyrus_L | 0.674 | 1.962 | 1.02E-03 | 4.35E-03 |
| FS_aparc.DKTatlas_lh_thickness_pericalcarine. | 0.458 | 1.580 | 1.47E-03 | 4.36E-03 |
| FS_aparc.DKTatlas_rh_volume_entorhinal. | -0.524 | 0.592 | 1.49E-03 | 4.39E-03 |
| FS_aparc.DKTatlas_rh_thickness_medialorbitofrontal. | -0.500 | 0.607 | 1.52E-03 | 4.44E-03 |
| T1_GM_parcellation_R_Sup_Parietal_Lobule_vol_LM | -0.471 | 0.624 | 1.69E-03 | 4.92E-03 |
| T1_GM_parcellation_R_Sup_Front_Gyr_vol_LM | -0.529 | 0.589 | 1.77E-03 | 5.11E-03 |
| FS_aparc.DKTatlas_lh_volume_fusiform. | -0.542 | 0.581 | 1.85E-03 | 5.28E-03 |
| T2_FLAIR_WMHvol_External_capsule_R | 0.533 | 1.704 | 1.62E-03 | 6.36E-03 |
| T1_GM_parcellation_R_Mid_Temp_Gyr_temporoOcc_part_vol_LM | -0.483 | 0.617 | 2.26E-03 | 6.43E-03 |
| T1_GM_parcellation_R_Front_Medial_vol_LM | -0.486 | 0.615 | 2.28E-03 | 6.43E-03 |
| rfMRI_d100_NodeAmplitudes_19 | -0.895 | 0.409 | 2.05E-05 | 6.47E-03 |
| dMRI_MO_supramargR | -0.832 | 0.435 | 2.57E-04 | 7.02E-03 |

| | | | | |
|---|---|---|---|---|
| FS_HippSubfield_lh_volume_molecular.layer.HP.body. | -0.453 | 0.636 | 2.61E-03 | 7.31E-03 |
| rfMRI_d100_NodeAmplitudes_37 | -0.858 | 0.424 | 2.75E-05 | 7.94E-03 |
| FS_aparc.DKTatlas_rh_thickness_paraFS_Hippocampal. | -0.450 | 0.638 | 2.88E-03 | 8.02E-03 |
| FS_aparc.DKTatlas_lh_volume_medialorbitofrontal. | -0.506 | 0.603 | 2.91E-03 | 8.04E-03 |
| FS_aparc.DKTatlas_lh_thickness_inferiorparietal. | -0.445 | 0.641 | 3.01E-03 | 8.24E-03 |
| T1_GM_parcellation_L_Pallidum_vol_LM | 0.456 | 1.577 | 3.04E-03 | 8.27E-03 |
| T2_FLAIR_WMHvol_Retrolenticular_part_of_internal_capsule_R | 0.458 | 1.581 | 2.27E-03 | 8.28E-03 |
| FS_HippSubfield_rh_volume_subiculum.head. | -0.486 | 0.615 | 3.17E-03 | 8.51E-03 |
| T1_GM_parcellation_R_Inf_Front_Gyr_pars_triangularis_vol_LM | -0.446 | 0.640 | 3.15E-03 | 8.51E-03 |
| IDP_dMRI_ProbtrackX_ISOVF_unc_r | 0.726 | 2.067 | 3.53E-04 | 8.56E-03 |
| FS_aparc.DKTatlas_rh_volume_inferiortemporal. | -0.507 | 0.603 | 3.32E-03 | 8.87E-03 |
| FS_HippSubfield_lh_volume_HATA. | -0.447 | 0.640 | 3.37E-03 | 8.87E-03 |
| FS_aparc.DKTatlas_rh_area_middletemporal. | -0.547 | 0.579 | 3.36E-03 | 8.87E-03 |
| FS_aparc.DKTatlas_lh_volume_superiorfrontal. | -0.518 | 0.596 | 3.56E-03 | 9.28E-03 |
| FS_aparc.DKTatlas_rh_volume_insula. | -0.513 | 0.599 | 3.57E-03 | 9.28E-03 |
| IDP_dMRI_ProbtrackX_ISOVF_cgh_l | 0.662 | 1.938 | 4.68E-04 | 9.28E-03 |
| dMRI_MO_R_amyg | -0.771 | 0.462 | 4.39E-04 | 9.28E-03 |
| T1_GM_parcellation_L_Inf_Front_Gyr_pars_opercularis_vol_LM | -0.457 | 0.633 | 3.66E-03 | 9.44E-03 |
| T2_FLAIR_WMHvol_Posterior_corona_radiata_L | 0.460 | 1.584 | 2.88E-03 | 9.80E-03 |
| T2_FLAIR_WMHvol_Superior_longitudinal_fasciculus_L | 0.473 | 1.604 | 3.15E-03 | 1.00E-02 |
| IDP_dMRI_ProbtrackX_ICVF_fma | -0.672 | 0.510 | 5.57E-04 | 1.01E-02 |
| FS_aparc.DKTatlas_lh_volume_parstriangularis. | -0.457 | 0.633 | 4.21E-03 | 1.08E-02 |
| T1_GM_parcellation_L_Occ_Pole_vol_LM | -0.456 | 0.634 | 4.23E-03 | 1.08E-02 |
| FS_aparc.DKTatlas_rh_thickness_inferiortemporal. | -0.448 | 0.639 | 4.33E-03 | 1.09E-02 |
| FS_aparc.DKTatlas_rh_volume_fusiform. | -0.480 | 0.619 | 4.34E-03 | 1.09E-02 |
| rfMRI_d100_NodeAmplitudes_13 | -0.845 | 0.429 | 4.19E-05 | 1.12E-02 |
| IDP_T1_FIRST_right_amygdala_volume | -0.420 | 0.657 | 4.84E-03 | 1.19E-02 |
| FS_aparc.DKTatlas_lh_area_superiorparietal. | -0.488 | 0.614 | 4.91E-03 | 1.19E-02 |
| FS_aparc.DKTatlas_rh_thickness_pericalcarine. | 0.413 | 1.512 | 4.86E-03 | 1.19E-02 |
| T1_GM_parcellation_R_Mid_Front_Gyr_vol_LM | -0.488 | 0.614 | 4.82E-03 | 1.19E-02 |
| T1_GM_parcellation_R_Parahippocampal_Gyr_post_vol_LM | -0.444 | 0.641 | 4.88E-03 | 1.19E-02 |
| T1_NBMvol_L | -0.462 | 0.630 | 4.79E-03 | 1.19E-02 |
| T1_NBMvol_R | -0.440 | 0.644 | 4.96E-03 | 1.20E-02 |

| | | | | |
|---|---|---|---|---|
| T2_FLAIR_WMHvol_Posterior_limb_of_internal_capsule_R | 0.559 | 1.749 | 4.37E-03 | 1.24E-02 |
| T2_FLAIR_WMHvol_Tapetum_L | 0.424 | 1.528 | 4.13E-03 | 1.24E-02 |
| IDP_dMRI_ProbtrackX_ISOVF_ptr_l | 0.671 | 1.957 | 7.51E-04 | 1.26E-02 |
| T1_GM_parcellation_L_Parahippocampal_Gyr_post_vol_LM | -0.439 | 0.645 | 5.33E-03 | 1.28E-02 |
| T2_FLAIR_WMHvol_Retrolenticular_part_of_internal_capsule_L | 0.505 | 1.657 | 4.76E-03 | 1.28E-02 |
| T2_FLAIR_WMHvol_Posterior_corona_radiata_R | 0.422 | 1.525 | 5.68E-03 | 1.45E-02 |
| IDP_dMRI_ProbtrackX_MD_atr_r | 0.668 | 1.951 | 1.34E-03 | 1.46E-02 |
| IDP_dMRI_ProbtrackX_MD_ifo_l | 0.689 | 1.991 | 1.02E-03 | 1.46E-02 |
| IDP_dMRI_ProbtrackX_MD_ilf_l | 0.652 | 1.919 | 1.05E-03 | 1.46E-02 |
| IDP_dMRI_ProbtrackX_ICVF_ilf_l | -0.631 | 0.532 | 1.25E-03 | 1.46E-02 |
| IDP_dMRI_ProbtrackX_ICVF_ptr_l | -0.626 | 0.534 | 1.22E-03 | 1.46E-02 |
| IDP_dMRI_ProbtrackX_ICVF_ptr_r | -0.641 | 0.527 | 1.33E-03 | 1.46E-02 |
| dMRI_tractvol_fma | -0.814 | 0.443 | 1.12E-03 | 1.46E-02 |
| FS_aparc.DKTatlas_rh_volume_paraFS_Hippocampal. | -0.422 | 0.656 | 6.27E-03 | 1.49E-02 |
| FS_aparc.DKTatlas_rh_volume_superiorfrontal. | -0.535 | 0.586 | 6.36E-03 | 1.50E-02 |
| T1_GM_parcellation_L_Precentral_Gyr_vol_LM | -0.461 | 0.631 | 6.53E-03 | 1.54E-02 |
| IDP_dMRI_ProbtrackX_MD_ilf_r | 0.630 | 1.878 | 1.57E-03 | 1.56E-02 |
| IDP_dMRI_ProbtrackX_ICVF_ilf_r | -0.617 | 0.540 | 1.57E-03 | 1.56E-02 |
| IDP_dMRI_ProbtrackX_MD_str_r | 0.635 | 1.886 | 1.66E-03 | 1.57E-02 |
| T1_GM_parcellation_R_Inf_Front_Gyr_pars_opercularis_vol_LM | -0.406 | 0.666 | 7.16E-03 | 1.67E-02 |
| IDP_dMRI_ProbtrackX_MD_ifo_r | 0.635 | 1.886 | 1.93E-03 | 1.69E-02 |
| IDP_dMRI_ProbtrackX_ICVF_cgh_r | -0.634 | 0.531 | 1.89E-03 | 1.69E-02 |
| FS_aparc.DKTatlas_lh_area_inferiortemporal. | -0.471 | 0.625 | 7.35E-03 | 1.71E-02 |
| IDP_dMRI_ProbtrackX_OD_str_l | -0.588 | 0.556 | 2.05E-03 | 1.72E-02 |
| IDP_dMRI_ProbtrackX_FA_cgh_l | -0.595 | 0.552 | 2.36E-03 | 1.72E-02 |
| IDP_dMRI_ProbtrackX_FA_ptr_l | -0.581 | 0.560 | 2.53E-03 | 1.72E-02 |
| IDP_dMRI_ProbtrackX_MD_ptr_r | 0.598 | 1.819 | 2.24E-03 | 1.72E-02 |
| IDP_dMRI_ProbtrackX_MD_slf_l | 0.586 | 1.797 | 2.76E-03 | 1.72E-02 |
| IDP_dMRI_ProbtrackX_MD_slf_r | 0.593 | 1.809 | 2.63E-03 | 1.72E-02 |
| IDP_dMRI_ProbtrackX_ICVF_slf_r | -0.580 | 0.560 | 2.71E-03 | 1.72E-02 |
| IDP_dMRI_ProbtrackX_OD_str_r | -0.573 | 0.564 | 2.36E-03 | 1.72E-02 |
| dMRI_MD_parahippL | 0.681 | 1.977 | 2.58E-03 | 1.72E-02 |
| dMRI_tractvol_cst_l | 0.714 | 2.041 | 2.54E-03 | 1.72E-02 |
| T2_FLAIR_WMHvol_Tapetum_R | 0.397 | 1.487 | 7.23E-03 | 1.76E-02 |

| | | | | |
|---|---|---|---|---|
| FS_HippSubfield_rh_volume_molecular.layer.HP.head. | -0.411 | 0.663 | 7.92E-03 | 1.82E-02 |
| T1_GM_parcellation_Brain.Stem_vol_LM | 0.420 | 1.521 | 7.88E-03 | 1.82E-02 |
| rfMRI_d100_NodeAmplitudes_11 | -0.773 | 0.462 | 9.61E-05 | 1.83E-02 |
| rfMRI_d100_partialcorr_238 | 0.797 | 2.219 | 8.73E-05 | 1.83E-02 |
| rfMRI_d100_partialcorr_360 | -0.828 | 0.437 | 7.46E-05 | 1.83E-02 |
| rfMRI_d25_NodeAmplitudes_1 | -0.752 | 0.471 | 1.00E-04 | 1.83E-02 |
| rfMRI_d25_NodeAmplitudes_14 | -0.786 | 0.456 | 9.36E-05 | 1.83E-02 |
| rfMRI_d25_fullcorr_105 | 0.761 | 2.141 | 9.86E-05 | 1.83E-02 |
| IDP_dMRI_ProbtrackX_OD_slf_l | -0.569 | 0.566 | 3.17E-03 | 1.92E-02 |
| FS_aparc.DKTatlas_lh_volume_parsopercularis. | -0.430 | 0.651 | 8.70E-03 | 1.99E-02 |
| dMRI_MD_precuneousL | 0.671 | 1.957 | 3.37E-03 | 1.99E-02 |
| IDP_dMRI_ProbtrackX_ICVF_slf_l | -0.559 | 0.572 | 3.56E-03 | 2.04E-02 |
| IDP_T1_FIRST_left_amygdala_volume | -0.372 | 0.689 | 9.16E-03 | 2.08E-02 |
| IDP_dMRI_ProbtrackX_MD_unc_r | 0.570 | 1.768 | 3.81E-03 | 2.12E-02 |
| IDP_dMRI_ProbtrackX_ISOVF_unc_l | 0.571 | 1.770 | 3.89E-03 | 2.12E-02 |
| dMRI_tractvol_ml_l | 0.744 | 2.104 | 4.07E-03 | 2.16E-02 |
| IDP_dMRI_ProbtrackX_MD_cst_r | 0.543 | 1.721 | 4.37E-03 | 2.27E-02 |
| T2_FLAIR_WMHvol_Anterior_limb_of_internal_capsule_R | 0.444 | 1.559 | 1.05E-02 | 2.28E-02 |
| T2_FLAIR_WMHvol_Posterior_limb_of_internal_capsule_L | 0.489 | 1.631 | 1.04E-02 | 2.28E-02 |
| T2_FLAIR_WMHvol_Superior_fronto.occipital_fasciculus_L | 0.415 | 1.515 | 1.07E-02 | 2.28E-02 |
| dMRI_MD_R_amyg | 0.652 | 1.919 | 4.59E-03 | 2.33E-02 |
| FS_aparc.DKTatlas_lh_thickness_supramarginal. | -0.391 | 0.676 | 1.07E-02 | 2.41E-02 |
| T2_FLAIR_WMHvol_Anterior_corona_radiata_L | 0.394 | 1.483 | 1.19E-02 | 2.43E-02 |
| IDP_dMRI_ProbtrackX_ICVF_cgh_l | -0.554 | 0.574 | 5.07E-03 | 2.51E-02 |
| IDP_dMRI_ProbtrackX_MO_cgh_l | -0.551 | 0.577 | 5.44E-03 | 2.60E-02 |
| IDP_dMRI_ProbtrackX_ICVF_ifo_l | -0.542 | 0.582 | 5.61E-03 | 2.60E-02 |
| IDP_dMRI_ProbtrackX_ICVF_ifo_r | -0.554 | 0.575 | 5.56E-03 | 2.60E-02 |
| T1_GM_parcellation_R_Parietal_Operculum_vol_LM | -0.412 | 0.662 | 1.18E-02 | 2.65E-02 |
| rfMRI_d100_partialcorr_961 | -0.755 | 0.470 | 1.55E-04 | 2.68E-02 |
| FS_aparc.DKTatlas_rh_area_fusiform. | -0.457 | 0.633 | 1.22E-02 | 2.70E-02 |
| T1_GM_parcellation_R_Occ_Fusiform_Gyr_vol_LM | -0.396 | 0.673 | 1.22E-02 | 2.70E-02 |
| IDP_dMRI_ProbtrackX_ICVF_ar_r | -0.513 | 0.599 | 5.99E-03 | 2.72E-02 |
| rfMRI_d100_NodeAmplitudes_24 | -0.740 | 0.477 | 1.84E-04 | 3.03E-02 |
| rfMRI_d100_NodeAmplitudes_12 | -0.757 | 0.469 | 2.02E-04 | 3.04E-02 |
| rfMRI_d100_fullcorr_442 | 0.697 | 2.007 | 1.94E-04 | 3.04E-02 |

| | | | | |
|---|---|---|---|---|
| T1_GM_parcellation_R_Heschl_Gyr_vol_LM | -0.407 | 0.666 | 1.38E-02 | 3.05E-02 |
| rfMRI_d100_fullcorr_135 | 0.742 | 2.100 | 2.23E-04 | 3.21E-02 |
| IDP_dMRI_ProbtrackX_FA_fmi | -0.527 | 0.591 | 7.33E-03 | 3.23E-02 |
| IDP_dMRI_ProbtrackX_ISOVF_fmi | 0.491 | 1.634 | 7.56E-03 | 3.23E-02 |
| dMRI_MO_supramargL | -0.570 | 0.565 | 7.57E-03 | 3.23E-02 |
| rfMRI_d100_fullcorr_1095 | -0.729 | 0.483 | 2.35E-04 | 3.26E-02 |
| IDP_dMRI_ProbtrackX_MD_unc_l | 0.555 | 1.742 | 7.96E-03 | 3.27E-02 |
| IDP_dMRI_ProbtrackX_OD_ar_l | -0.496 | 0.609 | 7.92E-03 | 3.27E-02 |
| FS_aparc.DKTatlas_rh_thickness_isthmuscingulate. | -0.363 | 0.696 | 1.51E-02 | 3.30E-02 |
| FS_aparc.DKTatlas_rh_area_inferiorparietal. | -0.439 | 0.644 | 1.52E-02 | 3.32E-02 |
| IDP_dMRI_ProbtrackX_MO_atr_r | -0.484 | 0.617 | 8.23E-03 | 3.32E-02 |

#### Supplementary Figure 1: T2* IDPs

Comparison of T2* IDPs derived from the scanner-combined and the separate coil images, showing their equivalence. IDPs from one scan (in green) differed and were excluded from further analysis.

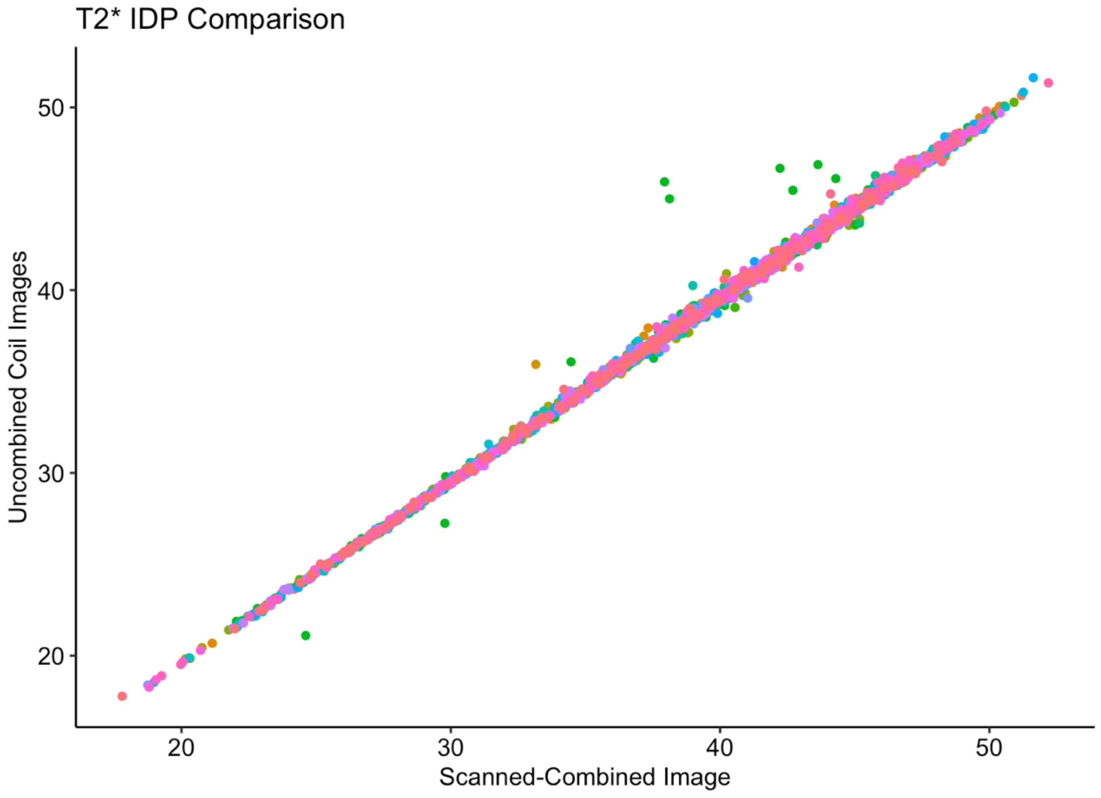

#### Supplementary Figure 2: P-values of associations with age without ACE-III adjustment

FDR-corrected p-values for associations between IDPs and age without covarying for ACE-III total cognitive score. Each dot represents one IDP, grouped by analysis tool/method and colour-coded by scan modality if significant. SIENAX, Structural Image Evaluation using Normalization of Atrophy (cross-sectionally); FIRST, FMRIB's Integrated Registration and Segmentation Tool; VBM, voxel-based morphometry; BIANCA, Brain Intensity AbNormality Classification Algorithm; FA, fractional anisotropy; MD, mean diffusivity; MO, mode of anisotropy; OD, orientation dispersion index; ICVF, intra-cellular volume fraction; ISOVF, isotropic volume fraction; CBF, cerebral blood flow; NBM, nucleus basalis of Meynert; WMH, white matter hyperintensity; PSMD, peak width of skeletonised mean diffusivity.

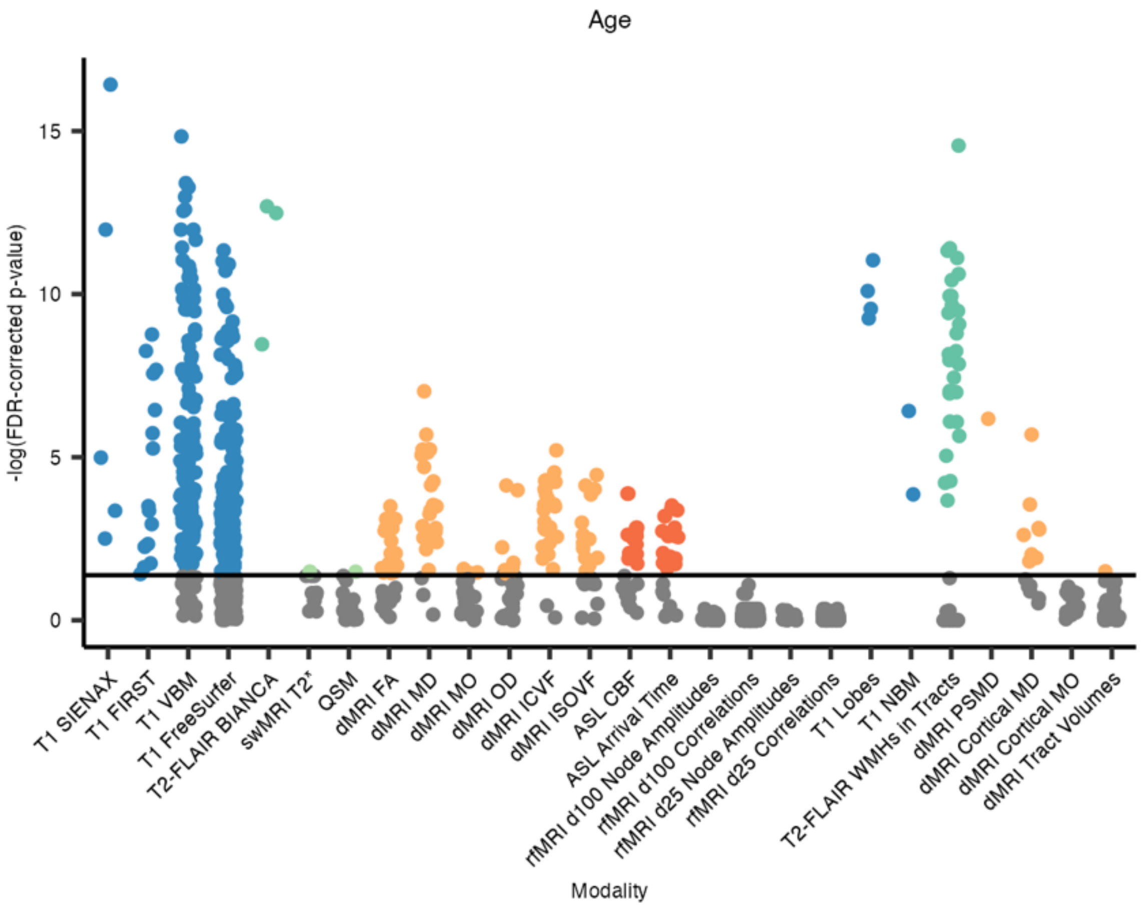

#### Supplementary Figure 3: Beta coefficients of associations with age without ACE-III adjustment

Beta coefficients for associations between IDPs and age without covarying for ACE-III total cognitive score. All numeric variables are unit standardised, meaning that 1 standard deviation (SD) increase in an IDP value is associated with a β SD difference in age. Coloured dots indicate associations significant at the 5% FDR level, hierarchical by modality. SIENAX, Structural Image Evaluation using Normalization of Atrophy (cross-sectionally); FIRST, FMRIB's Integrated Registration and Segmentation Tool; VBM, voxel-based morphometry; BIANCA, Brain Intensity AbNormality Classification Algorithm; FA, fractional anisotropy; MD, mean diffusivity; MO, mode of anisotropy; OD, orientation dispersion index; ICVF, intra-cellular volume fraction; ISOVF, isotropic volume fraction; CBF, cerebral blood flow; NBM, nucleus basalis of Meynert; WMH, white matter hyperintensity; PSMD, peak width of skeletonised mean diffusivity.

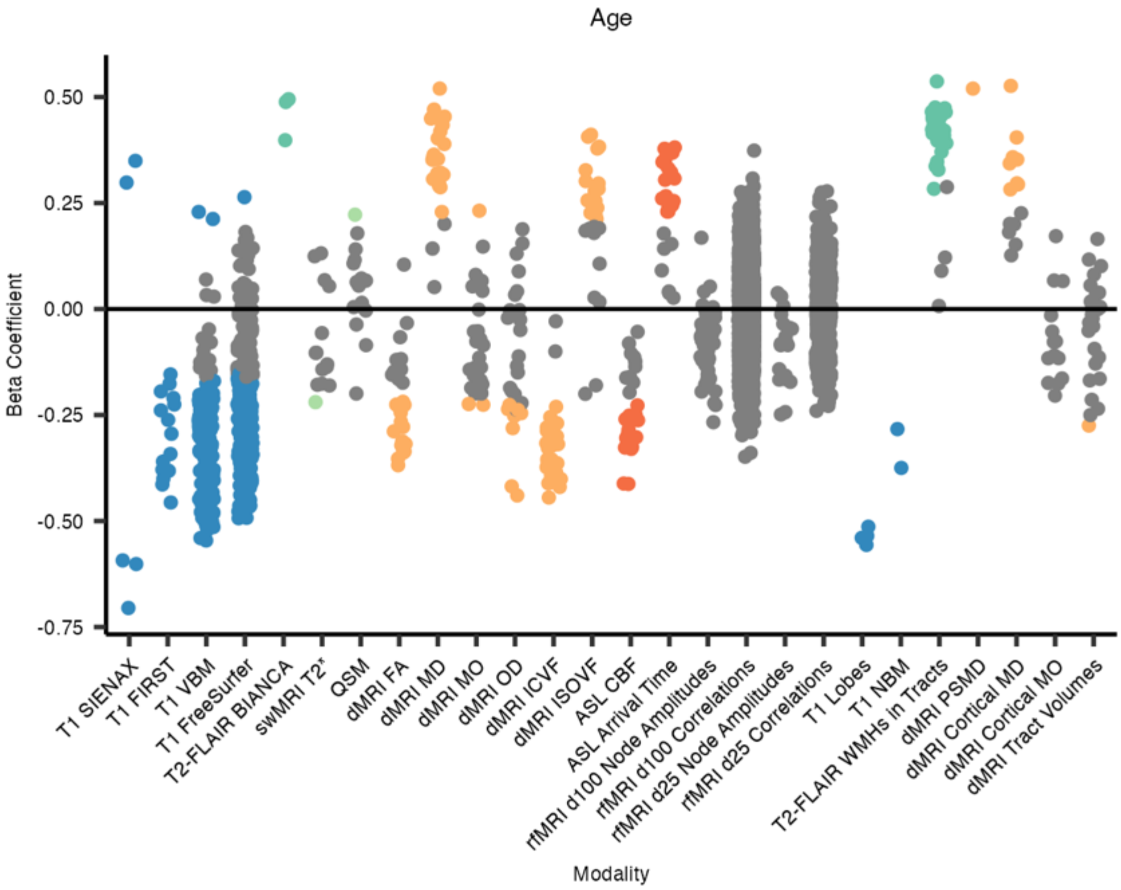

#### Supplementary Figure 4: P-values of associations with age controlling for volume

FDR-corrected p-values for associations between IDPs and age, controlling for ROI volume with T2* and QSM, and tract volume with tractography-based IDPs (dMRI FA, MD, MO, OD, ICVF, ISOVF). Because the swMRI/QSM modality is no longer significant, the FDR-adjusted threshold (solid line) drops from 0.05(5/6) to 0.05(4/6). SIENAX, Structural Image Evaluation using Normalization of Atrophy (cross-sectionally); FIRST, FMRIB's Integrated Registration and Segmentation Tool; VBM, voxel-based morphometry; BIANCA, Brain Intensity AbNormality Classification Algorithm; FA, fractional anisotropy; MD, mean diffusivity; MO, mode of anisotropy; OD, orientation dispersion index; ICVF, intra-cellular volume fraction; ISOVF, isotropic volume fraction; CBF, cerebral blood flow; NBM, nucleus basalis of Meynert; WMH, white matter hyperintensity; PSMD, peak width of skeletonised mean diffusivity.

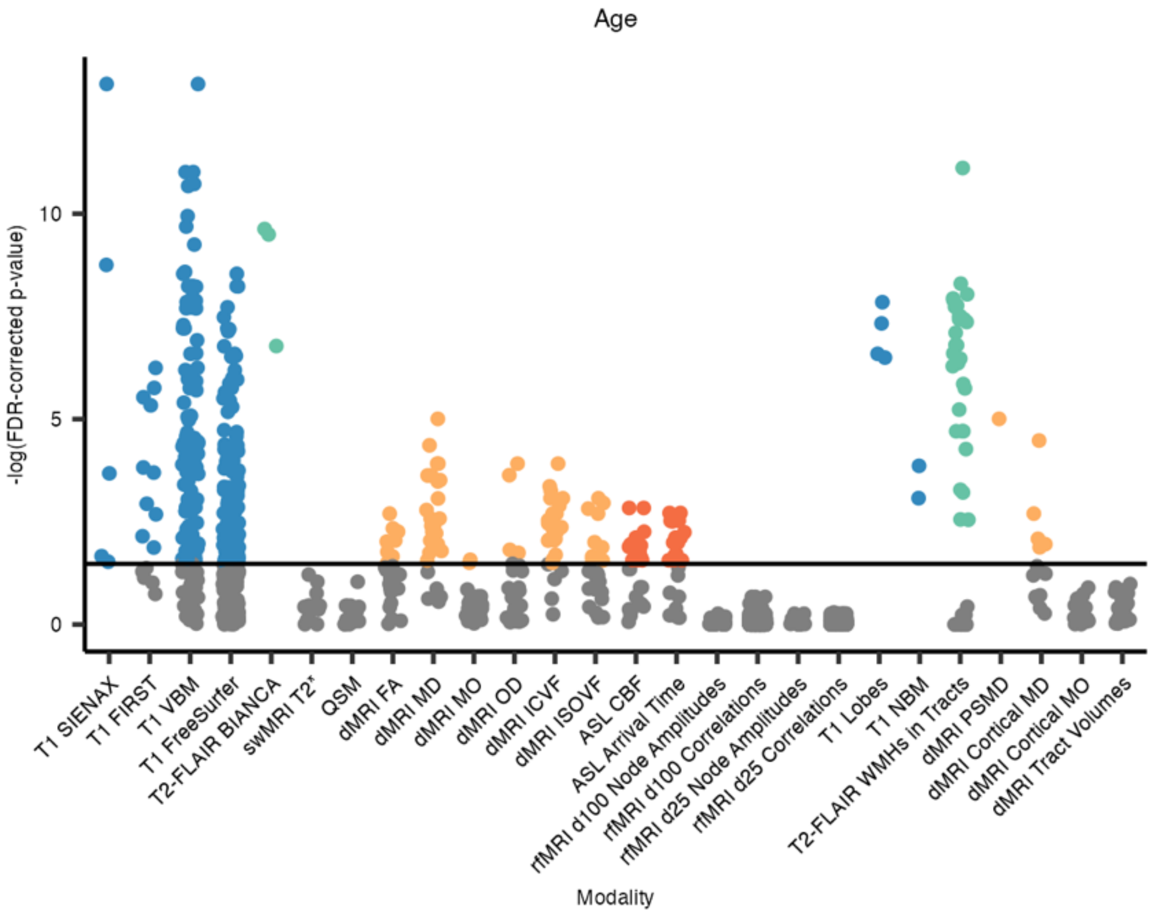

#### Supplementary Figure 5: Beta coefficients of associations with age controlling for volume

Beta coefficients for associations between IDPs and age, controlling for ROI volume with T2* and QSM, and tract volume with tractography-based IDPs (dMRI FA, MD, MO, OD, ICVF, ISOVF). SIENAX, Structural Image Evaluation using Normalization of Atrophy (cross-sectionally); FIRST, FMRIB's Integrated Registration and Segmentation Tool; VBM, voxel-based morphometry; BIANCA, Brain Intensity AbNormality Classification Algorithm; FA, fractional anisotropy; MD, mean diffusivity; MO, mode of anisotropy; OD, orientation dispersion index; ICVF, intra-cellular volume fraction; ISOVF, isotropic volume fraction; CBF, cerebral blood flow; NBM, nucleus basalis of Meynert; WMH, white matter hyperintensity; PSMD, peak width of skeletonised mean diffusivity.

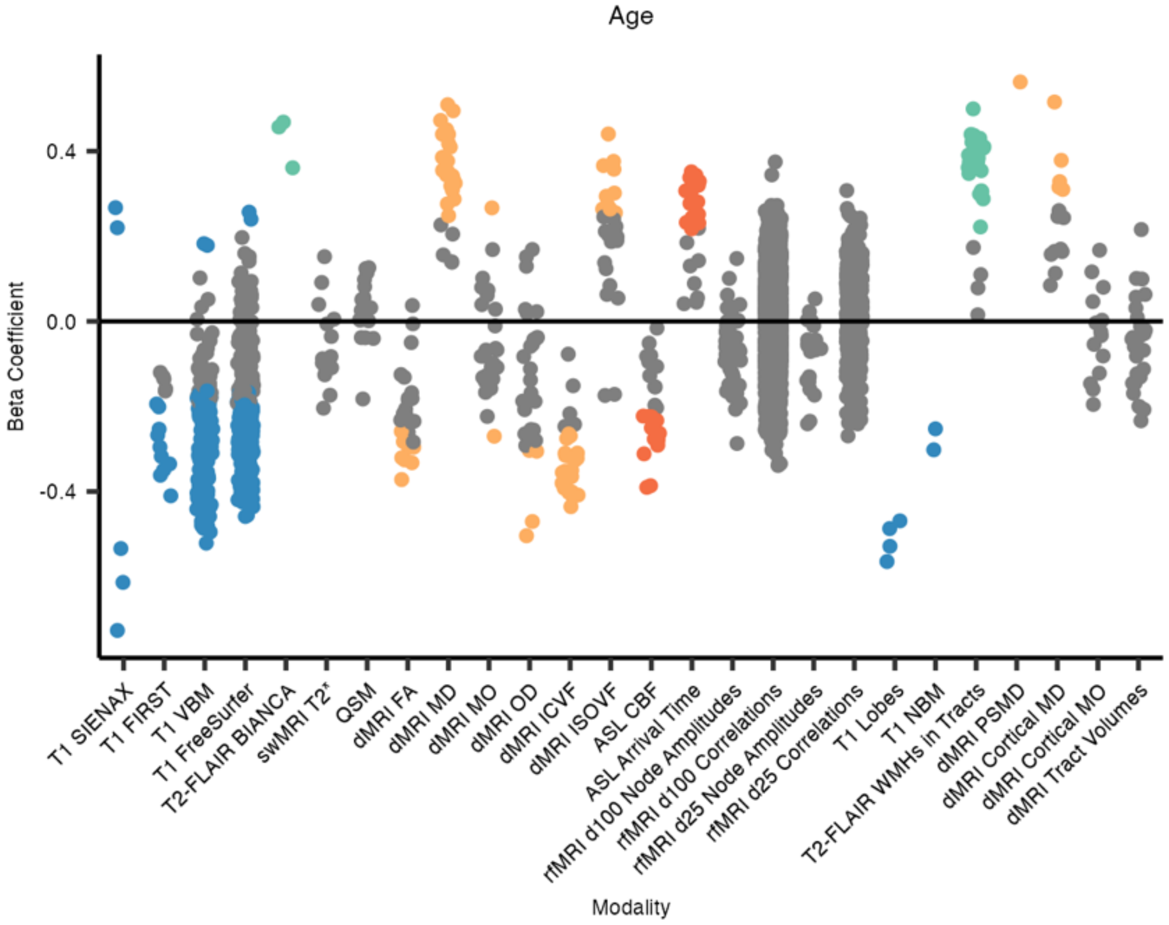

#### Supplementary Figure 6: P-values of associations with cognition controlling for volume

FDR-corrected p-values for associations between IDPs and cognition, controlling for ROI volume with T2* and QSM, and tract volume with tractography-based IDPs (dMRI FA, MD, MO, OD, ICVF, ISOVF). SIENAX, Structural Image Evaluation using Normalization of Atrophy (cross-sectionally); FIRST, FMRIB's Integrated Registration and Segmentation Tool; VBM, voxel-based morphometry; BIANCA, Brain Intensity AbNormality Classification Algorithm; FA, fractional anisotropy; MD, mean diffusivity; MO, mode of anisotropy; OD, orientation dispersion index; ICVF, intra-cellular volume fraction; ISOVF, isotropic volume fraction; CBF, cerebral blood flow; NBM, nucleus basalis of Meynert; WMH, white matter hyperintensity; PSMD, peak width of skeletonised mean diffusivity.

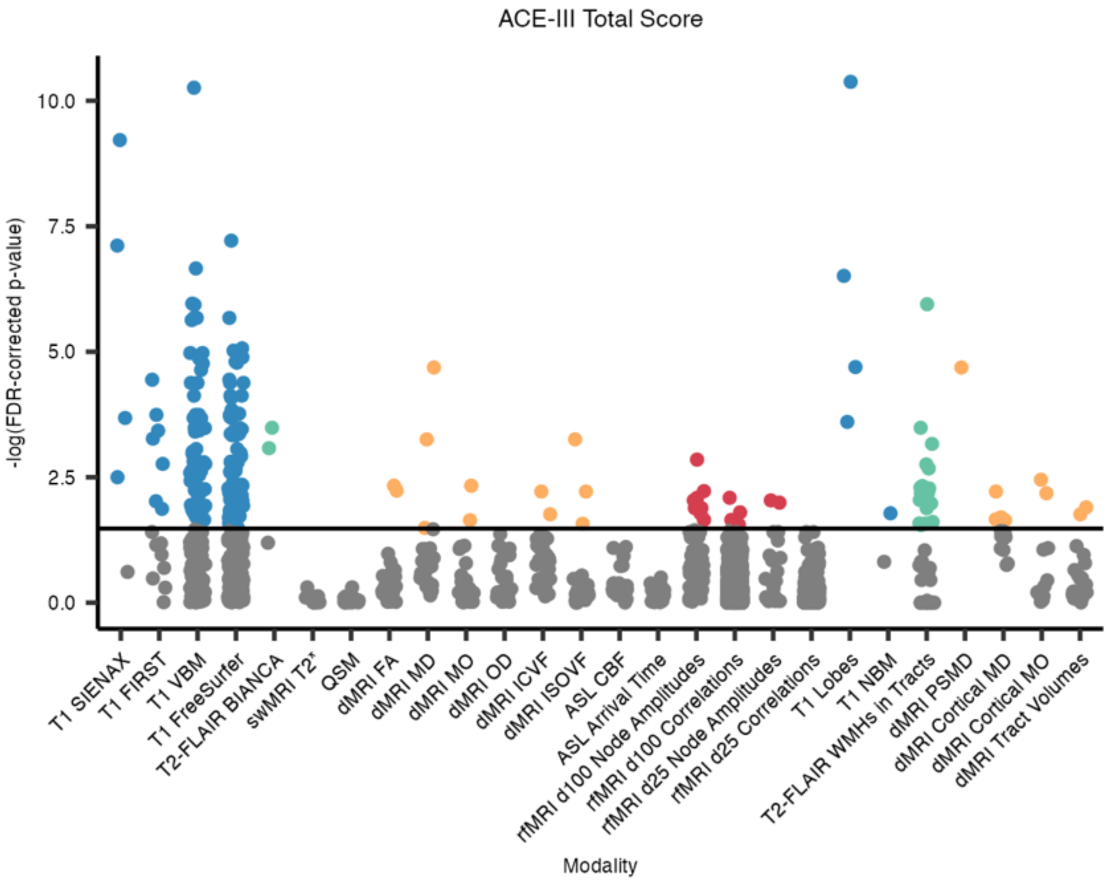

#### Supplementary Figure 7: Beta coefficients of associations with cognition controlling for volume

Beta coefficients for associations between IDPs and cognition, controlling for ROI volume with T2* and QSM, and tract volume with tractography-based IDPs (dMRI FA, MD, MO, OD, ICVF, ISOVF). SIENAX, Structural Image Evaluation using Normalization of Atrophy (cross-sectionally); FIRST, FMRIB's Integrated Registration and Segmentation Tool; VBM, voxel-based morphometry; BIANCA, Brain Intensity AbNormality Classification Algorithm; FA, fractional anisotropy; MD, mean diffusivity; MO, mode of anisotropy; OD, orientation dispersion index; ICVF, intra-cellular volume fraction; ISOVF, isotropic volume fraction; CBF, cerebral blood flow; NBM, nucleus basalis of Meynert; WMH, white matter hyperintensity; PSMD, peak width of skeletonised mean diffusivity.

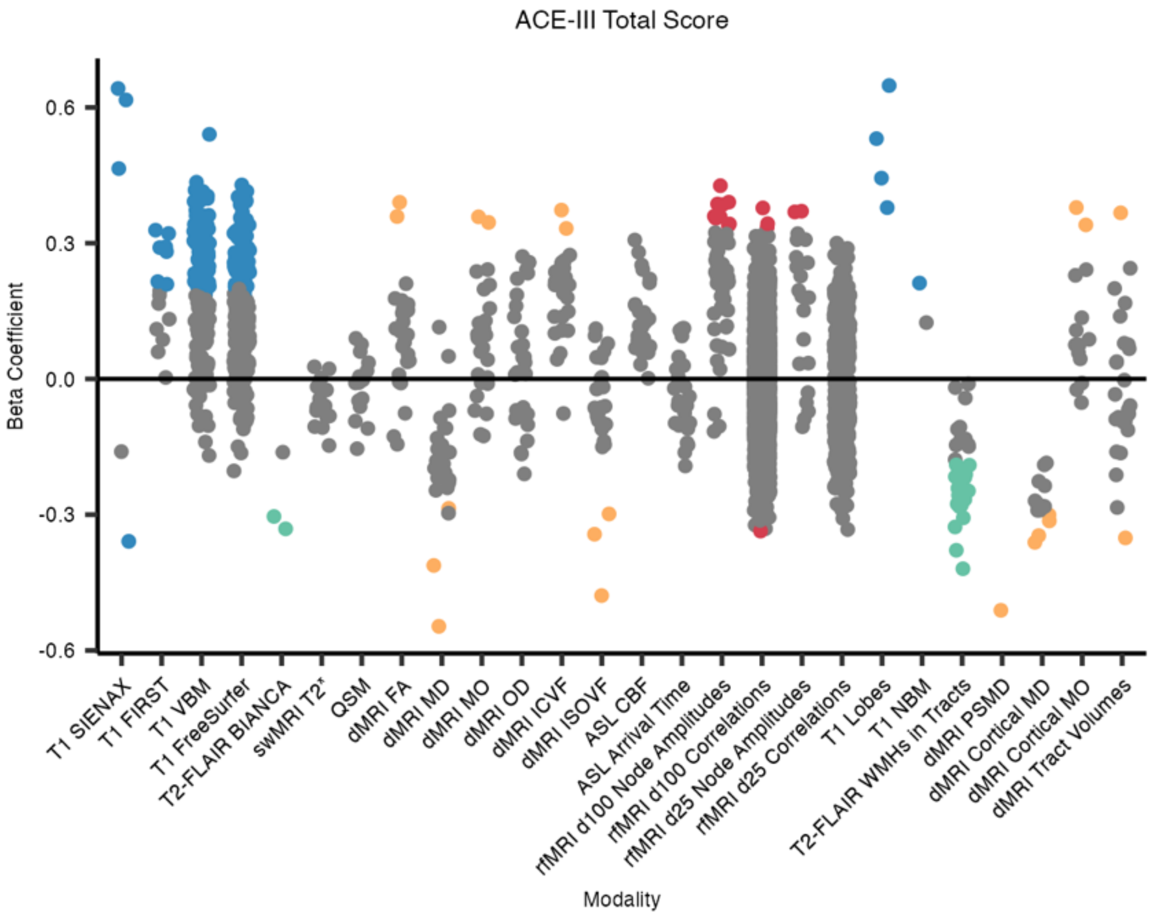
